# Interventions to improve hand hygiene in community settings: A systematic review of theories, barriers and enablers, behavior change techniques, and hand hygiene station design features

**DOI:** 10.1101/2025.03.11.25323730

**Authors:** Sridevi K. Prasad, Jedidiah S. Snyder, Erin LaFon, Lilly A. O’Brien, Hannah Rogers, Oliver Cumming, Joanna Esteves Mills, Bruce Gordon, Marlene Wolfe, Matthew C. Freeman, Bethany A. Caruso

## Abstract

This systematic review identified and examined the theories, barriers and enablers, behaviour change techniques (BCTs), and design features of interventions that have been leveraged effectively to improve and sustain hand hygiene in community settings. It was conducted to support the development of the WHO Guidelines for Hand Hygiene in Community Settings. We searched PubMed, Web of Science, EMBASE, CINAHL, Global Health, Cochrane Library, Global Index Medicus, Scopus, PAIS Index, WHO IRIS, UN Digital Library and World Bank eLibrary for studies published between January 1, 1980, and March 29, 2023, and consulted experts. Eligible studies had an intervention that targeted hand hygiene behaviour, quantitatively measured hand hygiene practice, were published in English after January 1, 1980, and were set in non-healthcare community settings. Studies in healthcare settings, nursing homes, or long-term care facilities were excluded. Two reviewers independently extracted data from each study and assessed risk of bias (Mixed Method Appraisal Tool). 223 eligible studies (including 247,398 participants) met inclusion criteria, 82% of which were effective at improving hand hygiene. A minority (28%) used theory to inform intervention design. Interventions did not always address identified barriers or enablers. Most interventions addressed ‘action knowledge’ (e.g. hand washing instruction), which was not a widely reported barrier or enabler. Interventions did not extensively address the physical environment (e.g., resource availability) despite its importance for hand hygiene. Interventions leveraged a variety of BCT combinations, limiting comparability. We did not conduct a meta-analysis on effectiveness due to heterogeneity across studies. Ten studies reported hand hygiene station design adaptation effectiveness, six examined variations in frequency or intensity of intervention delivery, and four focused on people with disabilities, revealing gaps in evidence. Findings are limited by inconsistent intervention reporting but more consistent identification and leveraging of barriers and enablers would likely improve effectiveness of hand hygiene interventions.

**Funding:** This work was supported by the World Health Organization (PO number: 203046633) and the Foreign and Commonwealth Development Office (FCDO).

PROSPERO registration number CRD42023429145.

**What is already known on this topic:**

- Hand hygiene can prevent infectious diseases, yet little is known about what interventions have been delivered in community settings and if and how they are effective at influencing hygiene behaviours.

**What this study adds:**

- This systematic review examined hand hygiene interventions across community settings to assess if theory informed design and effectiveness, how and if barriers and enablers were leveraged, and to understand what intervention functions, behaviour change techniques (BCTs), and hand hygiene station design features have been tested.
- Most hand hygiene interventions in community settings were found to have been effective, though are not comparable because of variability in setting, focal population, outcome tested, and interventions strategy.
- Despite their effectiveness, interventions did not always address identified barriers or enablers, potentially limiting impact.

**How this study might affect research practice or policy:**

- Evidence from this review demonstrates the need for greater alignment between identified behavioural barriers / enablers and intervention activities.
- Researchers need to improve how they describe and report on interventions to facilitate understanding of what interventions were trying to do, how, and among whom, which can facilitate future learning.
- Further research is needed that includes people with disabilities and to understand how hand hygiene station design adaptions and intervention frequency or intensity influence effectiveness.

## 1. INTRODUCTION

Hand hygiene, whether through handwashing with water and soap or other methods such as the use of alcohol-based hand rubs (ABHR), is a critical public health measure. Hand hygiene interventions are relatively inexpensive to implement [1] and can prevent several infectious diseases, including enteric [2] and respiratory [3] infections, which account for a large burden of disease [4] and high healthcare costs [5]. Establishing global guidelines and recommendations is essential to guide hand hygiene initiatives, protect public health, and strengthen resilient health systems [6].

According to the Ottawa Charter, community settings are where ‘health is created and lived by people within the setting of their everyday life; where they learn, work, play, and love,’ [7] and include domestic, public, and institutional spaces [8]. Guidelines for hand hygiene in healthcare settings are well-established [9–12], and additional guidelines emphasize investing in hand hygiene as a core public health measure [13–16]. Despite the recognized importance of hand hygiene, and the unprecedented prioritization driven by the COVID-19 pandemic [16], gaps and inconsistencies remain in global guidance on specific measures [17]. A recent scoping review identified 51 existing international guidelines and highlighted a lack of consistent evidence-based recommendations and identified four areas where clear recommendations are needed for hand hygiene in community settings: (1) effective hand hygiene; (2) minimum requirements; (3) behavior change; and (4) government measures [8].

In their review, MacLeod et al. (2023) found few international guidelines provided recommendations related to hand hygiene behavior change, indicating a need for global guidance based on existing evidence and established behavioral theory. However, current understanding of which hand hygiene interventions in community settings are effective is limited, constraining the creation of such guidance. While evidence reviews do exist, their scopes are limited. For example, reviews that have synthesized behavior change theories and techniques used to design hand hygiene interventions are limited to settings with children [18,19] and evidence synthesized on hand washing station designs or adaptations are limited to tippy taps and nudges, respectively [20,21]. Recent reviews that have synthesized measures to promote hand hygiene focus specifically on COVID-19 [22,23]. While these reviews and syntheses are informative, there is a need for a systematic review that identifies the theories, intervention functions, behavior change techniques, and design features that have been effective across all public settings.

The goal of this systematic review was to comprehensively examine the theories, barriers and enablers, intervention functions, behavior change techniques, and design features of interventions that have been leveraged effectively to improve and sustain hand hygiene in community settings. The priority question and sub-questions for this review were generated through an extensive consultation process by WHO with external experts [24,25], following a scoping review of current international guidelines [8].

## 2. METHODS

### 2.1. Research questions

This systematic review sought to assess interventions to improve hand hygiene in community settings by answering the following seven questions: (A) Which have been designed using behavior change theories? (B) Which have effectively leveraged identified barriers and enablers of hand hygiene in community settings? (C) What behavior change techniques have been implemented to effectively improve and sustain handwashing practices? (D) What hand hygiene station designs have been effective at improving and sustaining hand hygiene? (E) What hand hygiene station design adaptations (e.g., placement, nudges, and cues) have been effective at improving and sustaining hand hygiene? (F) What level of frequency and intensity of behavior change interventions is necessary to effectively improve hand hygiene? (G) How do hand hygiene practices vary by population groups, risk scenarios or over time?

### 2.2. Search strategy

This review was pre-registered with PROSPERO (registration number: CRD42023429145) and is reported following the Preferred Reporting Items for Systematic Reviews and Meta-Analyses [26] (PRISMA) criteria (See S1 – PRISMA Checklist). This review was part of an integrated protocol for multiple related reviews to synthesize the evidence for effective hand hygiene in community settings [25]. We adopted a two-phased approach for identifying relevant studies. Phase 1 involved a broad search to capture all studies on hand hygiene in community settings that were relevant across multiple related systematic reviews. The outcome of phase 1 was a reduced sample from which further screening, specific to this review, was performed. A full description of the procedures followed for searches, study inclusion, outcomes data collection, analysis, and reporting of the multiple related reviews is presented in the published protocol [25].

This search included studies published between January 1, 1980, and March 29, 2023, and published in English—unless the title and abstract was published in English and/or a non-English language article was referenced in an existing systematic review. We searched 12 peer-reviewed and grey literature databases. PubMed, Web of Science, EMBASE (Elsevier), CINAHL (EBSCOhost), Global Health (CAB), Cochrane Library, Global Index Medicus, Scopus (Elsevier), Public Affairs Information Service (PAIS) Index (ProQuest) were searched on March 23, 2023 and WHO Institutional Repository for Information Sharing (IRIS), UN Digital Library, and World Bank eLibrary were searched on March 28, 2023 using search terms related to hand hygiene broadly and restrictions on terms related to healthcare settings in the titles. We searched trial registries (International Clinical Trials Registry Platform, clinicaltrials.gov) for trials related to hand hygiene in community settings on March 29, 2023.

We conducted manual searches of reference lists of four relevant systematic reviews [18–21]. We only searched references for articles that were included in the respective reviews. These reviews included 82 eligible references of which 46 were duplicates, 17 were already identified in our database search, and 19 were added to phase 2 title and abstract screening. We contacted 35 content experts and organizations, using snowballing methods, between April and May 2023 for information on relevant unpublished literature.

### 2.3. Selection criteria

Eligible study designs were mixed methods studies, randomized and non-randomized control trials, or before-after studies of interventions to improve hand hygiene in community settings.

Hand hygiene is defined as any hand cleansing undertaken for the purpose of removing or deactivating pathogens from hands and efficacious hand hygiene is defined as any practice which effectively removes or deactivates pathogens from hands and thereby has the potential to limit disease transmission [10]. Consistent with MacLeod et al. (2023), the term ‘community settings’ includes locations where healthcare is not routinely delivered, and broadly spans all places where people ‘learn, play, work and love’, including domestic (e.g., households), public (e.g., markets, public transportation hubs, vulnerable populations [e.g., people experiencing homelessness], parks, squares, or other public outdoor spaces, shops, restaurants, and cafes), and institutional (e.g., workplace, schools and universities, places of worship, prisons and places of detention) spaces [8]. Studies were excluded if they were in healthcare settings or were animal research. Studies in nursing homes and long-term care facilities were excluded as part of phase 2 screening as these were determined to be locations where healthcare is delivered. There were no geographic restrictions.

We used Covidence software for systematic reviews [27]. In both phases, screening of each article (phase 1 – title and abstract only; phase 2 – title and abstract, then full text review) was performed independently by two reviewers, with discordance between reviewers reconciled by a third reviewer. To identify linked studies, one reviewer screened all included studies and mapped the intervention names, descriptions, trial registration numbers (if applicable). Linked studies were grouped together based on intervention name or trial registration number. During this mapping process, the studies included were reassessed to confirm they met the eligibility criteria. Studies marked for exclusion were independently assessed by two reviewers to confirm exclusion. The stages and related outcomes of the search and screening process are described in the PRISMA flow chart (S2 – Figure 1).

**Figure 1.**
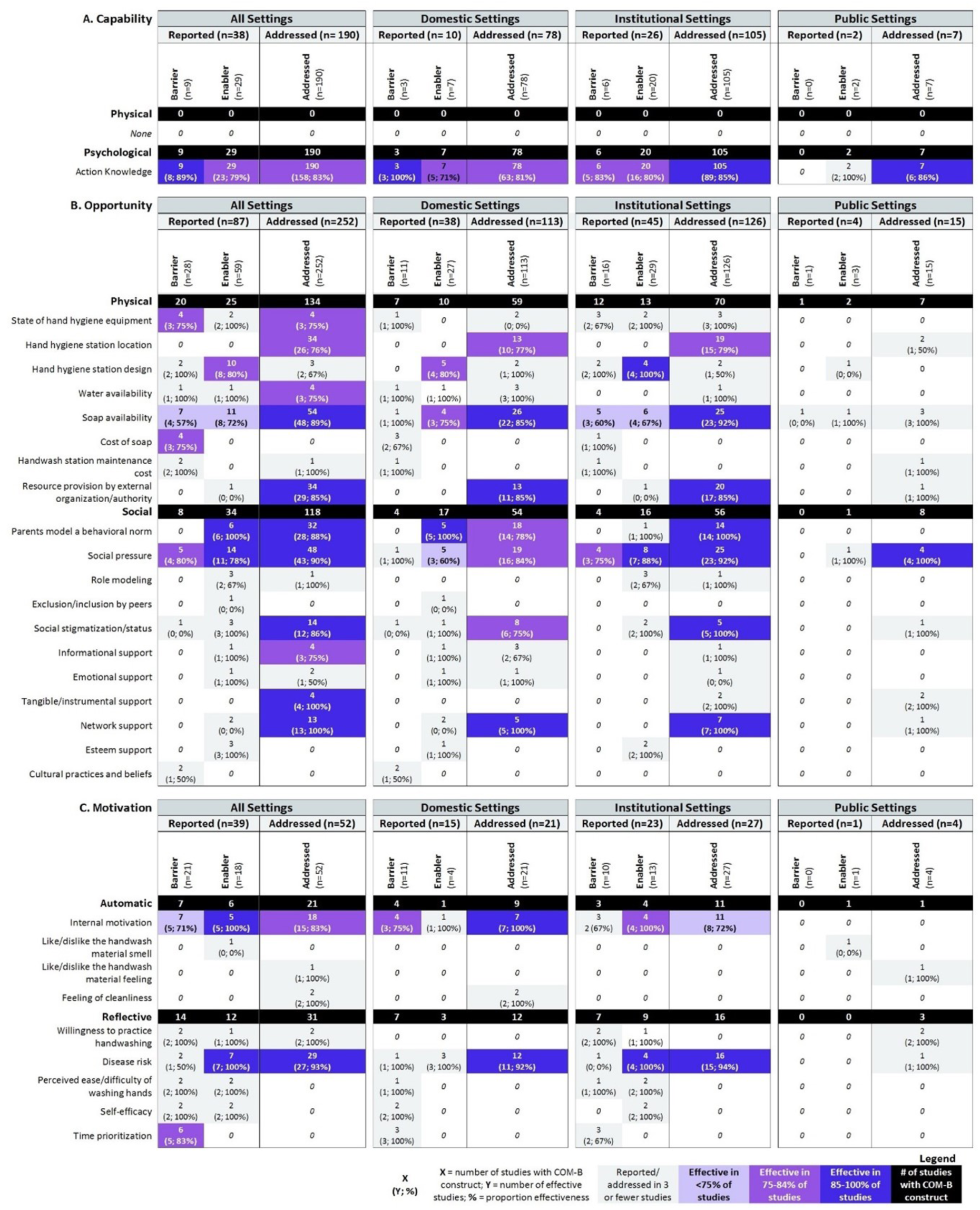
Summary of effective barriers and enablers targeted in hand hygiene interventions and categorized by (A) Capability, (B) Opportunity, and (C) Motivation components of the COM-B framework (N=223). If study authors explicitly mentioned barriers or enablers that influenced their intervention design, these were captured as ‘Reported barrier/enabler’. Otherwise, review authors assumed what barriers or enablers were addressed by the intervention design which were captured as ‘Addressed’. There could have been multiple barriers/enablers reported per study as studies could report multiple intervention categories. Proportion effectiveness refers to the number of studies that reported the intervention was effective at improving hand hygiene outcomes related to that barrier/enabler. Of those that were reported effective, we also provide proportions by setting type (domestic, institution, or public). Effectiveness proportions were only provided for barriers/enablers that had at least 3 or more studies reported/deduced.

### 2.4. Data analysis

Reviewers [SKP, EL] independently double extracted data from each included study using a customized data extraction tool and the Mixed Method Appraisal Tool (MMAT) for assessing risk of bias [28,29]. For appraising the quality of mixed methods studies, the individual components were assessed using the appropriate categories: the qualitative component, the quantitative category for the quantitative component, and the mixed-methods category. Final scores were determined based solely on the quantitative components of the MMAT assessment, given that only quantitative data were extracted from the mixed methods studies included in the review. Any conflicts between reviewers over data extraction and bias assessment were resolved by discussion while re-checking the source. All data extraction templates are provided in the supplement (S3 – Covidence extraction sheet; S4 – Outcome extraction sheet.

To provide a broad overview of included studies, we extracted information about study characteristics, including the region, location, setting, primary research population, primary outcome, hand hygiene practice assessed, and intervention categories. Table 1 provides definitions for categories used to code study interventions.

**Table 1.** Definitions and examples of intervention categories and constructs from the COM-B framework^1^ used to code study interventions.

| Intervention category | Definition | Examples |
| --- | --- | --- |
| **Material provision** | Provision of material resources to aid in handwashing | • Provision of soap<br>• Provision of alcohol-based hand rub<br>• Handwashing station installation |
| **Education programs** | Curriculum-based teaching programs, where instruction is delivered using lectures | • School-based education<br>• Food hygiene education<br>• Education – Adults/Community-based |
| **Training programs** | Demonstration-based teaching programs, where instruction is delivered using interactive activities (e.g., Glo-Germ demonstrations, food preparation demonstrations, etc.) | • School-based training<br>• Food hygiene training<br>• Training – Adults/Community-based |
| **Multimedia messaging** | Media-based communication and messages used to improve handwashing | • Posters<br>• Videos |

|  |  | • SMS<br>• Radio messages<br>• Art installations/flipcharts<br>• Pamphlets<br>• Multiple media types |
| --- | --- | --- |
| **Hygiene promotion** | Community-based programs to promote handwashing that encompass different interactive activities | • Games or competitions<br>• Plays, skits, songs or dramas<br>• Group discussions<br>• Policy development or institutional strengthening<br>• Public pledging ceremonies<br>• Household visits from community health workers<br>• Diaries and journals<br>• Provision of incentives (gift cards, face masks, creams, etc.) |
| COM-B construct | Definition<sup>2</sup> | Examples |
| **Capability** | **Knowledge, skills and abilities required to engage in a particular behavior** |  |
| **Physical Capability** | Physical strength, skill or stamina | • Physical ability to wash hands |
| **Psychological Capability** | Knowledge/ psychological strength, skills or stamina | • Knowledge on how to wash hands |
| **Opportunity** | **External factors which make the execution of a particular behavior possible** |  |
| **Physical Opportunity** | Opportunities provided by the environment, such as time, location and resource | • Availability of soap |
| **Social Opportunity** | Opportunities as a result of social factors, like cultural norms and social cues | • Cultural norm to wash hands |
| **Motivation** | **The internal processes which influence decision making and behaviors** |  |
| **Automatic Motivation** | Automatic processes, such as our desires, impulses and inhibitions | • Have habit of washing hands |
| **Reflective Motivation** | Reflective processes, such as making plans and evaluating things that have already happened | • Believe handwashing reduces illness risk |
| COM-B Intervention Function | Definition<sup>1</sup> | • Examples |
| **Education** | Increasing knowledge or understanding | • Curriculum on hand hygiene |
| **Persuasion** | Using communication to induce positive or negative feelings or stimulate action | • Multimedia messaging (e.g., flyers, radio messages) drawing on disgust or fear to promote handwashing |
| **Incentivization** | Creating an expectation of reward | • Providing prizes for people who wash hands |
| **Coercion** | Creating an expectation of punishment or cost | • Shaming people who do not wash hands |
| **Training** | Imparting skills | • Workshops demonstrating how to wash hands |
| **Restriction** | Using rules to reduce opportunity to engage in target behavior/Increase target behavior by reducing opportunity to engage in competing behaviors | • Rules that children have to wash hands before eating lunch |
| **Environmental Restructuring** | Changing physical or social context | • Provision of soap<br>• Installation of handwashing station |
| **Modeling** | Providing example for people to aspire to or imitate | • Videos showing proper handwashing technique<br>• Community leaders demonstrating proper handwashing technique |
| **Enablement** | Increasing means/reducing barriers to increase capability or opportunity (beyond education, training, or environmental restructuring) | • Community health workers conducting weekly visits to bring handwashing materials and provide additional engagement on hand hygiene |

Note: <sup>1</sup>Definitions from (Michie, van Stralen, and West 2011); <sup>2</sup>Definitions from (Social Change UK)

To determine intervention effectiveness, we assessed whether authors of the included studies either reported their intervention to have a statistically significant effect in improving hand hygiene outcomes or if they reported an increase in hand hygiene outcome comparing the treatment group to a control scenario.

To understand the proportion of studies that reported the use of a behavioral theory in the intervention design (Question a), we extracted information on what theories were identified as being used, and report on the frequency of theory use and the proportion of the interventions that were effective at improving hand hygiene outcomes.

To determine which interventions effectively leveraged identified barriers and enablers to hand hygiene outcome(s) in community settings (Question b), we first extracted data on which barriers and enablers were *reported* by study authors to influence the focal hand hygiene behavior. Beyond what study authors reported, we also determined what barriers and enablers the described interventions *addressed*; which are referred to in subsequent tables as ‘reported’ and ‘addressed’ barriers and enablers. The *reported* and *addressed* barriers and enablers were independently identified by two reviewers and then reconciled if there was disagreement. Because an intervention component could address the same component regardless of whether it was reported to be a barrier or enabler, we present addressed barriers and enablers as one category. For example, if an intervention provides soap, this addresses both a potential barrier (lack of soap) and a potential facilitator (presence of soap). We categorized all identified reported barriers and enablers, and whether or not they were addressed according to the constructs of the COM-B framework. This framework identifies capability, opportunity, and motivation as essential conditions that interact to enable behavior, using a codebook that was pre-reviewed by one of the creators of the COM-B framework [31]. Definitions for each of the constructs from the COM-B framework are provided in Table 1. Finally, we used intervention effectiveness data to understand which barriers and enablers were effectively leveraged in domestic, institutional, or public settings.

To determine which behavioral change techniques (BCTs) were implemented and which were effective at improving and sustaining handwashing practices in community settings (Question c), we used the Behavioral Change Techniques Taxonomy, version 1 [32] to identify and categorize BCTs used in each study. A BCT is ‘an observable and replicable component [of an intervention] designed to change behavior. It is the smallest component compatible with retaining the postulated active ingredients and can be used alone or in combination with other BCTs’ [33]. Because interventions can leverage one or more BCT, for each intervention, we identified the ‘BCT Package’, defined as the suite of BCTs used for the described interventions, for each intervention. We then determined the frequency of each BCT package and the proportion that were effective by community setting type. Finally, we identified the intervention function(s), the broad categories of BCTs, associated with each BCT package [33]. Definitions for each of the intervention functions, which are used to categorize BCTs in the COM-B framework, are provided in Table 1. Table 1 also provides definitions for the intervention categories, which comprise the actual activities that studies implemented to improve handwashing practice.

To determine which hand hygiene station designs (Question d) and adaptations (Question e) have been effective at improving and sustaining hand hygiene, we identified all interventions that included hand hygiene stations and extracted data on reported design features and their effectiveness at improving and sustaining hand hygiene practices in community settings. We identified all interventions studies that reported assessing hand hygiene station adaptations and extracted data and reported on which adaptations were effective.

To determine the level of frequency and intensity of behavior change interventions that have been effective at improving hand hygiene (Question f), we first identified all interventions that indicated varying the frequency or intensity of their behavior change intervention. Among those we extracted data on frequency and intensity and report which were effective at improving hand hygiene.

To assess variability by population groups, risk scenarios and over time, we extracted data on focal population groups, risk scenarios, and time to report on variability.

Due to the heterogeneity of the studies’ interventions and outcome measures, a meta-analysis was not conducted on the associations between theories, barriers and enablers, intervention functions and behavior change techniques, or on design features to improve and sustain hand hygiene.

### 2.5 Ethics and patient involvement statements

As this is a review of published documents, no ethical approval was required. Patients or the public were not involved directly in the design, or conduct, or reporting, or dissemination plans of our research. This evidence synthesis supports the forthcoming WHO Guidelines for Hand Hygiene in Community Settings, which developed the study questions in broad consultation with key partners and networks of partners.

### 2.6 Data Availability

All data and extraction templates will be made publicly available upon publication at Figshare.

## 3. RESULTS

### 3.1. Characteristics of the studies included in this review

We identified 260 reports of eligible studies, derived from 223 studies meeting our inclusion criteria once duplicate mention of interventions in different studies were grouped; 93% (208) were quantitative studies and 7% (15) were mixed methods studies (S2 – PRISMA flow chart and S5 – Overview of included studies). We found studies in all WHO regions, with the highest representation from Africa (29%), South-East Asia (28%) and the Americas (22%) (Table 2). Most studies delivered interventions in domestic (45%) or institutional (54%) settings with very few delivered in public settings (4%) (some delivered in multiple settings). Schools (37%) and universities (10%) were the most common locations after households. Other institutional settings included workplaces (5%) and childcare centers (1%) and public settings represented were markets (2%), internally displaced persons camps (1%), community health clubs (1%), and public transportation hubs (1%). More than half of the included studies (72%) identified hand hygiene as the primary outcome of interest and 52% of studies targeted other behaviors in addition to hand hygiene. The largest proportion of studies (33%) focused on both girls and boys; 25% did not clearly specify the focal population, 20% focused on women only, and 20% focused on women and men.

**Table 2.** Characteristics of the included studies (N=223).

| Descriptive characteristics of studies | Total<br>n (%) |
| --- | --- |
| **Total number of studies** | **223 (100.0)** |
| **Region\*** |  |
| African Region | 64 (28.7) |
| South-East Asian Region | 63 (28.3) |
| Region of the Americas | 49 (21.9) |
| Western Pacific Region | 27 (12.1) |
| European Region | 22 (9.9) |
| Eastern Mediterranean Region | 8 (3.6) |
| Unspecified | 1 (0.5) |
| **Location\*** |  |
| Rural | 84 (37.7) |
| Urban | 62 (27.8) |
| Peri-urban | 13 (5.8) |
| Unspecified | 83 (37.2) |
| **Setting\*** |  |
| Domestic (Households) | 101 (45.3) |
| Institutional | 120 (53.8) |
| Childcare centers | 3 (1.3) |
| Schools | 83 (37.2) |
| Universities | 22 (9.9) |
| Workplaces | 12 (5.4) |
| Public | 8 (3.6) |
| Markets | 4 (1.8) |
| Internally displaced persons camps | 2 (0.9) |
| Community health clubs | 1 (0.4) |
| Public transportation hubs | 1 (0.4) |
| Unspecified | 1 (0.4) |
| **Gender of primary research population** |  |
| Adults – Men | 3 (1.3) |
| Adults – Women | 45 (20.2) |
| Adults – Women and men | 43 (19.2) |
| Children – Boys | 3 (1.3) |
| Children - Girls | 2 (0.9) |
| Children – Girls and boys | 72 (32.7) |
| Unspecified | 55 (24.7) |
| **Primary outcome studied\*** |  |
| Hand hygiene | 161 (72.2) |
| Diarrheal diseases | 19 (8.5) |
| Food hygiene | 21 (9.5) |
| Nutrition | 17 (7.6) |
| Respiratory infections | 14 (6.3) |
| Other infectious diseases | 12 (5.4) |
| Other WASH-related behaviors | 12 (5.4) |
| Other non-infectious health outcomes | 11 (3.9) |
| School absenteeism | 8 (3.6) |
| Mental/social well-being | 3 (1.3) |
| Soil-transmitted helminths | 1 (0.4) |
| Work absenteeism | 1 (0.4) |
| **Specific hand hygiene practice assessed\*** |  |
| Handwashing with soap and water | 143 (64.1) |
| Handrubbing with alcohol-based rub | 19 (8.5) |
| Handwashing with water only | 15 (6.7) |
| Handwashing with soap alternatives | 5 (2.2) |
| Hand drying | 3 (1.3) |
| Handwashing with non-alcoholic antiseptics | 3 (1.3) |
| Handwashing - unspecified | 63 (28.3) |
| **Intervention also targeted other behaviors aside from hand hygiene** | **112 (50.2)** |
| **Hand hygiene intervention category\*\*** |  |
| **Material provision** | **89 (39.9)** |
| Provision of soap | 53 (23.7) |
| Installation of handwashing stations | 46 (20.6) |
| Provision of alcohol-based hand rub | 16 (7.2) |
| **Education programs** | **99 (44.4)** |
| School-based education | 44 (19.7) |
| Food hygiene education | 3 (1.3) |
| Education – Adults/community-based | 52 (23.3) |
| **Training programs** | **86 (38.6)** |
| School-based training | 35 (15.7) |
| Food hygiene training | 8 (3.6) |
| Training - Adults/community-based | 43 (19.3) |
| **Multimedia messaging** | **93 (41.7)** |
| Posters | 38 (17.0) |
| Videos | 19 (8.5) |
| Art installations/flipcharts | 8 (3.6) |
| Radio messages | 7 (3.1) |
| Pamphlets | 5 (2.2) |
| SMS | 3 (1.3) |
| Multiple | 10 (4.5) |
| Unspecified*** | 4 (1.8) |
| **Hygiene promotion** | **60 (26.9)** |
| Policy development or institutional strengthening | 12 (5.4) |
| Household visits from community health workers | 10 (4.5) |
| Group discussions | 9 (4.0) |
| Plays, skits, songs or dramas | 6 (2.7) |
| Games or competitions | 4 (1.8) |
| Diaries and journals | 4 (1.8) |
| Provision of other incentives (gift cards, creams, face masks, prizes, etc. | 3 (1.3) |
| Public pledging ceremonies | 2 (0.9) |
| Other | 12 (5.4) |
| **Study quality rating (mean)** | **3.3** |

**Note:** \*Descriptive statistics for region, location, setting, primary outcome studied, hand hygiene practice, and hand hygiene intervention represent variables that were multi-select. \*\*Interventions are not mutually exclusive so studies could have multiple intervention categories as well as multiple sub-categories included per study. \*\*\*Multimedia-unspecified refers to interventions where studies reported that they used media messaging but did not specify the mode of delivery.

Handwashing with soap and water was the most common hand hygiene practice targeted by interventions (64%), followed by hand cleansing with alcohol-based rub (9%); 28% of studies did not provide detail about the specific hand hygiene practice of focus (n=63). Hand hygiene intervention categories varied by study; none of the category types were dominant. Specifically, 40% of interventions included material provision (e.g., soap, hand hygiene stations), 44% included education (e.g., school-based education), 39% included trainings (e.g., food hygiene trainings), 42% included multi-media messaging (posters, videos), and 27% included other hygiene promotion activities (e.g., household visits from community health workers; group discussions, games). Soap was only provided in 24% of interventions. Overall, 82% of interventions were found to be effective. Additional study characteristics and the studies leveraged for each research sub-question can be found in the supplement (S5 – Overview of studies; S6 – Studies leveraged for each research question).

### 3.2. Quality of the studies included in this review

The mean study quality was 3.3 out of 5, overall indicating good quality (3.2 for non-randomized studies (n=121), 3.4 for randomized control trials (n=87), and 3.6 for mixed methods studies (n=15)). Quality appraisal scores for each included study are included in the supplement (S7 – MMAT Assessment).

### 3.3. Evidence to answer sub-questions

#### 3.3.a. (Question A) Among interventions to improve hand hygiene in community settings, which have been designed using behavior change theories?

To answer which interventions were designed with behavior change theories, we extracted which theories were identified as being used. Of the 223 studies identified in our review, 28% (n=63) reported using a theory or model to inform the design of the hand hygiene intervention (S8 – Theory table). The earliest study included in our review that reported using theory was published in 2009, but there is no noticeable change in the proportion of studies that have used theory over time (S9 – Theory bar chart). A greater proportion of studies in public settings (3/8; 38%) used theory, compared to institutional (35/120; 29%) and domestic (25/101; 25%) settings, but there were few studies conducted in public settings. There was no difference in effectiveness among interventions using theory (52/63; 83%) and those not (131/160; 82%). The theories, models, and frameworks most commonly referenced include the *Theory of Planned Behavior* (n=14; 22%; 71% effective) [34], *Health Belief Model* (n=11; 18%; 91% effective) [35], *Behaviour Centered Design/Evo-Eco Model* (n=7; 11% 71% effective), [36], *RANAS (Risks, Attitudes, Norms, Abilities, and Self-regulation*) (n=6; 10%; 83% effective) [37], *COM-B (Capability, Opportunity, Motivation, and Behavior)* (n=6; 10%; 83% effective) [31], and *IBM-WASH (Integrated Behavioural Model for Water, Sanitation, and Hygiene)* (n=5; 8%; 80% effective) [38]. Other behavior change theories, models, and frameworks were noted (n=19); however, each one was only referenced across one or two studies. S5 indicates which theory was used for each study, if any.

#### 3.3.b. (Question B) Among interventions to improve hand hygiene in community settings, which have effectively leveraged identified barriers and enablers of hand hygiene in community settings?

To answer this question, we extracted the barriers and enablers that were *reported* by study authors as well as those that the described interventions *addressed*. In all settings, barriers and/or enablers were identified across all broad COM-B constructs and most sub-constructs (Figure 1). Overall, studies largely reported a given barrier or enabler less often than it was addressed. In other words, interventions acted on a given barrier or enabler more often than the barrier or enabler was explicitly acknowledged to be an issue. Studies did not always report the same barrier and enabling factor for a given COM-B construct. Representative examples of reported and addressed barriers/enablers by COM-B component can be found in the supplement (S10 – Examples of addressed vs reported misalignment).

Under Capability, no studies reported any *Physical Capability* barriers or enablers in any setting, and none were addressed. ‘Action Knowledge’ was identified as the only reported or addressed *Psychological Capability* barrier or enabler. Across all settings, ‘Action knowledge’ was reported to be a barrier or enabler far less frequently than it was addressed, indicating that interventions acted on ‘Action knowledge’ far more often than they reported it as a precondition. Specifically, ‘Action Knowledge’ was noted as a barrier in just 9 studies (3 Domestic settings [D], 6 Institutional settings [I], 0 Public settings [P]) and as an enabler in 29 studies (7 D, 20 I, 2 P). However, it was addressed in 190 (78 D, 105 I, 7 P). Across all studies and settings in which ‘Action Knowledge’ was either reported or addressed to be a barrier or an enabler, 70% or more interventions were found to be effective.

Under Opportunity, several *Physical* and *Social Opportunity* barriers and enablers were reported and addressed across settings, yet these were reported or addressed least often among those based in public settings. The *Physical Opportunities* barriers/enablers most frequently reported or addressed include: ‘State of hygiene equipment’, ‘Hand hygiene station location’, ‘Soap availability’, and ‘Resource provision’. Those least often reported or addressed included ‘Water availability’, ‘Cost of soap’, and ‘Handwashing station maintenance cost.’ As with ‘Action Knowledge’ (Psychological Capability) there was notable discordance in the number reported and addressed for several barriers/enablers. For example, ‘Soap availability’ was reported as a barrier (7) or enabler (11) less often than it was addressed as a barrier or enabler based on intervention action taken (54). ‘Hand hygiene station location’ was addressed in 34 interventions but was never explicitly reported to be a barrier or enabler. And finally, ‘Hand hygiene station design’ was reported to be an enabler in 10 studies but was only addressed in three, indicating that several of the studies that reported it to be an enabler did not address it in the intervention. Among barriers and enablers classified as *Social Opportunities, ‘*Social pressure’ and ‘Parent model a behavioral norm’ were the most reported and addressed, and across settings in which these were either reported or addressed to be a barrier or an enabler, 60% or more interventions were found to be effective. The least reported *Social Opportunities* barriers/enablers include: ‘Role modeling’, ‘Exclusion/Inclusion by peers,’ ‘Informational support’, ‘Emotional support,’ ‘Tangible/instrumental support,’ ‘Esteem support’, and ‘Cultural practices and beliefs.’

Under Motivation, several *Automatic* and *Reflective Motivation* barriers and enablers were reported and addressed across settings, yet these rarely were reported or addressed among studies based in public settings. ‘Internal motivation’ was the most reported and addressed *Automatic Motivation* factor (reported barrier: 7; reported enabler: 5; addressed: 18). Across all settings in which ‘Internal motivation’ barriers and enablers were either reported or addressed to be a barrier or an enabler, 60% or more interventions were found to be effective. The least reported *Automatic Motivation* barriers/enablers include: ‘Like/dislike of handwash material smell’, ‘Like/dislike of handwash material feeling’, and ‘Feeling of cleanliness.’ ‘Disease Risk’ was the most identified *Reflective Motivation* factor (reported barrier: 2; reported enabler: 7; addressed: 29). Across settings in which ‘Disease risk’ barriers and enablers were either reported or addressed, intervention effectiveness was variable. The least reported *Reflective Motivation* barriers/enablers include: ‘Willingness to practice handwashing,’ ‘Perceived ease/difficulty of washing hands’, ‘Self-efficacy’, and ‘Time prioritization.’

#### 3.3.c. (Question C) Among interventions to improve hand hygiene in community settings, what behavior change techniques have been implemented to effectively improve and sustain handwashing practices?

To identify which behavior change techniques were implemented, we used the Behavioral Change Techniques Taxonomy, version 1 [32] to categorize the BCTs used in each intervention. From the 223 studies, 437 BCT packages were identified, representing 18 different BCT package ‘types’ (S11 – Frequency of BCT packages). Nearly half (45%) of studies evaluated just one package, 28% evaluated two; 17% evaluated three, the remaining 10% evaluated between 4 and 6 packages. Roughly a third (32%) of studies evaluated a package with just one BCT, 75% evaluated packages that had two BCTs, and 49% evaluated a package that had three BCTs.

Among the different package types identified, 17 different BCTs were used (a-q in Table 3), and four BCTs were dominant (Table 4). The most common BCT is ‘a. Instruction on how to perform the behavior’, which was used in five (28%) package types (IDs: 1, 2, 8, 9, 16) accounting for 240 of the 437 packages (55%). The second most common BCT is ‘b. Information about health consequences’, which was used in five (28%) package types (IDs: 1, 2, 12, 14, 17) accounting for 183 of the 437 packages (42%). The third most common BCT is ‘c. Adding objects to the environment’, which was used in five (28%) package types (IDs: 2, 3, 4, 5, 6) accounting for 166 of the 437 packages (38%). Finally, the fourth most common BCT is ‘f. Demonstration of behavior’, which was used in four (22%) package types (IDs: 7, 8, 12, 17) accounting for 99 of the 437 packages (23%). The remaining BCTs featured in 13% or fewer packages (range: 2 (0.5%) - 55 (12.6%)).

**Table 3.** Frequency of individual behavior change techniques (BCTs) used across the package types (N=437)

| BCT | Package type ID | Number of Packages per type evaluated | Inclusion across Packages |
| --- | --- | --- | --- |
| a. Instruction on how to perform the behavior | 1 | 105 | 240 (54.5%) |
|  | 2 | 61 |  |
|  | 8 | 43 |  |
|  | 9 | 19 |  |
|  | 16 | 12 |  |
| b. Information about health consequences | 1 | 105 | 183 (41.8%) |
|  | 2 | 61 |  |
|  | 12 | 10 |  |
|  | 14 | 3 |  |
|  | 17 | 4 |  |
| c. Adding objects to the environment | 2 | 61 | 166 (38.0%) |
|  | 3 | 53 |  |
|  | 4 | 35 |  |
|  | 5 | 9 |  |
|  | 6 | 8 |  |
| d. Restructuring the physical environment | 4 | 35 | 44 (10.0%) |
|  | 5 | 9 |  |
| e. Prompts/cues | 5 | 9 | 29 (6.7%) |
|  | 6 | 8 |  |
|  | 16 | 12 |  |
| f. Demonstration of behavior | 7 | 42 | 99 (22.7%) |
|  | 8 | 43 |  |
|  | 12 | 10 |  |
|  | 17 | 4 |  |
| g. Social support (practical) | 7 | 42 | 55 (12.6%) |
|  | 12 | 10 |  |
|  | 14 | 3 |  |
| h. Feedback on behavior | 8 | 43 | 43 (9.8%) |
| i. Social support (unspecified) | 10 | 12 | 12 (2.7%) |
| j. Restructuring the social environment | 10 | 12 | 12 (2.7%) |
| k. Self-monitoring of behavior | 11 | 7 | 7 (1.6%) |
| l. Goal setting (behavior) | 11 | 7 | 7 (1.6%) |
| m. Action planning | 13 | 9 | 11 (2.5%) |
|  | 15 | 2 |  |
| n. Information on others' approval | 13 | 9 | 11 (2.5%) |
|  | 15 | 2 |  |
| o. Commitment | 15 | 2 | 2 (0.5%) |
| p. Material incentive | 17 | 4 | 4 (0.9%) |
| q. Incentive (outcome) | 18 | 3 | 3 (0.6%) |

**Table 4.**
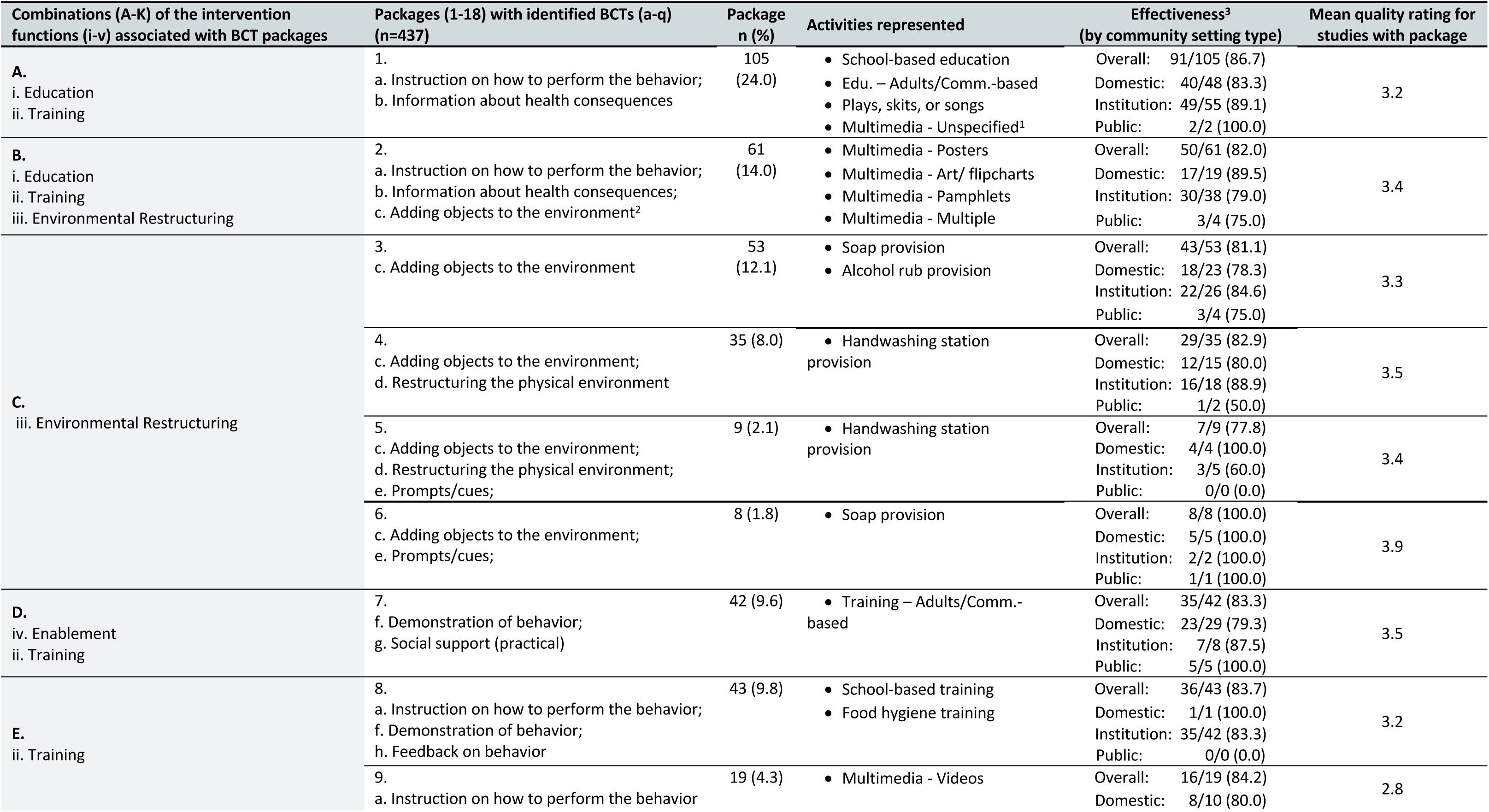
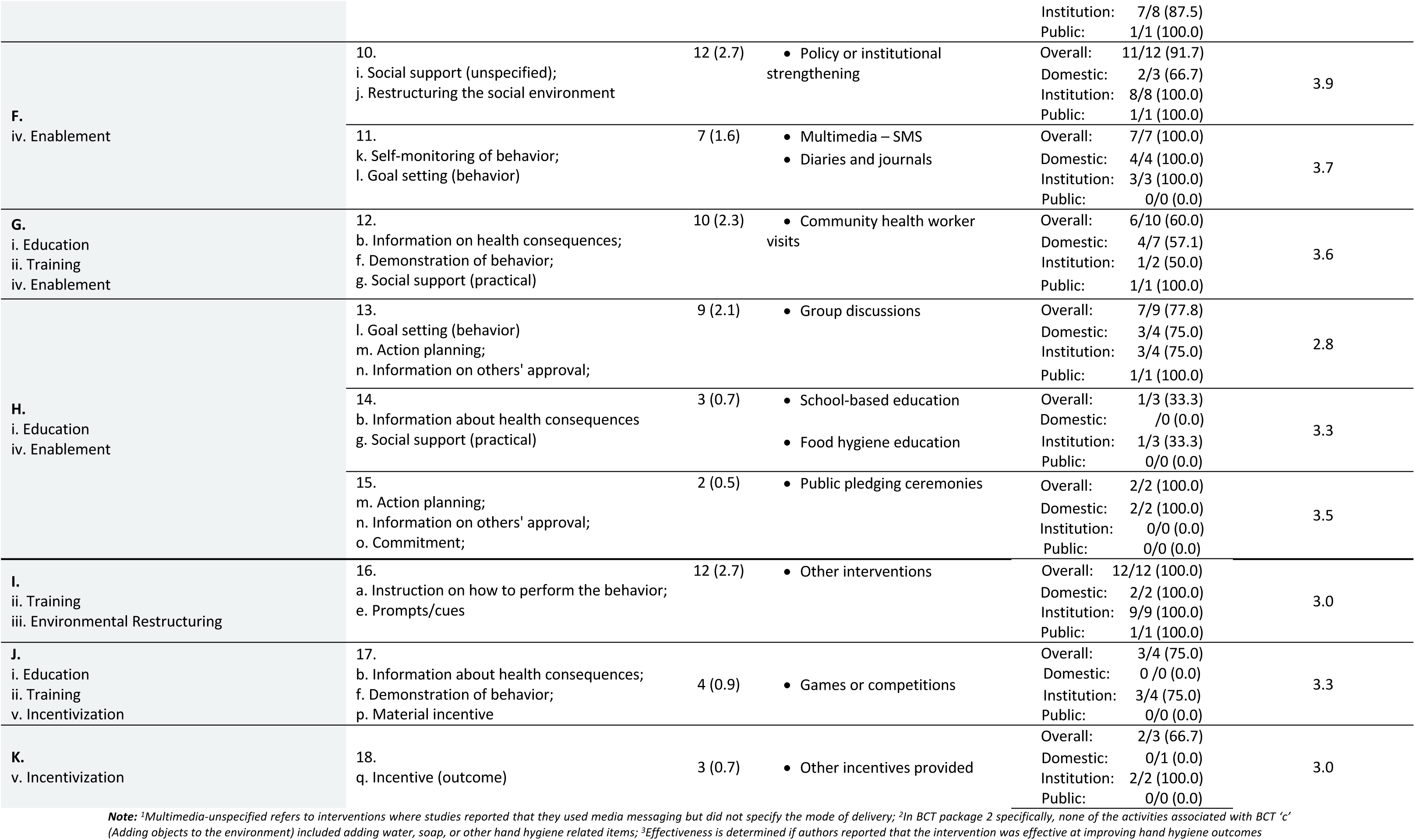
Frequency of identified behavior change technique (BCT) packages and represented activities by associated intervention functions, with their effectiveness and study quality rating across included studies (N=223).

We do not report on effectiveness of every individual BCT as most (14/17; 82%) were only used in combination with other BCTs; thus, isolating the individual contribution of each BCT in a package is not possible. Only three BCTs were evaluated independently, without other BCTs: ‘a. instructions on how to perform the behavior’ (Package 9: overall effectiveness 84.2%, (16/19)); ‘c. Adding objects to the environment’ (Package 3: overall effectiveness 81.1%, (43/53)); and ‘q. Incentive’ (Package 18: overall effectiveness 67.7%, (2/3)) (Table 4). However, use in isolation only accounts for a small proportion of the overall use of BCTs ‘a’ and ‘c’. For BCT ‘a’, its isolated use in Package 9 accounts for only 8% (19/240) of its overall use, and for BCT ‘c’ its isolated use in Package 3 accounts for only 32% (53/166) of its overall use.

Four BCT intervention package types (packages 1-4, Table 4) represent 58% (254/437) of the intervention packages assessed. Packages 1 and 2 leverage Education and Training intervention functions, and Packages 2, 3, and 4 leverage the Environmental Restructuring intervention function. These four packages were generally effective overall and across settings, though the number of evaluations in public settings are limited for all four. Specifically, Package 1, which has two BCTs (a. Instruction on how to perform the behavior and b. Information about health consequences) represents 24% (105) of the packages evaluated. This package was only evaluated in two public settings (100% effective) and was effective in both Domestic (83%; 40/48) and Institutional (89%; 49/55) settings. Package 2 includes three BCTs (a. Instruction on how to perform the behavior, b. Information about health consequences, and c. Adding objects to the environment) and represents 14% (61) of evaluated packages. This package was only evaluated in three public settings (75% effective) and was effective in both Domestic (90%; 17/19) and Institutional (79%; 30/38) settings. Package 3 includes one BCT (c. Adding objects to the environment) and represents 12% (53) of evaluated packages. This package was only evaluated in three public settings (75% effective) and was effective in Domestic (78%; 18/23) and Institutional settings and 85% (22/26) effective in institutional settings. Finally, Package 4, includes two BCTs (c. Adding objects to the environment and d. Restructuring the physical environment) and represents 8% (35) of evaluated packages. This package was only evaluated in two public settings (50% effective) and was effective in Domestic (80%; 12/15) and institutional (89%; 16/18) settings. The intervention functions, behavior change techniques, and intervention activities leveraged in each study can be found in the supplement (S12 – Intervention functions, BCTs, and activities).

#### 3.3.d. (Question D) Among interventions to improve hand hygiene in community settings, what hand hygiene station designs have been effective at improving and sustaining hand hygiene?

To answer which hand hygiene stations were effective at improving hand hygiene, we extracted data on design features from interventions that included hand hygiene stations. Hand hygiene station types and design features were reported in 46 (21%) of the included studies, the majority of which were for users in school (52%) and household (33%) settings. A summary of these designs and their reported effectiveness are provided in the supplement (S13 – HH station designs). Of the 46 studies that reported providing a handwashing station, 37 (80%) were found to be effective. The most common station was a raised bucket with tap/outlet (n=20; 75% effective). Ten (22%) studies did not report the type of station design; among those, 70% were found to be effective.

Hand hygiene station designs varied by several features, including their mobility (fixed vs mobile), permanency (temporary vs permanent), and water supply (individual storage tank vs piped water), among others. A greater proportion of fixed hand washing stations were effective (92%) than those that were mobile (69%), a greater proportion of permanent stations were effective (91%) than those that were temporary (73%), and a great proportion of stations that had piped water for their water supply were effective (100%) than those that had individual storage tanks (85%). However, sample sizes across these strata were small as a substantial proportion of studies did not report on station mobility (46%), permanency (44%), water supply (61%), or material (85%), nor did they indicate the specific location of the station in its setting (86%).

#### 3.3.e. (Question E) Among interventions to improve hand hygiene in community settings, what hand hygiene station design adaptations (e.g., placement, nudges, and cues) have been effective at improving and sustaining hand hygiene?

To understand which adaptations were effective at improving hand hygiene, we extracted the adaptation types and effectiveness information from interventions that included hand hygiene stations. Ten studies evaluated hand hygiene station design adaptations, six (40%) of which performed better than the standard design (S14 – HH station design adaptations). Five studies evaluated the use of ‘nudges’ (e.g., painted or paved footpaths) and only three (60%) performed better than the standard design; the other two performed the same. Successful ‘nudge’ adaptations used painted footpaths [39,40] or arrows [41]. Adaptations that performed the same as the standard design employed similar nudges; both had paved footpaths and painted handwashing stations, and one also had painted shoeprints. Three evaluated cues to action (e.g., posters, automatic towel availability); two performed better and one performed worse. Successful ‘cue to action’ adaptations included providing a poster indicating higher prevalence (i.e., from one out of five to four out of five) of hand hygiene practices among the target user [42] or having towels that are automatically available without user action compared to motion-activated dispensers [43]. The ‘cue to action adaptation that performed worse was a poster warning about flu transmission, compared to the standard design which offered handwashing instructions [44]. Two studies evaluated changes to placement; placement in front of a nature-style background performed better than the standard in front of a simple background [45], and placement by a mirror performed worse [46].

#### 3.3.f. (Question F) Among interventions to improve hand hygiene in community settings, what level of frequency and intensity of behavior change interventions is necessary to effectively improve hand hygiene?

To assess which levels of frequency and intensity of behavior change interventions is effective at improving hand hygiene, we extracted data from interventions that were identified to vary in frequency or intensity. Six studies evaluated adaptations in the frequency (n=1) or intensity (n=5) of behavior change interventions, three of which—all assessing higher intensity—performed better than the standard design (S15 – Frequency and intensity). Successful adaptations included providing hygiene promotion activities (videos, Glo Germ, demonstrations, posters) with handwashing stations compared to providing handwashing stations alone [47]; providing an enhanced community-led total sanitation (CLTS) approach with additional discussions, skit, films, pledges, stickers, and report cards compared to CLTS alone [48]; and providing household visits from social workers and community health volunteers with trainings compared to trainings alone [49].

#### 3.3.g. (Question G) Among interventions to improve hand hygiene in community settings, how do hand hygiene practices vary by population groups, risk scenarios or over time?

To answer this research question, we extracted data on population groups, risk scenarios, as well as the number of time points reported in each study. Among all hand hygiene interventions studies, we report if and how handwashing practices are reported to vary by population groups, risk scenarios, and over time (S16 – Pop groups, risk, time). Among the 223 studies, 240 population groups were engaged, with some studies targeting multiple groups. The largest proportion of studies focused on children (83; 35%), of which 69 (83%) were effective.

Ten distinct vulnerable groups were identified across 34 studies: pregnant women (13; 38%), individuals with specific illnesses or risk factors (6; 18%), persons with disabilities (4; 12%), people of low income (2; 6%), children who are orphans or in foster care (2; 6%), people who are refugees or displaced (2; 6%), people belonging to specific ethnic or religious groups (2; 6%), people who inject drugs (1; 3%), newborns (1; 3%), and people who are immigrants or migrants (1; 3%). Sixty-two percent (8/13) of studies that focused on pregnant women reported interventions to be effective, and 83% (5/6) of studies that focused on individuals with specific illnesses or risk factors reported interventions to be effective.

Six risk scenarios were explored across studies, most focused on infectious disease: Flu (8; 47%), COVID-19 (4; 24%), internal displacement (2; 12%), hurricane (1; 6%), earthquake (1; 6%), and Ebola (1; 6%). Seventy-five percent (6/8) of studies that focused on Flu reported interventions to be effective, and 75% (3/4) of studies that focused on COVID-19 reported interventions to be effective.

Across studies, a minority collected data post-intervention at two or more time points: two time points (30); three (13); four (3), five (2), six (2). Seventy-nine percent (24/30) of studies that collected data two times post-intervention reported interventions to be effective, and 100% (13/13) of studies that collected data three times post-intervention reported interventions to be effective. Full citations of all included studies are in the supplement (S17 – Full ref list).

### 3.4 Patient Involvement Statement

Patients or the public were not involved directly in the design, or conduct, or reporting, or dissemination plans of our research. This evidence synthesis supports the forthcoming WHO Guidelines for Hand Hygiene in Community Settings; the study questions were developed in broad consultation with a network of key partners. Findings from this review will be disseminated alongside the Guidelines.

## 4. DISCUSSION

Our review of hand hygiene interventions in community settings found that a large proportion (82%) of interventions were effective at improving hand hygiene. Most interventions took place in households or schools; there is limited research set in workplaces and public settings in general (e.g., markets). Below, we discuss considerations for each of the focal research questions in turn. Notably: (a) theory was not extensively used in intervention design; the proportion of interventions that used theory and were effective was no greater than those that did not. (b) A large proportion of interventions addressed ‘Action Knowledge’, or how to wash hands, despite this factor not being identified within our sample or in other studies as a notable barrier or enabler of hand hygiene. Further, interventions did not always address identified barriers or enablers, potentially missing critical opportunities to drive behavior. (c) There was substantial heterogeneity in the BCTs used across the interventions; further research would benefit from intentional design and selection of specific BCTs to evaluate. (d/e) The handwashing station designs used in this study were mostly effective at improving hand hygiene; only 10 studies assessed design adaptations, of which six outperformed the standard design. (f) Too few studies (6) assessed changes in frequency or intensity of behavior change interventions to draw firm conclusions. (g) Interventions that included or focused on people with disabilities were scarce. We identified inconsistencies in intervention reporting and thus conclude by presenting recommendations for intervention reporting to facilitate future learning and synthesis. Though we are not able to identify clear recommendations for each of our research questions, most of the interventions within this review were found to be effective, demonstrating that there are multiple ways to improve hand hygiene. For policymakers seeking to understand how to improve hand hygiene within their context, the library of studies included in this systematic review can be used to inform local efforts.

### Use of theories or models to inform intervention design

Consistent with other research, theory was not used extensively to inform intervention design, nor did theory use result in difference in intervention effectiveness [50]. We did not find evidence that the use of theory improved intervention effectiveness. 28% (n=63) of included studies utilized a theory or model for designing a hand hygiene intervention; 83% of those that used theory were effective compared to 82% that did not. In a 2014 meta-analysis, Prestwich and colleagues assessed physical activity and healthy eating interventions to discern the extent to which theory influenced the effectiveness of health behavior interventions. They found a marginally greater proportion of studies leveraged theory (56%) and only a weak relationship between the theory used, and the extent of theory use, with intervention effectiveness.

They also note that theory was not used extensively in the intervention design process [50]. However, we suggest that further research is needed before ruling out theory use in the design of hand hygiene interventions. Incorporating a theory of change into intervention design could ensure that the intervention is effective. Using a systematic approach to document and report on the use of theory of change would ensure that future research is able to assess the relationship between theory application and intervention effectiveness [51]. Assessing this relationship was beyond the scope of this review, but future research understanding these assessments would provide useful information for intervention design.

### Effective leveraging of identified barriers and enablers

Within the review, we identified a lack of alignment between what studies reported to be barriers and/or enablers and what interventions actually addressed, suggesting that studies need to improve reporting of known barriers and enablers or better leverage evidence in intervention design. It is widely acknowledged that understanding the factors (e.g., the barriers and enablers) that influence health behaviors is essential for designing interventions that can bring about change [36,37,52–54]. But it is not clear that studies optimally leveraged factors that influenced behavior. First, to be specific, we found studies to report barriers and enablers but not leverage them. Notably, ‘hand hygiene station design’ —a Physical Opportunity factor—was reported to be an enabler in ten studies and a barrier in two but was addressed in only three. Second, many of the factors addressed by interventions in our review were not reported within the studies to be barriers or enablers in need of addressing. Specifically, ‘Action Knowledge’ was reported to be a barrier or enabler in 38 (17%) studies yet was addressed in 190 (85%). In one study, for example, the intervention utilized multimedia messages and visits from community health workers to improve knowledge of hand hygiene behaviors, but the authors did not report this Psychological Capability (‘Action Knowledge’) factor to be an enabler or a barrier [55]. Additionally, no studies reported hand hygiene station locations to be a barrier or an enabler, yet 34 (15%) interventions addressed hand hygiene station location. Taken together, our findings suggest the need for hand hygiene intervention studies to more extensively justify why specific barriers or enablers are or are not addressed. This justification is particularly necessary when known behavioral barriers and enablers are not addressed to ensure that decisions were deliberate (e.g., addressing the factor is too expensive or impractical). Further, it is also crucial to justify why interventions address factors that have not been reported to influence the behavior a priori. Addressing factors not identified as barriers or enablers may unnecessarily increase intervention costs. And while interventions may be effective when addressing factors that were not pre-identified, the potential additive benefit of doing so may be hard to determine or non-existent.

We also found there to be a mismatch between the barriers and enablers that were largely addressed among the interventions we reviewed in our study, and the barriers and enablers identified by other studies to be the greatest determinants of hand hygiene. As noted, in this review, we found that the majority (85%; 190/223) of interventions addressed ‘Action Knowledge’. In contrast, two recent systematic reviews that explored barriers and enablers to hand hygiene in community settings both found resources in the environment to be the most reported barriers or enablers [56,57]. The findings from these systematic reviews raise a critical if not obvious point — people cannot clean their hands if they do not have the resources to do so, regardless of how motivated they are or their level of knowledge about proper technique. Because access to these resources is fundamental, interventions and programs need to either provide access to adequate resources or ensure they are already in place if hand hygiene is expected to improve or be sustained. Otherwise, efforts to motivate hand hygiene or instill knowledge will not be effective, could waste valuable resources, and may even be unethical if people are subject to messaging yet lack the ability to act. Moreover, the resources provided should suit the needs of a given population, enabling hand washing to be as easy, convenient and quick as possible, particularly given the time and responsibility constraints that have also been reported [56–63]. Indeed, across the public health sector, interventions that change the environmental context are the most recommended because they have been shown to ‘require less individual effort and have the greatest population impact’ [64]. Hand hygiene efforts should take heed to ensure the environmental context is sufficient and optimized to enable hand hygiene.

Within this review, we are not able to identify specific barriers or enablers that interventions must address in order to be effective at improving hand hygiene. Most of the studies within our review did not report which barriers and enablers were impacting hand hygiene practice as they were primarily quantitative and were not all drawing on previous qualitative or formative research. Though this review cannot recommend specific barriers or enablers to leverage, a recent review synthesizes qualitative studies to understand the most common barriers and enablers that impact hand hygiene practice [57].

### Implementation of behavior change techniques

The interventions in this review used a range of BCTs across multiple package types and settings, limiting identification of which specific BCTs are most effective in improving hand hygiene. Four BCT intervention package types addressing various combinations of BCTs represent the majority of the intervention packages identified in our review. These combinations of intervention packages primarily were associated with three intervention functions (education, training, and environmental restructuring). However, it was unclear from these analyses what the added value of each additional BCT component had in improving hand hygiene. The substantial variation in the types and number of BCTs used varied between packages. Other systematic reviews have also found substantial heterogeneity in the behavior change techniques used in interventions. In assessing behavior change techniques used in physical activity interventions, the authors found that there was substantial variation in the number of BCTs used per intervention but they did not find that any specific combinations of BCTs that were effective [65]. Other systematic reviews have highlighted the difficulty in evaluating effectiveness of BCTs and have recommended more intentional design of BCT evaluations to assess which ones are effective [66], going as far as proposing an ontology of behavior change interventions so that there is a standard terminology and classification system that can be used to describe and design behavior change interventions [67]. Those working in hand hygiene could consider leveraging the findings herein as a sector-specific taxonomy of hand hygiene intervention types to inform and identify future interventions. Moreover, to better understand which BCTs are effective at improving hand hygiene, interventions need to be deliberately and thoughtfully designed, with consideration to which BCTs are most needed in a specific context. The BCTs that are chosen should relate back to and address barriers and enablers that are relevant in a specific context. This would also require better delineation and understanding of contextual considerations that drive hygiene interventions.

### Hand hygiene station designs and associated adaptations

Consistent with other reviews, implementing hand hygiene stations were found to be effective in improving hand hygiene practice [20]. Among the 223 studies reviewed, 21% (n=46) reported using a hand hygiene station in their intervention design. Data from a 2020 review on the use of tippy-taps on hand hygiene revealed there were increases in hand hygiene practice after the tippy-tap had been introduced among the studies that measured hand hygiene practice [20]. In this review, 80% of studies that used hand hygiene stations were effective in improving hand hygiene outcomes. The most common design being a raised bucket with tap/outlet, showing a 75% effectiveness rate. The effectiveness varied based on station features such as mobility, permanency, and water supply, with fixed, permanent stations with piped water supply showing higher effectiveness rates. However, the analysis of design features and hand hygiene station types was limited by incomplete reporting on station characteristics and settings across many studies, further underscoring the need for more comprehensive data and reporting.

Among the limited sample, hand hygiene adaptations were effective, though more research is warranted. Of the 46 studies that implemented a hand hygiene station, only ten studies examined adaptations in hand hygiene station design. Of the six studies that outperformed the original design, successful adaptations included aesthetic improvements [45], nudges such as painted footpaths and arrows [39–41], and cues to action, such as posters indicating higher hand hygiene prevalence or readily available towels [42,43]. Other research has highlighted the importance of design adaptations in improving acceptability and adoption of the hand hygiene station. In a qualitative assessment of hand hygiene stations in displacement camps, participants reported that there would be greater acceptability of the hand hygiene station if the station had been redesigned to be more aesthetically pleasing and functional [68]. Since only ten studies evaluated adaptations in hand hygiene station design within this review, additional research on design adaptations is needed to understand how adaptations may improve effectiveness of hand hygiene station interventions.

### Variations of hand hygiene practice across vulnerable groups, risk scenarios, and over time

While our review did identify studies examining hand hygiene across various vulnerable groups and different risk scenarios, there were few studies that examined a key vulnerable group, people with disabilities. Only four studies included people with disabilities as their primary focal population. Two studies evaluated hand hygiene intervention adaptations for intellectually disabled children and adults [69,70], one adapted an education hygiene program for hearing-impaired children [71], and one did not specify the types of disability included in their focal population nor did they include how they adapted their hand hygiene intervention [72]. People with disabilities face additional barriers in being able to access hand hygiene. Recent qualitative research understanding the experiences of people with various disabilities in Tanzania highlighted how hand hygiene was inaccessible [73]. Some people with physical disabilities were not able to access water needed for handwashing or were not able to use the handwashing station. For participants who had visual disabilities, educational information provided through leaflets or brochures were inaccessible. For hand hygiene interventions to be truly accessible for everyone, additional research is needed to understand how these interventions should be adapted for all types and levels of disability.

### Need for improved, systematic reporting of hand hygiene interventions

A common limitation across the included literature was inconsistent reporting of hand hygiene interventions in the included studies. A recent scoping review of the 40 most cited evaluations of WASH interventions published in the last 10 years (2012–2022) found that inconsistent reporting of WASH implementation illustrates a major challenge in the sector [74]. Although our review revealed a wealth of knowledge on effective theories, barriers and enablers, and behavior change techniques utilized in hand hygiene interventions, our findings were constrained by the quality of intervention reporting in the included studies. Notably, a quarter of the studies in our review failed to clearly specify the hand hygiene practices targeted for improvement or defined the focal population. Further, this review did not yield sufficient information to understand the level of frequency and intensity of behavior change interventions needed to effectively enhance hand hygiene in community settings (sub-question f); only six studies examined how changing frequency or intensity of hand hygiene interventions impacted hand hygiene outcomes. As described above, inconsistent reporting also affected our ability to learn about effective hand hygiene station design features. Study reporting should include detailed description of the intervention and reporting guidelines specific to hand hygiene are available [75].

### Strengths and Limitations

This review was part of an integrated protocol for multiple related reviews which included an exhaustive search strategy encompassing multiple databases and grey literature sources and a two-phased approach to identify relevant literature of hand hygiene in community settings. This is the first review to comprehensively examine interventions designed to improve hand hygiene in community settings. A key limitation of this effort, however, was the need to impose a structure for categorizing and presenting interventions that was different than what was used by the studies themselves. Specifically, we leveraged the COM-B model and accompanying behavior change wheel [31], which we found to be used by only 6 (3%) of included studies. To use COM-B and the associated resources, we needed to classify reported barriers and aspects of the interventions ourselves. Our ability to make these classifications was limited by the information available in the studies, potentially limiting our ability to make classification decisions that would align with what the original study authors might have done. Despite the potential challenges with imposing an external framework, the use of COM-B is also a strength of this review. As the hand hygiene interventions included in the study are quite varied, using COM-B provided a means to identify and group like components to determine what basic elements were comparable and most leveraged.

## 5. CONCLUSION

We found a large proportion (82%) of interventions to be effective. However, our ability to compare interventions was limited due to variability in research outcomes, settings, and intervention components, functions and behavior change techniques leveraged, preventing us from making specific recommendations to use or not use specific intervention types in community settings. That said, findings suggest critical ways that future interventions can be strengthened. Specifically, interventions can better leverage barriers and enablers of hand hygiene practices. Decisions to include interventions components should be aligned with known barriers and enablers. Failing to address known barriers and enablers can limit or prevent impact, and adding additional components that are not needed may not be resource (e.g., cost, time) effective. Finally, interventions need to consider if resources in the physical environment are available for those engaged to wash hands, like water, soap, and hygiene stations. As learned from intervention research in the broader public health field, there will be greater likelihood of impact if these aspects of the environment do exist. If settings do not already have these critical hand hygiene components in the environment, interventions that seek to improve hand hygiene only through motivation, social pressure, or by increasing knowledge should be reconsidered.

## Data Availability

All data produced will be made available online at publication via FigShare.

## Authors’ contributions

OC, JEM, and BG conceived the review and designed the specific research questions. BAC and MW are the guarantors of the review. BAC and SKP conceived and designed the specific analysis strategy described herein. SKP led design of the data extraction process with BAC, JSS, and LAO support. HKR led the literature search process. SKP and EL conducted screenings. SKP and EL conducted the data extraction. SKP led the analysis, led manuscript writing, and oversaw revisions. BAC and JSS drafted portions of the manuscript. All authors read and approved the final version of the manuscript.

## Acknowledgements

The authors would like to extend their gratitude to Prof. Susan Michie for reviewing and providing edits to the codebook for classifying enablers and barriers according to the COM-B model. We acknowledge the team of screeners who contributed substantially to the review process as part of phase 1 and phase 2 screening (Erika Canda, Lanelle Nwogalanya, Jordan Honeycutt, Michael Horner-Ibler, Rosemary Madaki, Kennedy Files, Nick An, Shahreen Hussain, Norah McKinley, Josef Zhao, Kainalu Bailey) and Dewan Muhammad Shoaib for work on the bar chart.

