## Supplementary material for "Interventions to improve hand hygiene in community settings: A systematic review of theories, barriers and enablers, behavior change techniques, and hand hygiene station design features": S2

**S2** – PRISMA flow chart

**S2** – PRISMA flow chart

***Supplementary File 2:*** **PRISMA flow chart.**

We identified 260 reports of eligible studies, which represented 223 studies that met inclusion criteria once duplicate mention of interventions in different studies were grouped.

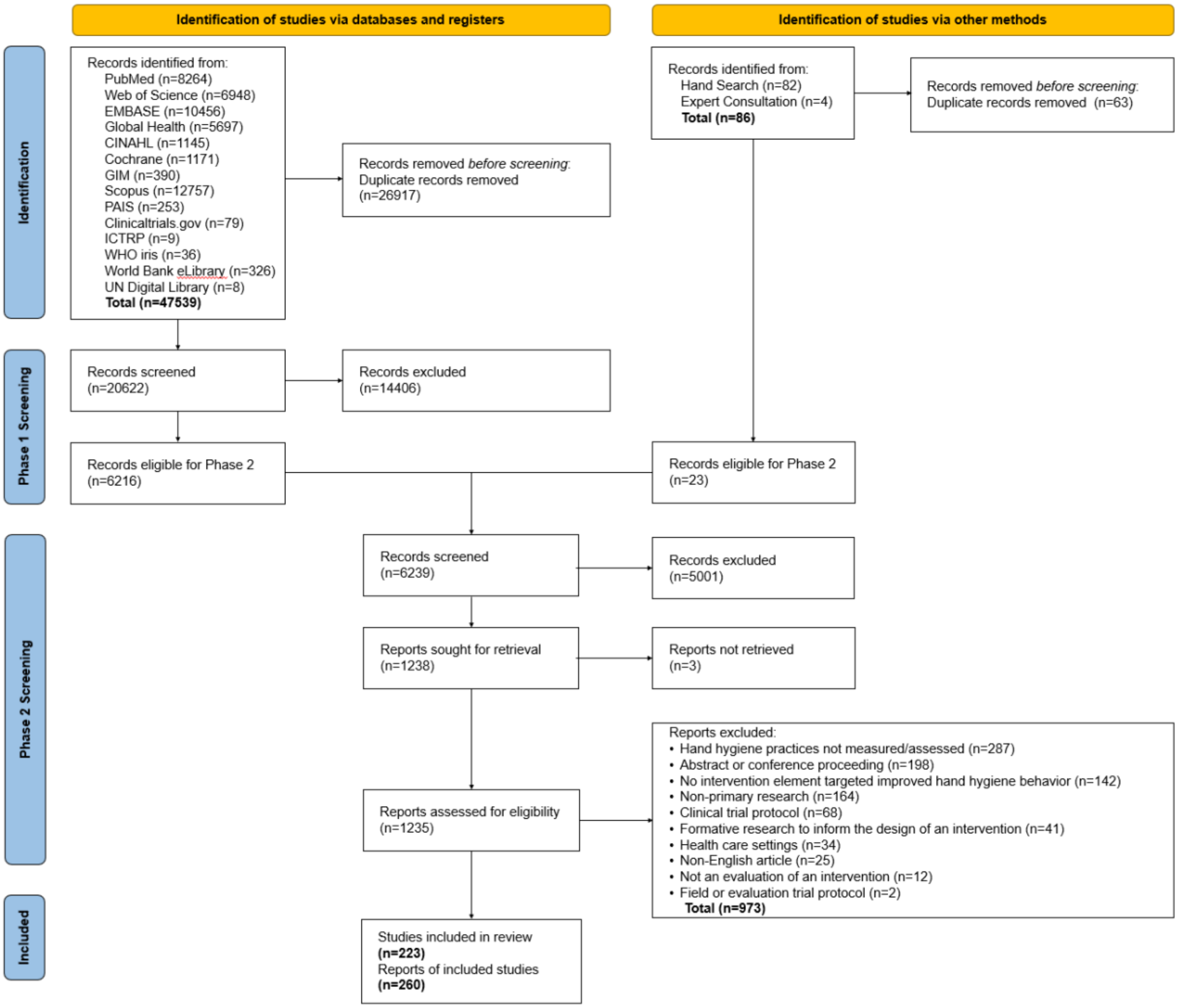
