## Supplementary material for "Interventions to improve hand hygiene in community settings: A systematic review of theories, barriers and enablers, behavior change techniques, and hand hygiene station design features": S3

**S3 –** Covidence extraction sheet

### Extraction fields for RQ3.2

| # | Field | Details | Entry | Source |
|---|---|---|---|---|
| **1. Information about the extraction** | | | | |
| 1.1 | Reviewer ID | First and last initial (e.g., JS) | Text | |
| 1.2 | Date of extraction | mm/dd/yy | Text | |
| **2. Information about the study** | | | | |
| 2.1 | Author | Family name of first author | Text<br>999 = Not applicable | |
| 2.2 | Publication year | | Text<br>999 = Not applicable | |
| 2.3 | Title | Title of study that data are extracted from | Text | |
| 2.4 | Link | Paste the DOI or hyperlink to publication | Text<br>999 = Not applicable | |
| 2.5 | Study type | | *Select one*<br>(1) Journal article<br>(2) Grey literature (e.g., unpublished academic papers [e.g., thesis], non-peer reviewed papers, research and committee reports, government reports, conference papers/abstract, ongoing research)<br>(777) Other - specify | |
| 2.6 | Study designc | | *Select one*<br>(1) Descriptive (survey)<br>(2) Descriptive (qualitative) | |

| | | | | |
|---|---|---|---|---|
| | | | (3) Descriptive (mixed methods)<br>(4) Randomized controlled trial<br>(5) Non-randomized control trial<br>(6) Quasi-experimental<br>(7) Case-control<br>(8) Cross-sectional<br>(9) Cohort<br>(10) Non-primary research<br>(777) Other – specify | |
| 2.7 | Registered trial | Does the study report that it is linked to a registered trial? (e.g., clinicaltrials.gov, ICTRP) | *Select one*<br>(1) Yes<br>(0) No | |
| 2.7.1 | Trial # | If linked to a registered trial, paste trial registration number | Text<br>(999) Not applicable | |
| **3. Eligibility** | | | | |
| 3.1 | Confirm eligibility | Confirm eligible based on RQ criteria in PICO(S) or SPIDER format<br><br>DO NOT PROCEED IF ALL ARE NOT CHECKED | *Check multiple*<br>• Sample - The study includes general populations in community settings<br><br>• PoI/Intervention - The study includes the correct PoIs (a-e) and/or interventions (f-g) of: (a) Behavior change theories among interventions to improve hand hygiene in community settings; (b) Effective leveraging of identified barriers and | Phase 2 criteria |

| | | | |
|---|---|---|---|
| | | | enablers of hand hygiene among interventions to improve hand hygiene in community settings; (c) Behavior change techniques to promote handwashing among interventions to improve hand hygiene in community settings; (d) Hand hygiene station design among interventions to improve hand hygiene in community settings; (e) Hand hygiene practices among key population groups and risk scenarios in community settings; (f) Design adaptations (e.g., placement, nudges, and cues) of hand hygiene stations; (g) Varying frequencies and intensities of behavior change interventions to promote effective hand hygiene<br><br>• Design - The study is an experimental or quasi-experimental study, randomized controlled trial, non-randomized |

| | | | | |
|---|---|---|---|---|
| | | | control trial, or before-after study<br><br>• Evaluation - The study evaluates effective (and sustained) hand hygiene or variations in hand hygiene practices<br><br>• Research type - The study uses qualitative, quantitative, and/or mixed methods | |
| **4. Setting** | | | | |
| 4.1 | Country | Which country is represented in the study? (List all countries separated by a comma, if study is from multiple sites) | Text<br>(999) Not applicable | |
| 4.2 | Region | Which region is represented in the study? | *Check multiple*<br>(1) Africa<br>(2) Asia<br>(3) Europe<br>(4) Latin America/Caribbean<br>(5) Middle East<br>(6) North America<br>(8) Oceania<br>(9) Multiple Regions<br>(10) Unspecified<br>(999) Not applicable | Women's Empowerment WASH SR |
| 4.3 | Urban/Rural | Does the setting of the population fall under any of these specific categories? Select all that apply | *Check multiple*<br>(1) Urban<br>(2) Rural<br>(3) Peri-urban | |

| | | | | |
|---|---|---|---|---|
| | | | (777) Other - specify<br>(888) Not reported<br>(999) Not applicable | |
| 4.4 | Community setting | Does the setting of the population fall under any of these specific categories? Select all that apply | *Check multiple*<br>(1) Domestic - Households<br>(2) Public - Markets<br>(3) Public - Public transportation hubs<br>(4) Public - Parks, squares, or other public outdoor spaces,<br>(5) Institutions - Workplace<br>(6) Institutions - Schools<br>(7) Institutions - Universities<br>(8) Institutions - Places of worship<br>(9) Institutions - Prisons and places of detention<br>(10) Internally displaced people camps<br>(777) Other - specify<br>(888) Not reported<br>(999) Not applicable | Macleod et al |
| **5. Methods** | | | | |
| 5.1 | Aim of study | Paste the aim/objective/purpose/goal as stated in the study | Text<br>(888) Not reported<br>(999) Not applicable | |
| 5.2 | Primary study outcome | What was the primary outcome for this study? | *Select one*<br>(1) Hand hygiene<br>(2) Diarrheal diseases<br>(3) Respiratory infections<br>(4) Influenza<br>(5) Infectious diseases<br>(6) Nutrition<br>(7) Mental/social well-being | |

| | | | |
|---|---|---|---|
| | | | (8) Neglected tropical diseases<br>(9) School absenteeism<br>(10) COVID-19<br>(11) Food hygiene<br>(777) Other – specify<br>(999) Not applicable |
| 5.3 | Start date | What is the study start date?<br>Month, Year | Text<br>(888) Not reported |
| 5.4 | End date | What is the study end date?<br>Month, Year | Text<br>(888) Not reported |
| **6. Participants** | | | |
| 6.1 | Study participants | What is the group of people that researchers are examining in the study? Select all that apply | *Check multiple*<br>(1) General population<br>(2) Adults (Women and Men)<br>(3) Adults (Women only)<br>(4) Adults (Men only)<br>(5) Children (Girls and Boys)<br>(6) Children (Girls only)<br>(7) Children (Boys only)<br>(8) Mother-child dyads<br>(9) Food workers<br>(10) Non-food occupational workers<br>(777) Other – specify<br>(888) Not reported<br>(999) Not applicable |
| 6.2 | Vulnerable populations | Does study examine any of the following vulnerable populations? Select all that apply | *Check multiple*<br>(1) Individuals with specific illness or risk factors<br>(2) Specific ethnic or religious groups |

| | | | |
|---|---|---|---|
| | | | (3) Persons experiencing homelessness<br>(4) Persons with disabilities<br>(5) Immigrants and migrants<br>(6) Refugees and displaced persons<br>(7) Elderly<br>(8) Pregnant women<br>(777) Other - specify<br>(888) Not reported<br>(999) Not applicable |
| 6.3 | Number of participants | What is the total number of participants/sample size? | Text<br>(888) Not reported<br>(999) Not applicable |
| 6.3.1 | Sample size – Intervention arm 1 | What is the sample size for intervention arm/group 1? | Text<br>(888) Not reported<br>(999) Not applicable |
| 6.3.2 | Sample size – Intervention arm 2 | What is the sample size for intervention arm/group 2? | Text<br>(888) Not reported<br>(999) Not applicable |
| 6.3.3 | Sample size – Control arm | What is the sample size for control arm/group? | Text<br>(888) Not reported<br>(999) Not applicable |
| 6.4 | Age group | What is the age range in the eligibility criteria for participants? | Text<br>(888) Not reported |
| 6.5 | Sex | What is the sex of the primary research population? | *Select one*<br>(1) Male<br>(2) Female<br>(3) Both male and female<br>(4) Unspecified |
| **7. Intervention**<br>This section also addresses:<br>**RQ3.2b Among interventions to improve hand hygiene in community settings, which have effectively leveraged identified** | | | |

| | | | |
|---|---|---|---|
| **barriers and enablers of hand hygiene in community settings?**<br>**RQ3.2c Among interventions to improve hand hygiene in community settings, what behavior change techniques have been implemented to effectively improve and sustain handwashing practices?** | | | |
| 7.1 | Protocol | In the methods, is there reference to a published study protocol? | (0) No<br>(1) Yes |
| 7.1.1 | Protocol citation | If published protocol, paste full citation | Citation<br>(999) Not applicable |
| 7.2 | Intervention | Does the study report that it is linked to an intervention? | (0) No<br>(1) Yes |
| 7.2.1 | Intervention name | If linked to an intervention, paste the name of the intervention | Text<br>(888) Not reported<br>(999) Not applicable |
| 7.2.2 | Intervention overall effectiveness | Is the intervention effective at achieving the primary outcome? (Note: effectiveness is defined by the authors) | *Select one*<br>(0) No<br>(1) Yes |
| 7.3 | Intervention aim behaviors | Did the intervention target other behaviors in addition to handwashing? | *Select one*<br>(0) No<br>(1) Yes |
| 7.4 | Multi-arm interventions | Does the study have multiple intervention arms that targeted hand hygiene? | (0) No<br>(1) Yes |
| 7.5 | Soap provision | Did the study provide **soap**? | *Select one*<br>(0) No<br>(1) Yes<br>(999) Not applicable |
| 7.5.1 | Intervention text | For this component, please copy in the author's description. | Text<br>(888) Not reported<br>(999) Not applicable |
| 7.5.2 | Multi-arm | If the study is a multi-arm trial, which arm provided this intervention? | Text<br>(999) Not applicable |

| | | | |
|---|---|---|---|
| 7.5.3 | Explicit barrier | What barrier did the authors explicitly note that this intervention activity addressed? | Text<br>(888) Not noted<br>(999) Not applicable |
| 7.5.4 | Assumed barriers | If the barrier was not noted by the authors, what is the assumed barrier? | Text<br>(999) Not applicable |
| 7.5.5 | Explicit enabler | What enabler did the authors explicitly note that this intervention activity addressed? | Text<br>(888) Not noted<br>(999) Not applicable |
| 7.5.6 | Assumed enabler | If the enabler was not noted by the authors, what is the assumed enabler? | Text<br>(999) Not applicable |
| 7.5.7 | COM-B category | Which COM-B category was addressed through the intervention activity? | *Select multiple*<br>(1) Physical capability<br>(2) Psychological capability<br>(3) Physical opportunity<br>(4) Social opportunity<br>(5) Automatic motivation<br>(6) Reflective motivation<br>(999) Not applicable |
| 7.5.8 | Intervention function | Which COM-B intervention function maps to the behavior change technique above? | *Select multiple*<br>(1) Education<br>(2) Persuasion<br>(3) Incentivization<br>(4) Coercion<br>(5) Training<br>(6) Restriction<br>(7) Environmental Restructuring<br>(8) Modelling<br>(9) Enablement<br>(999) Not applicable |

| | | | |
|---|---|---|---|
| 7.5.9 | Behavior change technique – Goals and planning | Goals and planning: For this component, what behavior change techniques map to the intervention activity described above? | *Select multiple*<br>(1) Goal setting (behavior)<br>(2) Problem solving<br>(3) Goal setting (outcome)<br>(4) Action planning<br>(5) Review behavior goals<br>(6) Discrepancy between current behavior and goal<br>(7) Review outcome goals<br>(8) Behavioral contract<br>(9) Commitment<br>(999) Not applicable |
| 7.5.10 | Behavior change technique – Feedback and monitoring | Feedback and monitoring: For this component, what behavior change techniques map to the intervention activity described above? | *Select multiple*<br>(1) Monitoring of behavior by others without feedback<br>(2) Feedback on behavior<br>(3) Self-monitoring of behavior<br>(4) Self-monitoring of outcome(s) of behavior)<br>(5) Monitoring outcome(s) of behavior by others without feedback<br>(6) Biofeedback<br>(7) Feedback on outcome(s) of behavior<br>(999) Not applicable |
| 7.5.11 | Behavior change technique – Social support | Social support: For this component, what behavior change techniques map to the intervention activity described above? | *Select multiple*<br>(1) Social support (unspecified)<br>(2) Social support (practical)<br>(3) Social support (emotional)<br>(999) Not applicable |

| | | | |
|---|---|---|---|
| 7.5.12 | Behavior change technique – Shaping knowledge | Shaping knowledge: For this component, what behavior change techniques map to the intervention activity described above? | *Select multiple*<br>(1) Instruction on how to perform a behavior<br>(2) Information about antecedents<br>(3) Re-attribution<br>(4) Behavioral experiments<br>(999) Not applicable |
| 7.5.13 | Behavior change technique – Natural consequences | Natural consequences: For this component, what behavior change techniques map to the intervention activity described above? | *Select multiple*<br>(1) Information about health consequences<br>(2) Salience of consequences<br>(3) Information about social and environmental consequences<br>(4) Monitoring of emotional consequences<br>(5) Anticipated regret<br>(6) Information about emotional consequences<br>(999) Not applicable |
| 7.5.14 | Behavior change technique – Comparison of behavior | Comparison of behavior: For this component, what behavior change techniques map to the intervention activity described above? | *Select multiple*<br>(1) Demonstration of behavior<br>(2) Social comparison<br>(3) Information about others' approval<br>(999) Not applicable |
| 7.5.15 | Behavior change technique – Associations | Associations: For this component, what behavior change techniques map to the intervention activity described above? | *Select multiple*<br>(1) Prompts/cues<br>(2) Cue signaling reward<br>(3) Reduce prompts/cues<br>(4) Remove access to the reward<br>(5) Remove aversive stimulus |

| | | | |
|---|---|---|---|
| | | | (6) Satiation<br>(7) Exposure<br>(8) Associative learning<br>(999) Not applicable |
| 7.5.16 | Behavior change technique – Repetition and substitution | Repetition and substitution: For this component, what behavior change techniques map to the intervention activity described above? | *Select multiple*<br>(1) Behavioral practice/rehearsal<br>(2) Behavior substitution<br>(3) Habit formation<br>(4) Habit reversal<br>(5) Overcorrection<br>(6) Generalization of a target behavior<br>(7) Graded tasks<br>(999) Not applicable |
| 7.5.17 | Behavior change technique – Comparison of outcomes | Comparison of outcomes: For this component, what behavior change techniques map to the intervention activity described above? | *Select multiple*<br>(1) Credible source<br>(2) Pros and cons<br>(3) Comparative imagining of future outcomes<br>(999) Not applicable |
| 7.5.18 | Behavior change technique – Reward and threat | Reward and threat: For this component, what behavior change techniques map to the intervention activity described above? | *Select multiple*<br>(1) Material incentive (behavior)<br>(2) Material reward (behavior)<br>(3) Non-specific reward<br>(4) Social reward<br>(5) Social incentive<br>(6) Non-specific incentive<br>(7) Self-incentive<br>(8) Incentive (outcome)<br>(9) Self-reward<br>(10) Reward (outcome)<br>(11) Future punishment |

| | | | |
|---|---|---|---|
| | | | (999) Not applicable |
| 7.5.19 | Behavior change technique – Regulation | Regulation: For this component, what behavior change techniques map to the intervention activity described above? | *Select multiple*<br>(1) Pharmocological support<br>(2) Reduce negative emotions<br>(3) Conserving mental resources<br>(4) Paradoxical instructions<br>(999) Not applicable |
| 7.5.20 | Behavior change technique – Antecedents | Antecedents: For this component, what behavior change techniques map to the intervention activity described above? | *Select multiple*<br>(1) Restructuring the physical environment<br>(2) Restructuring the social environment<br>(3) Avoidance/reducing exposure to cues for the behavior<br>(4) Distraction<br>(5) Adding objects to the environment<br>(6) Body changes<br>(999) Not applicable |
| 7.5.21 | Behavior change technique – Identity | Identity: For this component, what behavior change techniques map to the intervention activity described above? | *Select multiple*<br>(1) Identification of self as role model<br>(2) Framing/reframing<br>(3) Incompatible beliefs<br>(4) Valued self-identity<br>(5) Identity associated with changed behavior<br>(999) Not applicable |
| 7.5.22 | Behavior change technique – Scheduled consequences | Scheduled consequences: For this component, what behavior change techniques map to the | *Select multiple*<br>(1) Behavior cost<br>(2) Punishment<br>(3) Remove reward |

| | | | |
|---|---|---|---|
| | | intervention activity described above? | (4) Reward approximation<br>(5) Rewarding completion<br>(6) Situation-specific reward<br>(7) Reward incompatible behavior<br>(8) Reward alternative behavior<br>(9) Reduce reward frequency<br>(10) Remove punishment<br>(999) Not applicable |
| 7.5.23 | Behavior change technique – Self-belief | Self-belief: For this component, what behavior change techniques map to the intervention activity described above? | *Select multiple*<br>(1) Verbal persuasion about capability<br>(2) Mental rehearsal of successful performance<br>(3) Focus on past success<br>(4) Self-talk<br>(999) Not applicable |
| 7.5.24 | Behavior change technique – Covert learning | Covert learning: For this component, what behavior change techniques map to the intervention activity described above? | *Select multiple*<br>(1) Imaginary punishment<br>(2) Imaginary reward<br>(3) Vicarious consequences<br>(999) Not applicable |
| 7.6 | Provision of alcohol rub/gel/hand sanitizer | Did the study provide **alcohol rub/gel/hand sanitizer?** | *Select one*<br>(0) No<br>(1) Yes<br>(999) Not applicable |
| 7.6.1 | Intervention text | For this component, please copy in the author's description. | Text<br>(888) Not reported<br>(999) Not applicable |
| 7.6.2 | Multi-arm | If the study is a multi-arm trial, which arm provided this intervention? | Text<br>(999) Not applicable |

| | | | |
|---|---|---|---|
| 7.6.3 | Explicit barrier | What barrier did the authors explicitly note that this intervention activity addressed? | Text<br>(888) Not noted<br>(999) Not applicable |
| 7.6.4 | Assumed barriers | If the barrier was not noted by the authors, what is the assumed barrier? | Text<br>(999) Not applicable |
| 7.6.5 | Explicit enabler | What enabler did the authors explicitly note that this intervention activity addressed? | Text<br>(888) Not noted<br>(999) Not applicable |
| 7.6.6 | Assumed enabler | If the enabler was not noted by the authors, what is the assumed enabler? | Text<br>(999) Not applicable |
| 7.6.7 | COM-B category | Which COM-B category was addressed through the intervention activity? | *Select multiple*<br>(1) Physical capability<br>(2) Psychological capability<br>(3) Physical opportunity<br>(4) Social opportunity<br>(5) Automatic motivation<br>(6) Reflective motivation<br>(999) Not applicable |
| 7.6.8 | Intervention function | Which COM-B intervention function maps to the behavior change technique above? | *Select multiple*<br>(1) Education<br>(2) Persuasion<br>(3) Incentivization<br>(4) Coercion<br>(5) Training<br>(6) Restriction<br>(7) Environmental Restructuring<br>(8) Modelling<br>(9) Enablement<br>(999) Not applicable |

| | | | |
|---|---|---|---|
| 7.6.9 | Behavior change technique – Goals and planning | Goals and planning: For this component, what behavior change techniques map to the intervention activity described above? | *Select multiple*<br>(1) Goal setting (behavior)<br>(2) Problem solving<br>(3) Goal setting (outcome)<br>(4) Action planning<br>(5) Review behavior goals<br>(6) Discrepancy between current behavior and goal<br>(7) Review outcome goals<br>(8) Behavioral contract<br>(9) Commitment<br>(999) Not applicable |
| 7.6.10 | Behavior change technique – Feedback and monitoring | Feedback and monitoring: For this component, what behavior change techniques map to the intervention activity described above? | *Select multiple*<br>(1) Monitoring of behavior by others without feedback<br>(2) Feedback on behavior<br>(3) Self-monitoring of behavior<br>(4) Self-monitoring of outcome(s) of behavior)<br>(5) Monitoring outcome(s) of behavior by others without feedback<br>(6) Biofeedback<br>(7) Feedback on outcome(s) of behavior<br>(999) Not applicable |
| 7.6.11 | Behavior change technique – Social support | Social support: For this component, what behavior change techniques map to the intervention activity described above? | *Select multiple*<br>(1) Social support (unspecified)<br>(2) Social support (practical)<br>(3) Social support (emotional)<br>(999) Not applicable |

| | | | |
|---|---|---|---|
| 7.6.12 | Behavior change technique – Shaping knowledge | Shaping knowledge: For this component, what behavior change techniques map to the intervention activity described above? | *Select multiple*<br>(1) Instruction on how to perform a behavior<br>(2) Information about antecedents<br>(3) Re-attribution<br>(4) Behavioral experiments<br>(999) Not applicable |
| 7.6.13 | Behavior change technique – Natural consequences | Natural consequences: For this component, what behavior change techniques map to the intervention activity described above? | *Select multiple*<br>(1) Information about health consequences<br>(2) Salience of consequences<br>(3) Information about social and environmental consequences<br>(4) Monitoring of emotional consequences<br>(5) Anticipated regret<br>(6) Information about emotional consequences<br>(999) Not applicable |
| 7.6.14 | Behavior change technique – Comparison of behavior | Comparison of behavior: For this component, what behavior change techniques map to the intervention activity described above? | *Select multiple*<br>(1) Demonstration of behavior<br>(2) Social comparison<br>(3) Information about others' approval<br>(999) Not applicable |
| 7.6.15 | Behavior change technique – Associations | Associations: For this component, what behavior change techniques map to the intervention activity described above? | *Select multiple*<br>(1) Prompts/cues<br>(2) Cue signaling reward<br>(3) Reduce prompts/cues<br>(4) Remove access to the reward<br>(5) Remove aversive stimulus |

| | | | |
|---|---|---|---|
| | | | (6) Satiation<br>(7) Exposure<br>(8) Associative learning<br>(999) Not applicable |
| 7.6.16 | Behavior change technique – Repetition and substitution | Repetition and substitution: For this component, what behavior change techniques map to the intervention activity described above? | *Select multiple*<br>(1) Behavioral practice/rehearsal<br>(2) Behavior substitution<br>(3) Habit formation<br>(4) Habit reversal<br>(5) Overcorrection<br>(6) Generalization of a target behavior<br>(7) Graded tasks<br>(999) Not applicable |
| 7.6.17 | Behavior change technique – Comparison of outcomes | Comparison of outcomes: For this component, what behavior change techniques map to the intervention activity described above? | *Select multiple*<br>(1) Credible source<br>(2) Pros and cons<br>(3) Comparative imagining of future outcomes<br>(999) Not applicable |
| 7.6.18 | Behavior change technique – Reward and threat | Reward and threat: For this component, what behavior change techniques map to the intervention activity described above? | *Select multiple*<br>(1) Material incentive (behavior)<br>(2) Material reward (behavior)<br>(3) Non-specific reward<br>(4) Social reward<br>(5) Social incentive<br>(6) Non-specific incentive<br>(7) Self-incentive<br>(8) Incentive (outcome)<br>(9) Self-reward<br>(10) Reward (outcome)<br>(11) Future punishment |

| | | | |
|---|---|---|---|
| | | | (999) Not applicable |
| 7.6.19 | Behavior change technique – Regulation | Regulation: For this component, what behavior change techniques map to the intervention activity described above? | *Select multiple*<br>(1) Pharmocological support<br>(2) Reduce negative emotions<br>(3) Conserving mental resources<br>(4) Paradoxical instructions<br>(999) Not applicable |
| 7.6.20 | Behavior change technique – Antecedents | Antecedents: For this component, what behavior change techniques map to the intervention activity described above? | *Select multiple*<br>(1) Restructuring the physical environment<br>(2) Restructuring the social environment<br>(3) Avoidance/reducing exposure to cues for the behavior<br>(4) Distraction<br>(5) Adding objects to the environment<br>(6) Body changes<br>(999) Not applicable |
| 7.6.21 | Behavior change technique – Identity | Identity: For this component, what behavior change techniques map to the intervention activity described above? | *Select multiple*<br>(1) Identification of self as role model<br>(2) Framing/reframing<br>(3) Incompatible beliefs<br>(4) Valued self-identity<br>(5) Identity associated with changed behavior<br>(999) Not applicable |
| 7.6.22 | Behavior change technique – Scheduled consequences | Scheduled consequences: For this component, what behavior change techniques map to the | *Select multiple*<br>(1) Behavior cost<br>(2) Punishment<br>(3) Remove reward |

| | | | |
|---|---|---|---|
| | | intervention activity described above? | (4) Reward approximation<br>(5) Rewarding completion<br>(6) Situation-specific reward<br>(7) Reward incompatible behavior<br>(8) Reward alternative behavior<br>(9) Reduce reward frequency<br>(10) Remove punishment<br>(999) Not applicable |
| 7.6.23 | Behavior change technique – Self-belief | Self-belief: For this component, what behavior change techniques map to the intervention activity described above? | *Select multiple*<br>(1) Verbal persuasion about capability<br>(2) Mental rehearsal of successful performance<br>(3) Focus on past success<br>(4) Self-talk<br>(999) Not applicable |
| 7.6.24 | Behavior change technique – Covert learning | Covert learning: For this component, what behavior change techniques map to the intervention activity described above? | *Select multiple*<br>(1) Imaginary punishment<br>(2) Imaginary reward<br>(3) Vicarious consequences<br>(999) Not applicable |
| 7.7 | Installation of handwashing station | Did the study provide/install **a handwashing station?** | *Select one*<br>(0) No<br>(1) Yes<br>(999) Not applicable |
| 7.7.1 | Intervention text | For this component, please copy in the author's description. | Text<br>(888) Not reported<br>(999) Not applicable |
| 7.7.2 | Multi-arm | If the study is a multi-arm trial, which arm provided this intervention? | Text<br>(999) Not applicable |

| | | | |
|---|---|---|---|
| 7.7.3 | Explicit barrier | What barrier did the authors explicitly note that this intervention activity addressed? | Text<br>(888) Not noted<br>(999) Not applicable |
| 7.7.4 | Assumed barriers | If the barrier was not noted by the authors, what is the assumed barrier? | Text<br>(999) Not applicable |
| 7.7.5 | Explicit enabler | What enabler did the authors explicitly note that this intervention activity addressed? | Text<br>(888) Not noted<br>(999) Not applicable |
| 7.7.6 | Assumed enabler | If the enabler was not noted by the authors, what is the assumed enabler? | Text<br>(999) Not applicable |
| 7.7.7 | COM-B category | Which COM-B category was addressed through the intervention activity? | *Select multiple*<br>(1) Physical capability<br>(2) Psychological capability<br>(3) Physical opportunity<br>(4) Social opportunity<br>(5) Automatic motivation<br>(6) Reflective motivation<br>(999) Not applicable |
| 7.7.8 | Intervention function | Which COM-B intervention function maps to the behavior change technique above? | *Select multiple*<br>(1) Education<br>(2) Persuasion<br>(3) Incentivization<br>(4) Coercion<br>(5) Training<br>(6) Restriction<br>(7) Environmental Restructuring<br>(8) Modelling<br>(9) Enablement<br>(999) Not applicable |

| | | | |
|---|---|---|---|
| 7.7.9 | Behavior change technique – Goals and planning | Goals and planning: For this component, what behavior change techniques map to the intervention activity described above? | *Select multiple*<br>(1) Goal setting (behavior)<br>(2) Problem solving<br>(3) Goal setting (outcome)<br>(4) Action planning<br>(5) Review behavior goals<br>(6) Discrepancy between current behavior and goal<br>(7) Review outcome goals<br>(8) Behavioral contract<br>(9) Commitment<br>(999) Not applicable |
| 7.7.10 | Behavior change technique – Feedback and monitoring | Feedback and monitoring: For this component, what behavior change techniques map to the intervention activity described above? | *Select multiple*<br>(1) Monitoring of behavior by others without feedback<br>(2) Feedback on behavior<br>(3) Self-monitoring of behavior<br>(4) Self-monitoring of outcome(s) of behavior)<br>(5) Monitoring outcome(s) of behavior by others without feedback<br>(6) Biofeedback<br>(7) Feedback on outcome(s) of behavior<br>(999) Not applicable |
| 7.7.11 | Behavior change technique – Social support | Social support: For this component, what behavior change techniques map to the intervention activity described above? | *Select multiple*<br>(1) Social support (unspecified)<br>(2) Social support (practical)<br>(3) Social support (emotional)<br>(999) Not applicable |

| | | | |
|---|---|---|---|
| 7.7.12 | Behavior change technique – Shaping knowledge | Shaping knowledge: For this component, what behavior change techniques map to the intervention activity described above? | *Select multiple*<br>(1) Instruction on how to perform a behavior<br>(2) Information about antecedents<br>(3) Re-attribution<br>(4) Behavioral experiments<br>(999) Not applicable |
| 7.7.13 | Behavior change technique – Natural consequences | Natural consequences: For this component, what behavior change techniques map to the intervention activity described above? | *Select multiple*<br>(1) Information about health consequences<br>(2) Salience of consequences<br>(3) Information about social and environmental consequences<br>(4) Monitoring of emotional consequences<br>(5) Anticipated regret<br>(6) Information about emotional consequences<br>(999) Not applicable |
| 7.7.14 | Behavior change technique – Comparison of behavior | Comparison of behavior: For this component, what behavior change techniques map to the intervention activity described above? | *Select multiple*<br>(1) Demonstration of behavior<br>(2) Social comparison<br>(3) Information about others' approval<br>(999) Not applicable |
| 7.7.15 | Behavior change technique – Associations | Associations: For this component, what behavior change techniques map to the intervention activity described above? | *Select multiple*<br>(1) Prompts/cues<br>(2) Cue signaling reward<br>(3) Reduce prompts/cues<br>(4) Remove access to the reward<br>(5) Remove aversive stimulus |

| | | | |
|---|---|---|---|
| | | | (6) Satiation<br>(7) Exposure<br>(8) Associative learning<br>(999) Not applicable |
| 7.7.16 | Behavior change technique – Repetition and substitution | Repetition and substitution: For this component, what behavior change techniques map to the intervention activity described above? | *Select multiple*<br>(1) Behavioral practice/rehearsal<br>(2) Behavior substitution<br>(3) Habit formation<br>(4) Habit reversal<br>(5) Overcorrection<br>(6) Generalization of a target behavior<br>(7) Graded tasks<br>(999) Not applicable |
| 7.7.17 | Behavior change technique – Comparison of outcomes | Comparison of outcomes: For this component, what behavior change techniques map to the intervention activity described above? | *Select multiple*<br>(1) Credible source<br>(2) Pros and cons<br>(3) Comparative imagining of future outcomes<br>(999) Not applicable |
| 7.7.18 | Behavior change technique – Reward and threat | Reward and threat: For this component, what behavior change techniques map to the intervention activity described above? | *Select multiple*<br>(1) Material incentive (behavior)<br>(2) Material reward (behavior)<br>(3) Non-specific reward<br>(4) Social reward<br>(5) Social incentive<br>(6) Non-specific incentive<br>(7) Self-incentive<br>(8) Incentive (outcome)<br>(9) Self-reward<br>(10) Reward (outcome)<br>(11) Future punishment |

| | | | |
|---|---|---|---|
| | | | (999) Not applicable |
| 7.7.19 | Behavior change technique – Regulation | Regulation: For this component, what behavior change techniques map to the intervention activity described above? | *Select multiple*<br>(1) Pharmocological support<br>(2) Reduce negative emotions<br>(3) Conserving mental resources<br>(4) Paradoxical instructions<br>(999) Not applicable |
| 7.7.20 | Behavior change technique – Antecedents | Antecedents: For this component, what behavior change techniques map to the intervention activity described above? | *Select multiple*<br>(1) Restructuring the physical environment<br>(2) Restructuring the social environment<br>(3) Avoidance/reducing exposure to cues for the behavior<br>(4) Distraction<br>(5) Adding objects to the environment<br>(6) Body changes<br>(999) Not applicable |
| 7.7.21 | Behavior change technique – Identity | Identity: For this component, what behavior change techniques map to the intervention activity described above? | *Select multiple*<br>(1) Identification of self as role model<br>(2) Framing/reframing<br>(3) Incompatible beliefs<br>(4) Valued self-identity<br>(5) Identity associated with changed behavior<br>(999) Not applicable |
| 7.7.22 | Behavior change technique – Scheduled consequences | Scheduled consequences: For this component, what behavior change techniques map to the | *Select multiple*<br>(1) Behavior cost<br>(2) Punishment<br>(3) Remove reward |

| | | | |
|---|---|---|---|
| | | intervention activity described above? | (4) Reward approximation<br>(5) Rewarding completion<br>(6) Situation-specific reward<br>(7) Reward incompatible behavior<br>(8) Reward alternative behavior<br>(9) Reduce reward frequency<br>(10) Remove punishment<br>(999) Not applicable |
| 7.7.23 | Behavior change technique – Self-belief | Self-belief: For this component, what behavior change techniques map to the intervention activity described above? | *Select multiple*<br>(1) Verbal persuasion about capability<br>(2) Mental rehearsal of successful performance<br>(3) Focus on past success<br>(4) Self-talk<br>(999) Not applicable |
| 7.7.24 | Behavior change technique – Covert learning | Covert learning: For this component, what behavior change techniques map to the intervention activity described above? | *Select multiple*<br>(1) Imaginary punishment<br>(2) Imaginary reward<br>(3) Vicarious consequences<br>(999) Not applicable |
| 7.8 | Education | Did the study implement an **education program on hand hygiene?** | *Select one*<br>(0) No<br>(1) Yes<br>(999) Not applicable |
| 7.8.1 | Intervention text | For this component, please copy in the author's description. | Text<br>(888) Not reported<br>(999) Not applicable |
| 7.8.2 | Multi-arm | If the study is a multi-arm trial, which arm provided this intervention? | Text<br>(999) Not applicable |

| | | | |
|---|---|---|---|
| 7.8.3 | Explicit barrier | What barrier did the authors explicitly note that this intervention activity addressed? | Text<br>(888) Not noted<br>(999) Not applicable |
| 7.8.4 | Assumed barriers | If the barrier was not noted by the authors, what is the assumed barrier? | Text<br>(999) Not applicable |
| 7.8.5 | Explicit enabler | What enabler did the authors explicitly note that this intervention activity addressed? | Text<br>(888) Not noted<br>(999) Not applicable |
| 7.8.6 | Assumed enabler | If the enabler was not noted by the authors, what is the assumed enabler? | Text<br>(999) Not applicable |
| 7.8.7 | COM-B category | Which COM-B category was addressed through the intervention activity? | *Select multiple*<br>(1) Physical capability<br>(2) Psychological capability<br>(3) Physical opportunity<br>(4) Social opportunity<br>(5) Automatic motivation<br>(6) Reflective motivation<br>(999) Not applicable |
| 7.8.8 | Intervention function | Which COM-B intervention function maps to the behavior change technique above? | *Select multiple*<br>(1) Education<br>(2) Persuasion<br>(3) Incentivization<br>(4) Coercion<br>(5) Training<br>(6) Restriction<br>(7) Environmental Restructuring<br>(8) Modelling<br>(9) Enablement<br>(999) Not applicable |

| | | | |
|---|---|---|---|
| 7.8.9 | Behavior change technique – Goals and planning | Goals and planning: For this component, what behavior change techniques map to the intervention activity described above? | *Select multiple*<br>(1) Goal setting (behavior)<br>(2) Problem solving<br>(3) Goal setting (outcome)<br>(4) Action planning<br>(5) Review behavior goals<br>(6) Discrepancy between current behavior and goal<br>(7) Review outcome goals<br>(8) Behavioral contract<br>(9) Commitment<br>(999) Not applicable |
| 7.8.10 | Behavior change technique – Feedback and monitoring | Feedback and monitoring: For this component, what behavior change techniques map to the intervention activity described above? | *Select multiple*<br>(1) Monitoring of behavior by others without feedback<br>(2) Feedback on behavior<br>(3) Self-monitoring of behavior<br>(4) Self-monitoring of outcome(s) of behavior)<br>(5) Monitoring outcome(s) of behavior by others without feedback<br>(6) Biofeedback<br>(7) Feedback on outcome(s) of behavior<br>(999) Not applicable |
| 7.8.11 | Behavior change technique – Social support | Social support: For this component, what behavior change techniques map to the intervention activity described above? | *Select multiple*<br>(1) Social support (unspecified)<br>(2) Social support (practical)<br>(3) Social support (emotional)<br>(999) Not applicable |

| | | | |
|---|---|---|---|
| 7.8.12 | Behavior change technique – Shaping knowledge | Shaping knowledge: For this component, what behavior change techniques map to the intervention activity described above? | *Select multiple*<br>(1) Instruction on how to perform a behavior<br>(2) Information about antecedents<br>(3) Re-attribution<br>(4) Behavioral experiments<br>(999) Not applicable |
| 7.8.13 | Behavior change technique – Natural consequences | Natural consequences: For this component, what behavior change techniques map to the intervention activity described above? | *Select multiple*<br>(1) Information about health consequences<br>(2) Salience of consequences<br>(3) Information about social and environmental consequences<br>(4) Monitoring of emotional consequences<br>(5) Anticipated regret<br>(6) Information about emotional consequences<br>(999) Not applicable |
| 7.8.14 | Behavior change technique – Comparison of behavior | Comparison of behavior: For this component, what behavior change techniques map to the intervention activity described above? | *Select multiple*<br>(1) Demonstration of behavior<br>(2) Social comparison<br>(3) Information about others' approval<br>(999) Not applicable |
| 7.8.15 | Behavior change technique – Associations | Associations: For this component, what behavior change techniques map to the intervention activity described above? | *Select multiple*<br>(1) Prompts/cues<br>(2) Cue signaling reward<br>(3) Reduce prompts/cues<br>(4) Remove access to the reward<br>(5) Remove aversive stimulus |

| | | | |
|---|---|---|---|
| | | | (6) Satiation<br>(7) Exposure<br>(8) Associative learning<br>(999) Not applicable |
| 7.8.16 | Behavior change technique – Repetition and substitution | Repetition and substitution: For this component, what behavior change techniques map to the intervention activity described above? | *Select multiple*<br>(1) Behavioral practice/rehearsal<br>(2) Behavior substitution<br>(3) Habit formation<br>(4) Habit reversal<br>(5) Overcorrection<br>(6) Generalization of a target behavior<br>(7) Graded tasks<br>(999) Not applicable |
| 7.8.17 | Behavior change technique – Comparison of outcomes | Comparison of outcomes: For this component, what behavior change techniques map to the intervention activity described above? | *Select multiple*<br>(1) Credible source<br>(2) Pros and cons<br>(3) Comparative imagining of future outcomes<br>(999) Not applicable |
| 7.8.18 | Behavior change technique – Reward and threat | Reward and threat: For this component, what behavior change techniques map to the intervention activity described above? | *Select multiple*<br>(1) Material incentive (behavior)<br>(2) Material reward (behavior)<br>(3) Non-specific reward<br>(4) Social reward<br>(5) Social incentive<br>(6) Non-specific incentive<br>(7) Self-incentive<br>(8) Incentive (outcome)<br>(9) Self-reward<br>(10) Reward (outcome)<br>(11) Future punishment |

| | | | |
|---|---|---|---|
| | | | (999) Not applicable |
| 7.8.19 | Behavior change technique – Regulation | Regulation: For this component, what behavior change techniques map to the intervention activity described above? | *Select multiple*<br>(1) Pharmocological support<br>(2) Reduce negative emotions<br>(3) Conserving mental resources<br>(4) Paradoxical instructions<br>(999) Not applicable |
| 7.8.20 | Behavior change technique – Antecedents | Antecedents: For this component, what behavior change techniques map to the intervention activity described above? | *Select multiple*<br>(1) Restructuring the physical environment<br>(2) Restructuring the social environment<br>(3) Avoidance/reducing exposure to cues for the behavior<br>(4) Distraction<br>(5) Adding objects to the environment<br>(6) Body changes<br>(999) Not applicable |
| 7.8.21 | Behavior change technique – Identity | Identity: For this component, what behavior change techniques map to the intervention activity described above? | *Select multiple*<br>(1) Identification of self as role model<br>(2) Framing/reframing<br>(3) Incompatible beliefs<br>(4) Valued self-identity<br>(5) Identity associated with changed behavior<br>(999) Not applicable |
| 7.8.22 | Behavior change technique – Scheduled consequences | Scheduled consequences: For this component, what behavior change techniques map to the | *Select multiple*<br>(1) Behavior cost<br>(2) Punishment<br>(3) Remove reward |

| | | | |
|---|---|---|---|
| | | intervention activity described above? | (4) Reward approximation<br>(5) Rewarding completion<br>(6) Situation-specific reward<br>(7) Reward incompatible behavior<br>(8) Reward alternative behavior<br>(9) Reduce reward frequency<br>(10) Remove punishment<br>(999) Not applicable |
| 7.8.23 | Behavior change technique – Self-belief | Self-belief: For this component, what behavior change techniques map to the intervention activity described above? | *Select multiple*<br>(1) Verbal persuasion about capability<br>(2) Mental rehearsal of successful performance<br>(3) Focus on past success<br>(4) Self-talk<br>(999) Not applicable |
| 7.8.24 | Behavior change technique – Covert learning | Covert learning: For this component, what behavior change techniques map to the intervention activity described above? | *Select multiple*<br>(1) Imaginary punishment<br>(2) Imaginary reward<br>(3) Vicarious consequences<br>(999) Not applicable |
| 7.9 | Training | Did the study implement a **training program on hand hygiene?** | *Select one*<br>(0) No<br>(1) Yes<br>(999) Not applicable |
| 7.9.1 | Intervention text | For this component, please copy in the author's description. | Text<br>(888) Not reported<br>(999) Not applicable |
| 7.9.2 | Multi-arm | If the study is a multi-arm trial, which arm provided this intervention? | Text<br>(999) Not applicable |

| | | | |
|---|---|---|---|
| 7.9.3 | Explicit barrier | What barrier did the authors explicitly note that this intervention activity addressed? | Text<br>(888) Not noted<br>(999) Not applicable |
| 7.9.4 | Assumed barriers | If the barrier was not noted by the authors, what is the assumed barrier? | Text<br>(999) Not applicable |
| 7.9.5 | Explicit enabler | What enabler did the authors explicitly note that this intervention activity addressed? | Text<br>(888) Not noted<br>(999) Not applicable |
| 7.9.6 | Assumed enabler | If the enabler was not noted by the authors, what is the assumed enabler? | Text<br>(999) Not applicable |
| 7.9.7 | COM-B category | Which COM-B category was addressed through the intervention activity? | *Select multiple*<br>(1) Physical capability<br>(2) Psychological capability<br>(3) Physical opportunity<br>(4) Social opportunity<br>(5) Automatic motivation<br>(6) Reflective motivation<br>(999) Not applicable |
| 7.9.8 | Intervention function | Which COM-B intervention function maps to the behavior change technique above? | *Select multiple*<br>(1) Education<br>(2) Persuasion<br>(3) Incentivization<br>(4) Coercion<br>(5) Training<br>(6) Restriction<br>(7) Environmental Restructuring<br>(8) Modelling<br>(9) Enablement<br>(999) Not applicable |

| | | | |
|---|---|---|---|
| 7.9.9 | Behavior change technique – Goals and planning | Goals and planning: For this component, what behavior change techniques map to the intervention activity described above? | *Select multiple*<br>(1) Goal setting (behavior)<br>(2) Problem solving<br>(3) Goal setting (outcome)<br>(4) Action planning<br>(5) Review behavior goals<br>(6) Discrepancy between current behavior and goal<br>(7) Review outcome goals<br>(8) Behavioral contract<br>(9) Commitment<br>(999) Not applicable |
| 7.9.10 | Behavior change technique – Feedback and monitoring | Feedback and monitoring: For this component, what behavior change techniques map to the intervention activity described above? | *Select multiple*<br>(1) Monitoring of behavior by others without feedback<br>(2) Feedback on behavior<br>(3) Self-monitoring of behavior<br>(4) Self-monitoring of outcome(s) of behavior)<br>(5) Monitoring outcome(s) of behavior by others without feedback<br>(6) Biofeedback<br>(7) Feedback on outcome(s) of behavior<br>(999) Not applicable |
| 7.9.11 | Behavior change technique – Social support | Social support: For this component, what behavior change techniques map to the intervention activity described above? | *Select multiple*<br>(1) Social support (unspecified)<br>(2) Social support (practical)<br>(3) Social support (emotional)<br>(999) Not applicable |

| | | | |
|---|---|---|---|
| 7.9.12 | Behavior change technique – Shaping knowledge | Shaping knowledge: For this component, what behavior change techniques map to the intervention activity described above? | *Select multiple*<br>(1) Instruction on how to perform a behavior<br>(2) Information about antecedents<br>(3) Re-attribution<br>(4) Behavioral experiments<br>(999) Not applicable |
| 7.9.13 | Behavior change technique – Natural consequences | Natural consequences: For this component, what behavior change techniques map to the intervention activity described above? | *Select multiple*<br>(1) Information about health consequences<br>(2) Salience of consequences<br>(3) Information about social and environmental consequences<br>(4) Monitoring of emotional consequences<br>(5) Anticipated regret<br>(6) Information about emotional consequences<br>(999) Not applicable |
| 7.9.14 | Behavior change technique – Comparison of behavior | Comparison of behavior: For this component, what behavior change techniques map to the intervention activity described above? | *Select multiple*<br>(1) Demonstration of behavior<br>(2) Social comparison<br>(3) Information about others' approval<br>(999) Not applicable |
| 7.9.15 | Behavior change technique – Associations | Associations: For this component, what behavior change techniques map to the intervention activity described above? | *Select multiple*<br>(1) Prompts/cues<br>(2) Cue signaling reward<br>(3) Reduce prompts/cues<br>(4) Remove access to the reward<br>(5) Remove aversive stimulus |

| | | | |
|---|---|---|---|
| | | | (6) Satiation<br>(7) Exposure<br>(8) Associative learning<br>(999) Not applicable |
| 7.9.16 | Behavior change technique – Repetition and substitution | Repetition and substitution: For this component, what behavior change techniques map to the intervention activity described above? | *Select multiple*<br>(1) Behavioral practice/rehearsal<br>(2) Behavior substitution<br>(3) Habit formation<br>(4) Habit reversal<br>(5) Overcorrection<br>(6) Generalization of a target behavior<br>(7) Graded tasks<br>(999) Not applicable |
| 7.9.17 | Behavior change technique – Comparison of outcomes | Comparison of outcomes: For this component, what behavior change techniques map to the intervention activity described above? | *Select multiple*<br>(1) Credible source<br>(2) Pros and cons<br>(3) Comparative imagining of future outcomes<br>(999) Not applicable |
| 7.9.18 | Behavior change technique – Reward and threat | Reward and threat: For this component, what behavior change techniques map to the intervention activity described above? | *Select multiple*<br>(1) Material incentive (behavior)<br>(2) Material reward (behavior)<br>(3) Non-specific reward<br>(4) Social reward<br>(5) Social incentive<br>(6) Non-specific incentive<br>(7) Self-incentive<br>(8) Incentive (outcome)<br>(9) Self-reward<br>(10) Reward (outcome)<br>(11) Future punishment |

| | | | |
|---|---|---|---|
| | | | (999) Not applicable |
| 7.9.19 | Behavior change technique – Regulation | Regulation: For this component, what behavior change techniques map to the intervention activity described above? | *Select multiple*<br>(1) Pharmocological support<br>(2) Reduce negative emotions<br>(3) Conserving mental resources<br>(4) Paradoxical instructions<br>(999) Not applicable |
| 7.9.20 | Behavior change technique – Antecedents | Antecedents: For this component, what behavior change techniques map to the intervention activity described above? | *Select multiple*<br>(1) Restructuring the physical environment<br>(2) Restructuring the social environment<br>(3) Avoidance/reducing exposure to cues for the behavior<br>(4) Distraction<br>(5) Adding objects to the environment<br>(6) Body changes<br>(999) Not applicable |
| 7.9.21 | Behavior change technique – Identity | Identity: For this component, what behavior change techniques map to the intervention activity described above? | *Select multiple*<br>(1) Identification of self as role model<br>(2) Framing/reframing<br>(3) Incompatible beliefs<br>(4) Valued self-identity<br>(5) Identity associated with changed behavior<br>(999) Not applicable |
| 7.9.22 | Behavior change technique – Scheduled consequences | Scheduled consequences: For this component, what behavior change techniques map to the | *Select multiple*<br>(1) Behavior cost<br>(2) Punishment<br>(3) Remove reward |

| | | | |
|---|---|---|---|
| | | intervention activity described above? | (4) Reward approximation<br>(5) Rewarding completion<br>(6) Situation-specific reward<br>(7) Reward incompatible behavior<br>(8) Reward alternative behavior<br>(9) Reduce reward frequency<br>(10) Remove punishment<br>(999) Not applicable |
| 7.9.23 | Behavior change technique – Self-belief | Self-belief: For this component, what behavior change techniques map to the intervention activity described above? | *Select multiple*<br>(1) Verbal persuasion about capability<br>(2) Mental rehearsal of successful performance<br>(3) Focus on past success<br>(4) Self-talk<br>(999) Not applicable |
| 7.9.24 | Behavior change technique – Covert learning | Covert learning: For this component, what behavior change techniques map to the intervention activity described above? | *Select multiple*<br>(1) Imaginary punishment<br>(2) Imaginary reward<br>(3) Vicarious consequences<br>(999) Not applicable |
| 7.10 | Multimedia messaging | Did the study use **multimedia messaging (posters, film/videos, pamphlets, etc.)** to promote hand hygiene? | *Select one*<br>(0)<br>(1) Yes<br>(999) Not applicable |
| 7.10.1 | Intervention text | For this component, please copy in the author's description. | Text<br>(888) Not reported<br>(999) Not applicable |
| 7.10.2 | Multi-arm | If the study is a multi-arm trial, which arm provided this intervention? | Text<br>(999) Not applicable |

| | | | |
|---|---|---|---|
| 7.10.3 | Explicit barrier | What barrier did the authors explicitly note that this intervention activity addressed? | Text<br>(888) Not noted<br>(999) Not applicable |
| 7.10.4 | Assumed barriers | If the barrier was not noted by the authors, what is the assumed barrier? | Text<br>(999) Not applicable |
| 7.10.5 | Explicit enabler | What enabler did the authors explicitly note that this intervention activity addressed? | Text<br>(888) Not noted<br>(999) Not applicable |
| 7.10.6 | Assumed enabler | If the enabler was not noted by the authors, what is the assumed enabler? | Text<br>(999) Not applicable |
| 7.10.7 | COM-B category | Which COM-B category was addressed through the intervention activity? | *Select multiple*<br>(1) Physical capability<br>(2) Psychological capability<br>(3) Physical opportunity<br>(4) Social opportunity<br>(5) Automatic motivation<br>(6) Reflective motivation<br>(999) Not applicable |
| 7.10.8 | Intervention function | Which COM-B intervention function maps to the behavior change technique above? | *Select multiple*<br>(1) Education<br>(2) Persuasion<br>(3) Incentivization<br>(4) Coercion<br>(5) Training<br>(6) Restriction<br>(7) Environmental Restructuring<br>(8) Modelling<br>(9) Enablement<br>(999) Not applicable |

| | | | |
|---|---|---|---|
| 7.10.9 | Behavior change technique – Goals and planning | Goals and planning: For this component, what behavior change techniques map to the intervention activity described above? | *Select multiple*<br>(1) Goal setting (behavior)<br>(2) Problem solving<br>(3) Goal setting (outcome)<br>(4) Action planning<br>(5) Review behavior goals<br>(6) Discrepancy between current behavior and goal<br>(7) Review outcome goals<br>(8) Behavioral contract<br>(9) Commitment<br>(999) Not applicable |
| 7.10.10 | Behavior change technique – Feedback and monitoring | Feedback and monitoring: For this component, what behavior change techniques map to the intervention activity described above? | *Select multiple*<br>(1) Monitoring of behavior by others without feedback<br>(2) Feedback on behavior<br>(3) Self-monitoring of behavior<br>(4) Self-monitoring of outcome(s) of behavior)<br>(5) Monitoring outcome(s) of behavior by others without feedback<br>(6) Biofeedback<br>(7) Feedback on outcome(s) of behavior<br>(999) Not applicable |
| 7.10.11 | Behavior change technique – Social support | Social support: For this component, what behavior change techniques map to the intervention activity described above? | *Select multiple*<br>(1) Social support (unspecified)<br>(2) Social support (practical)<br>(3) Social support (emotional)<br>(999) Not applicable |

| | | | |
|---|---|---|---|
| 7.10.12 | Behavior change technique – Shaping knowledge | Shaping knowledge: For this component, what behavior change techniques map to the intervention activity described above? | *Select multiple*<br>(1) Instruction on how to perform a behavior<br>(2) Information about antecedents<br>(3) Re-attribution<br>(4) Behavioral experiments<br>(999) Not applicable |
| 7.10.13 | Behavior change technique – Natural consequences | Natural consequences: For this component, what behavior change techniques map to the intervention activity described above? | *Select multiple*<br>(1) Information about health consequences<br>(2) Salience of consequences<br>(3) Information about social and environmental consequences<br>(4) Monitoring of emotional consequences<br>(5) Anticipated regret<br>(6) Information about emotional consequences<br>(999) Not applicable |
| 7.10.14 | Behavior change technique – Comparison of behavior | Comparison of behavior: For this component, what behavior change techniques map to the intervention activity described above? | *Select multiple*<br>(1) Demonstration of behavior<br>(2) Social comparison<br>(3) Information about others' approval<br>(999) Not applicable |
| 7.10.15 | Behavior change technique – Associations | Associations: For this component, what behavior change techniques map to the intervention activity described above? | *Select multiple*<br>(1) Prompts/cues<br>(2) Cue signaling reward<br>(3) Reduce prompts/cues<br>(4) Remove access to the reward<br>(5) Remove aversive stimulus |

| | | | |
|---|---|---|---|
| | | | (6) Satiation<br>(7) Exposure<br>(8) Associative learning<br>(999) Not applicable |
| 7.10.16 | Behavior change technique – Repetition and substitution | Repetition and substitution: For this component, what behavior change techniques map to the intervention activity described above? | *Select multiple*<br>(1) Behavioral practice/rehearsal<br>(2) Behavior substitution<br>(3) Habit formation<br>(4) Habit reversal<br>(5) Overcorrection<br>(6) Generalization of a target behavior<br>(7) Graded tasks<br>(999) Not applicable |
| 7.10.17 | Behavior change technique – Comparison of outcomes | Comparison of outcomes: For this component, what behavior change techniques map to the intervention activity described above? | *Select multiple*<br>(1) Credible source<br>(2) Pros and cons<br>(3) Comparative imagining of future outcomes<br>(999) Not applicable |
| 7.10.18 | Behavior change technique – Reward and threat | Reward and threat: For this component, what behavior change techniques map to the intervention activity described above? | *Select multiple*<br>(1) Material incentive (behavior)<br>(2) Material reward (behavior)<br>(3) Non-specific reward<br>(4) Social reward<br>(5) Social incentive<br>(6) Non-specific incentive<br>(7) Self-incentive<br>(8) Incentive (outcome)<br>(9) Self-reward<br>(10) Reward (outcome)<br>(11) Future punishment |

| | | | |
|---|---|---|---|
| | | | (999) Not applicable |
| 7.10.19 | Behavior change technique – Regulation | Regulation: For this component, what behavior change techniques map to the intervention activity described above? | *Select multiple*<br>(1) Pharmocological support<br>(2) Reduce negative emotions<br>(3) Conserving mental resources<br>(4) Paradoxical instructions<br>(999) Not applicable |
| 7.10.20 | Behavior change technique – Antecedents | Antecedents: For this component, what behavior change techniques map to the intervention activity described above? | *Select multiple*<br>(1) Restructuring the physical environment<br>(2) Restructuring the social environment<br>(3) Avoidance/reducing exposure to cues for the behavior<br>(4) Distraction<br>(5) Adding objects to the environment<br>(6) Body changes<br>(999) Not applicable |
| 7.10.21 | Behavior change technique – Identity | Identity: For this component, what behavior change techniques map to the intervention activity described above? | *Select multiple*<br>(1) Identification of self as role model<br>(2) Framing/reframing<br>(3) Incompatible beliefs<br>(4) Valued self-identity<br>(5) Identity associated with changed behavior<br>(999) Not applicable |
| 7.10.22 | Behavior change technique – Scheduled consequences | Scheduled consequences: For this component, what behavior change techniques map to the | *Select multiple*<br>(1) Behavior cost<br>(2) Punishment<br>(3) Remove reward |

| | | | |
|---|---|---|---|
| | | intervention activity described above? | (4) Reward approximation<br>(5) Rewarding completion<br>(6) Situation-specific reward<br>(7) Reward incompatible behavior<br>(8) Reward alternative behavior<br>(9) Reduce reward frequency<br>(10) Remove punishment<br>(999) Not applicable |
| 7.10.23 | Behavior change technique – Self-belief | Self-belief: For this component, what behavior change techniques map to the intervention activity described above? | *Select multiple*<br>(1) Verbal persuasion about capability<br>(2) Mental rehearsal of successful performance<br>(3) Focus on past success<br>(4) Self-talk<br>(999) Not applicable |
| 7.10.24 | Behavior change technique – Covert learning | Covert learning: For this component, what behavior change techniques map to the intervention activity described above? | *Select multiple*<br>(1) Imaginary punishment<br>(2) Imaginary reward<br>(3) Vicarious consequences<br>(999) Not applicable |
| 7.11 | Other intervention | Did the study implement another hand hygiene intervention not described above? | *Select one*<br>(0) No<br>(1) Yes<br>(999) Not applicable |
| 7.11.1 | Intervention text | For this component, please copy in the author's description. | Text<br>(888) Not reported<br>(999) Not applicable |
| 7.11.2 | Multi-arm | If the study is a multi-arm trial, which arm provided this intervention? | Text<br>(999) Not applicable |

| | | | |
|---|---|---|---|
| 7.11.3 | Explicit barrier | What barrier did the authors explicitly note that this intervention activity addressed? | Text<br>(888) Not noted<br>(999) Not applicable |
| 7.11.4 | Assumed barriers | If the barrier was not noted by the authors, what is the assumed barrier? | Text<br>(999) Not applicable |
| 7.11.5 | Explicit enabler | What enabler did the authors explicitly note that this intervention activity addressed? | Text<br>(888) Not noted<br>(999) Not applicable |
| 7.11.6 | Assumed enabler | If the enabler was not noted by the authors, what is the assumed enabler? | Text<br>(999) Not applicable |
| 7.11.7 | COM-B category | Which COM-B category was addressed through the intervention activity? | *Select multiple*<br>(1) Physical capability<br>(2) Psychological capability<br>(3) Physical opportunity<br>(4) Social opportunity<br>(5) Automatic motivation<br>(6) Reflective motivation<br>(999) Not applicable |
| 7.11.8 | Intervention function | Which COM-B intervention function maps to the behavior change technique above? | *Select multiple*<br>(1) Education<br>(2) Persuasion<br>(3) Incentivization<br>(4) Coercion<br>(5) Training<br>(6) Restriction<br>(7) Environmental Restructuring<br>(8) Modelling<br>(9) Enablement<br>(999) Not applicable |

| | | | |
|---|---|---|---|
| 7.11.9 | Behavior change technique – Goals and planning | Goals and planning: For this component, what behavior change techniques map to the intervention activity described above? | *Select multiple*<br>(1) Goal setting (behavior)<br>(2) Problem solving<br>(3) Goal setting (outcome)<br>(4) Action planning<br>(5) Review behavior goals<br>(6) Discrepancy between current behavior and goal<br>(7) Review outcome goals<br>(8) Behavioral contract<br>(9) Commitment<br>(999) Not applicable |
| 7.11.10 | Behavior change technique – Feedback and monitoring | Feedback and monitoring: For this component, what behavior change techniques map to the intervention activity described above? | *Select multiple*<br>(1) Monitoring of behavior by others without feedback<br>(2) Feedback on behavior<br>(3) Self-monitoring of behavior<br>(4) Self-monitoring of outcome(s) of behavior)<br>(5) Monitoring outcome(s) of behavior by others without feedback<br>(6) Biofeedback<br>(7) Feedback on outcome(s) of behavior<br>(999) Not applicable |
| 7.11.11 | Behavior change technique – Social support | Social support: For this component, what behavior change techniques map to the intervention activity described above? | *Select multiple*<br>(1) Social support (unspecified)<br>(2) Social support (practical)<br>(3) Social support (emotional)<br>(999) Not applicable |

| | | | |
|---|---|---|---|
| 7.11.12 | Behavior change technique – Shaping knowledge | Shaping knowledge: For this component, what behavior change techniques map to the intervention activity described above? | *Select multiple*<br>(1) Instruction on how to perform a behavior<br>(2) Information about antecedents<br>(3) Re-attribution<br>(4) Behavioral experiments<br>(999) Not applicable |
| 7.11.13 | Behavior change technique – Natural consequences | Natural consequences: For this component, what behavior change techniques map to the intervention activity described above? | *Select multiple*<br>(1) Information about health consequences<br>(2) Salience of consequences<br>(3) Information about social and environmental consequences<br>(4) Monitoring of emotional consequences<br>(5) Anticipated regret<br>(6) Information about emotional consequences<br>(999) Not applicable |
| 7.11.14 | Behavior change technique – Comparison of behavior | Comparison of behavior: For this component, what behavior change techniques map to the intervention activity described above? | *Select multiple*<br>(1) Demonstration of behavior<br>(2) Social comparison<br>(3) Information about others' approval<br>(999) Not applicable |
| 7.11.15 | Behavior change technique – Associations | Associations: For this component, what behavior change techniques map to the intervention activity described above? | *Select multiple*<br>(1) Prompts/cues<br>(2) Cue signaling reward<br>(3) Reduce prompts/cues<br>(4) Remove access to the reward<br>(5) Remove aversive stimulus |

| | | | |
|---|---|---|---|
| | | | (6) Satiation<br>(7) Exposure<br>(8) Associative learning<br>(999) Not applicable |
| 7.11.16 | Behavior change technique – Repetition and substitution | Repetition and substitution: For this component, what behavior change techniques map to the intervention activity described above? | *Select multiple*<br>(1) Behavioral practice/rehearsal<br>(2) Behavior substitution<br>(3) Habit formation<br>(4) Habit reversal<br>(5) Overcorrection<br>(6) Generalization of a target behavior<br>(7) Graded tasks<br>(999) Not applicable |
| 7.11.17 | Behavior change technique – Comparison of outcomes | Comparison of outcomes: For this component, what behavior change techniques map to the intervention activity described above? | *Select multiple*<br>(1) Credible source<br>(2) Pros and cons<br>(3) Comparative imagining of future outcomes<br>(999) Not applicable |
| 7.11.18 | Behavior change technique – Reward and threat | Reward and threat: For this component, what behavior change techniques map to the intervention activity described above? | *Select multiple*<br>(1) Material incentive (behavior)<br>(2) Material reward (behavior)<br>(3) Non-specific reward<br>(4) Social reward<br>(5) Social incentive<br>(6) Non-specific incentive<br>(7) Self-incentive<br>(8) Incentive (outcome)<br>(9) Self-reward<br>(10) Reward (outcome)<br>(11) Future punishment |

| | | | |
|---|---|---|---|
| | | | (999) Not applicable |
| 7.11.19 | Behavior change technique – Regulation | Regulation: For this component, what behavior change techniques map to the intervention activity described above? | *Select multiple*<br>(1) Pharmocological support<br>(2) Reduce negative emotions<br>(3) Conserving mental resources<br>(4) Paradoxical instructions<br>(999) Not applicable |
| 7.11.20 | Behavior change technique – Antecedents | Antecedents: For this component, what behavior change techniques map to the intervention activity described above? | *Select multiple*<br>(1) Restructuring the physical environment<br>(2) Restructuring the social environment<br>(3) Avoidance/reducing exposure to cues for the behavior<br>(4) Distraction<br>(5) Adding objects to the environment<br>(6) Body changes<br>(999) Not applicable |
| 7.11.21 | Behavior change technique – Identity | Identity: For this component, what behavior change techniques map to the intervention activity described above? | *Select multiple*<br>(1) Identification of self as role model<br>(2) Framing/reframing<br>(3) Incompatible beliefs<br>(4) Valued self-identity<br>(5) Identity associated with changed behavior<br>(999) Not applicable |
| 7.11.22 | Behavior change technique – Scheduled consequences | Scheduled consequences: For this component, what behavior change techniques map to the | *Select multiple*<br>(1) Behavior cost<br>(2) Punishment<br>(3) Remove reward |

| | | | |
|---|---|---|---|
| | | intervention activity described above? | (4) Reward approximation<br>(5) Rewarding completion<br>(6) Situation-specific reward<br>(7) Reward incompatible behavior<br>(8) Reward alternative behavior<br>(9) Reduce reward frequency<br>(10) Remove punishment<br>(999) Not applicable |
| 7.11.23 | Behavior change technique – Self-belief | Self-belief: For this component, what behavior change techniques map to the intervention activity described above? | *Select multiple*<br>(1) Verbal persuasion about capability<br>(2) Mental rehearsal of successful performance<br>(3) Focus on past success<br>(4) Self-talk<br>(999) Not applicable |
| 7.11.24 | Behavior change technique – Covert learning | Covert learning: For this component, what behavior change techniques map to the intervention activity described above? | *Select multiple*<br>(1) Imaginary punishment<br>(2) Imaginary reward<br>(3) Vicarious consequences<br>(999) Not applicable |
| **8. Hand Hygiene** | | | |
| 8.1 | Practice | Type of practice (which can still be check multiple) | *Check multiple*<br>(1) Handwashing with soap and water<br>(2) Handwashing with water<br>(3) Handwashing with alcohol-based hand rub<br>(4) Handwashing with non-alcoholic antiseptics |

| | | | |
|---|---|---|---|
| | | | (5) Handwashing with soap alternatives (e.g., ash)<br>(6) Handwashing - unspecified<br>(7) Hand drying<br>(777) Other – specify<br>(888) Not reported<br>(999) Not applicable |
| 8.2 | Outcome category | What category does this outcome fall into? | *Select one*<br>(1) Frequency of handwashing<br>(2) Efficacy of handwashing<br>(3) Consistency of handwashing<br>(777) Other - specify |
| 8.3 | Key moment | What key moment was this outcome looking at? | *Select one*<br>(1) Before, during, and after preparing food<br>(2) Before and after eating food<br>(3) Before and after caring for someone who is sick with vomiting or diarrhea<br>(4) Before and after treating a cut or wound<br>(5) Before feeding a child<br>(6) After using the toilet<br>(7) After changing diapers/cleaning a child's bottom<br>(8) After blowing your nose, coughing or sneezing<br>(9) After touching an animal, animal feed, or animal waste<br>(10) After touching garbage |

| | | | |
|---|---|---|---|
| | | | (11) After handling pet food or pet treats<br>(12) Key moments index – combination<br>(13) No specified key moment<br>(777) Other - specify |
| 8.4 | Participants | Copy in the population that's measured by the outcome | Text<br>(999) Not applicable |
| 8.5 | Study arm | If effect estimate is reported separately by study arm, indicate which arm is below. | Text<br>(999) Not applicable |
| 8.6 | Measurement | How was this outcome measured? | *Select one*<br>(1) Direct observation<br>(2) Self-reported<br>(3) Proxy indicator<br>(888) Not reported<br>(999) Not applicable |
| 8.6.1 | Direct observation indicator | Paste in the indicator/outcome description that authors used for measurement. | Text<br>(999) Not applicable |
| 8.6.2 | Self-report | If hand hygiene is assessed by self-reporting, paste the survey question used | Text<br>(999) Not applicable |
| 8.6.3 | Proxy indicator | If hand hygiene is assessed by proxy indicator, paste the indicator used | Text<br>(999) Not applicable |
| 8.6.4 | Sex disaggregation | Was the hand hygiene practice(s) above disaggregated by sex? | *Select one*<br>(0) No<br>(1) Yes<br>(999) Not applicable |
| 8.6.5 | Effectiveness | Did the intervention have an impact on this outcome?<br>(Note: Effectiveness is determined by study authors' determination) | *Select one*<br>(0) No<br>(1) Yes<br>(888) Not reported |

| | | | |
|---|---|---|---|
| | | | (999) Not applicable |
| 8.7.1 | Intervention group observations | What is the number of observations in the intervention group for this outcome? | Text<br>(888) Not reported<br>(999) Not applicable |
| 8.7.2 | Additional intervention group observations | What is the number of observations in the additional intervention group for this outcome? | Text<br>(888) Not reported<br>(999) Not applicable |
| 8.7.3 | Control group observations | What is the number of observations in the control group for this outcome? | Text<br>(888) Not reported<br>(999) Not applicable |
| 8.8.1 | Effect estimate | What is the effect estimate of hand hygiene practice? | Text<br>(888) Not reported<br>(999) Not applicable |
| 8.8.2 | Effect measure type | What type of effect measure is provided above? | *Select one*<br>(1) Risk ratio<br>(2) Odds ratio<br>(3) Prevalence ratio<br>(4) Rate ratio<br>(5) Mean difference<br>(6) Risk difference<br>(7) Proportion<br>(8) Number of events<br>(9) Percentage<br>(777) Other – specify<br>(999) Not applicable |
| 8.8.3 | Adjusted | Was the effect measure adjusted? | *Select one*<br>(1) No<br>(2) Yes<br>(888) Not reported<br>(999) Not applicable |
| 8.8.4 | Units | Copy in the units used for the effect (i.e. percentage points, number of events, etc.) | Text<br>(888) Not reported<br>(999) Not applicable |

| | | | | |
|---|---|---|---|---|
| 8.8.5 | Lower CI | What is the lower confidence interval bound? | Text<br>(888) Not reported<br>(999) Not applicable | |
| 8.8.6 | Upper CI | What is the upper confidence interval bound? | Text<br>(888) Not reported<br>(999) Not applicable | |
| 8.8.7 | p-value | What is the p-value? | Text<br>(888) Not reported<br>(999) Not applicable | |
| 8.8.8 | Standard deviation | What is the standard deviation? | Text<br>(888) Not reported<br>(999) Not applicable | |
| **RQ3.2a Among interventions to improve hand hygiene in community settings, which have been designed using behavior change theories?** | | | | |
| 1. | Theory | Did the intervention report using a behavior change theory in its design? Select all that apply | *Select multiple*<br>(1) IBM-WASH<br>(2) RANAS<br>(3) Behavior Centered Design/Evo-Eco Model<br>(4) COM-B<br>(5) Theory of Planned Behavior<br>(6) Health Belief Model<br>(7) Social Ecological Model<br>(8) Theoretical Domains Framework<br>(777) Other – specify<br>(999) No theory reported | White et al<br><br>The Handwashing Handbook |
| 2 | Theory mention | Where did the authors discuss the theory that they used? | *Select one*<br>(1) Protocol<br>(2) Cited formative research<br>(3) Primary research paper<br>(777) Other – specify<br>(999) No theory reported | |

| RQ3.2d Among interventions to improve hand hygiene in community settings, what hand hygiene station designs have been effective at improving and sustaining hand hygiene? | | | | |
|---|---|---|---|---|
| 1 | Handwashing station | Did the intervention use handwashing stations? | *Select one*<br>(0) No<br>(1) Yes<br>(999) Unclear | |
| 1.1 | Handwashing station design types | If handwashing stations were used, what type was used? | *Select one*<br>(1) Tippy tap<br>(2) Raised bucket with tap/outlet<br>(3) Two buckets suspended<br>(4) Suspended bottle or bag with outlet/hole/pop-up plug<br>(5) Sink with tap<br>(6) Foot pump sink<br>(7) Purpose-built all-in-one system<br>(8) Free standing water tank with taps/outlets<br>(9) Tube with outlets<br>(777) Other - specify<br>(888) Not reported<br>(999) Not applicable | UNICEF (2020) – Handwashing stations |
| 1.2 | Handwashing station design features – fixed vs mobile | If handwashing stations were used, was the station fixed or mobile? | *Select one*<br>(1) Fixed<br>(2) Mobile<br>(888) Not reported<br>(999) Not applicable | UNICEF (2020) – Handwashing stations |
| 1.3 | Handwashing station design features – permanency | If handwashing stations were used, was the station permanent or temporary? | *Select one*<br>(1) Permanent<br>(2) Temporary<br>(888) Not reported<br>(999) Not applicable | UNICEF (2020) – Handwashing stations |

| | | | | |
|---|---|---|---|---|
| 1.4 | Handwashing station design features – users | If handwashing stations were used, who was the primary user base? | *Select one*<br>(1) Household<br>(2) Community<br>(3) Schools<br>(777) Other – specify<br>(888) Not reported<br>(999) Not applicable | UNICEF (2020) – Handwashing stations |
| 1.5 | Handwashing station design features – water supply | If handwashing stations were used, what type of water supply was available? | *Select one*<br>(1) Individual storage tank<br>(2) Community storage tank<br>(3) Piped water<br>(777) Other – specify<br>(888) Not reported<br>(999) Not applicable | UNICEF (2020) – Handwashing stations |
| 1.6 | Handwashing station design features – size | If handwashing stations were used, what was the size of the station? | Text<br>(888) Not reported<br>(999) Not applicable | |
| 1.7 | Handwashing station design features – material | If handwashing station designs were used, what material were they made of? | Text<br>(888) Not reported<br>(999) Not applicable | |
| 1.8 | Handwashing station design features – other | If handwashing stations were used, were there any other design features not yet captured? | Text<br>(888) Not reported<br>(999) Not applicable | |
| **RQ3.2e Among interventions to improve hand hygiene in community settings, what design adaptations (e.g., placement, nudges, and cues) have been effective at improving and sustaining hand hygiene?** | | | | |
| 1 | Design adaptations | Did the intervention assess design adaptations of handwashing stations? | *Select one*<br>(0) No<br>(1) Yes<br>(999) Not applicable | |
| 1.1 | Design adaptations comparison | How are the authors comparing their standard handwashing station design with their adapted design? | *Select one*<br>(1) Through multi-arm trial<br>(2) Comparing to design tested in earlier study<br>(999) Not applicable | |

| | | | |
|---|---|---|---|
| 1.2 | Design adaptations standard | What was the handwashing station design that was the comparison/standard? | Text<br>(999) Not applicable |
| 1.3 | Design adaptations type | If design adaptations were implemented, what type of adaptation was used? | *Select one*<br>(1) Placement<br>(2) Nudges<br>(3) Cues<br>(777) Other – specify<br>(888) Not reported<br>(999) Not applicable |
| 1.4 | Design adaptation examples | If design adaptations were implemented, what were they? | *Text*<br>(888) Not reported<br>(999) Not applicable |
| 1.5 | Design adaptation effectiveness | Was the hand hygiene practice the same, better or worse in the adjusted design? | *Select one*<br>(1) Same<br>(2) Better<br>(3) Worse<br>(999) Not applicable |
| **RQ3.2f Among interventions to improve hand hygiene in community settings, what level of frequency and intensity of behavior change interventions is necessary to effectively improve hand hygiene?** | | | |
| 1 | Frequency | Was the frequency of the behavior change intervention varied? | *Select one*<br>(0) No<br>(1) Yes<br>(999) Unclear |
| 1.1 | Frequency comparison | How are the authors comparing their standard level of frequency for the intervention with their adjusted design? | *Select one*<br>(1) Through multi-arm trial<br>(2) Comparing to design tested in earlier study<br>(999) Not applicable |
| 1.2 | Frequency standard | What level of frequency of the behavior change intervention was used as the comparison/standard? | Text<br>(999) Not applicable |

| | | | |
|---|---|---|---|
| 1.3 | Frequency level | What was the level of frequency of the behavior change intervention for the adjusted design? | Text<br>(999) Not applicable |
| 1.4 | Frequency level – additional arm | If the authors tested additional changes to their design (e.g., varying frequency in a 3rd arm), please provide a description of the design change. | Text<br>(999) Not applicable |
| 1.5 | Frequency effectiveness | Was the hand hygiene practice the same, better or worse in the adjusted design? | *Select one*<br>(1) Same<br>(2) Better<br>(3) Worse<br>(999) Not applicable |
| 2 | Intensity | Was the intensity of the behavior change intervention varied? | *Select one*<br>(0) No<br>(1) Yes<br>(999) Unclear |
| 2.1 | Intensity comparison | How are the authors comparing their standard level of intensity for the intervention with their adjusted design? | *Select one*<br>(1) Through multi-arm trial<br>(2) Comparing to design tested in earlier study<br>(999) Not applicable |
| 2.2 | Intensity standard | What level of intensity of the behavior change intervention was used as the comparison/standard? | Text<br>(999) Not applicable |
| 2.3 | Intensity level | What was the level of intensity of the behavior change intervention for the adjusted design? | Text<br>(999) Not applicable |
| 2.4 | Intensity level – additional arm | If the authors tested additional changes to their design (e.g., varying intensity in a 3rd arm), | Text<br>(999) Not applicable |

| | | | |
|---|---|---|---|
| | | please provide a description of the design change. | |
| 2.5 | Intensity effectiveness | Was the hand hygiene practice the same, better or worse in the adjusted design? | *Select one*<br>(1) Same<br>(2) Better<br>(3) Worse<br>(999) Not applicable |
| **RQ3.2g Among interventions to improve hand hygiene in community settings, how do hand hygiene practices vary by population groups, risk scenarios, or over time?** | | | |
| 1 | Risk scenarios | Did the study evaluate hand hygiene in a risk scenario? | *Select one*<br>(0) No<br>(1) Yes<br>(999) Unclear |
| 1.1 | Risk scenario type | If yes, what was the risk scenario? | *Select one*<br>(1) COVID-19<br>(2) Flu<br>(3) Earthquake<br>(4) Flood<br>(5) Typhoon<br>(6) Forced migration<br>(7) Internal displacement<br>(8) Emergency setting<br>(777) Other – specify<br>(999) Not applicable |
| 1.2 | Risk scenario text | Please copy in the author's text about the risk scenario. | Text<br>(999) Not applicable |
| 2 | Over time | Did the study collect data at more than one time point after the intervention was implemented? | *Select one*<br>(0) No<br>(1) Yes<br>(999) Unclear |
| 2.1 | Over time – data points | If yes, how many time points did the study authors collect? | Text<br>(999) Not applicable |
| 2.2 | Outcome category | What category does this outcome fall into? | *Select one*<br>(1) Frequency of handwashing |

| | | | |
|---|---|---|---|
| | | | (2) Efficacy of handwashing<br>(3) Consistency of handwashing<br>(777) Other - specify |
| 2.3 | Key moment | What key moment was this outcome looking at? | *Select one*<br>(1) Before, during, and after preparing food<br>(2) Before and after eating food<br>(3) Before and after caring for someone who is sick with vomiting or diarrhea<br>(4) Before and after treating a cut or wound<br>(5) Before feeding a child<br>(6) After using the toilet<br>(7) After changing diapers/cleaning a child's bottom<br>(8) After blowing your nose, coughing or sneezing<br>(9) After touching an animal, animal feed, or animal waste<br>(10) After touching garbage<br>(11) After handling pet food or pet treats<br>(12) Key moments index – combination<br>(13) No specified key moment<br>(777) Other - specify |
| 2.4 | Participants | Copy in the population that's measured by the outcome | Text<br>(999) Not applicable |

| | | | |
|---|---|---|---|
| 2.5 | Study arm | If effect estimate is reported separately by study arm, indicate which arm is below. | Text<br>(999) Not applicable |
| 2.6 | Measurement | How was this outcome measured? | *Select one*<br>(1) Direct observation<br>(2) Self-reported<br>(3) Proxy indicator<br>(888) Not reported<br>(999) Not applicable |
| 2.6.1 | Direct observation indicator | Paste in the indicator/outcome description that authors used for measurement. | Text<br>(999) Not applicable |
| 2.6.2 | Self-report | If hand hygiene is assessed by self-reporting, paste the survey question used | Text<br>(999) Not applicable |
| 2.6.3 | Proxy indicator | If hand hygiene is assessed by proxy indicator, paste the indicator used | Text<br>(999) Not applicable |
| 2.6.4 | Sex disaggregation | Was the hand hygiene practice(s) above disaggregated by sex? | *Select one*<br>(0) No<br>(1) Yes<br>(999) Not applicable |
| 2.6.5 | Time elapsed | How much time has elapsed since the intervention | Text<br>(999) Not applicable |
| 2.7.1 | Intervention group observations | What is the number of observations in the intervention group for this outcome? | Text<br>(888) Not reported<br>(999) Not applicable |
| 2.7.2 | Additional intervention group observations | What is the number of observations in the additional intervention group for this outcome? | Text<br>(888) Not reported<br>(999) Not applicable |
| 2.7.3 | Control group observations | What is the number of observations in the control group for this outcome? | Text<br>(888) Not reported<br>(999) Not applicable |

| | | | |
|---|---|---|---|
| 2.8.1 | Effect estimate | What is the effect estimate of hand hygiene practice? | Text<br>(888) Not reported<br>(999) Not applicable |
| 2.8.2 | Effect measure type | What type of effect measure is provided above? | *Select one*<br>(1) Risk ratio<br>(2) Odds ratio<br>(3) Prevalence ratio<br>(4) Rate ratio<br>(5) Mean difference<br>(6) Risk difference<br>(7) Proportion<br>(8) Number of events<br>(9) Percentage<br>(777) Other – specify<br>(999) Not applicable |
| 2.8.3 | Adjusted | Was the effect measure adjusted? | *Select one*<br>(1) No<br>(2) Yes<br>(888) Not reported<br>(999) Not applicable |
| 2.8.4 | Units | Copy in the units used for the effect (i.e. percentage points, number of events, etc.) | Text<br>(888) Not reported<br>(999) Not applicable |
| 2.8.5 | Lower CI | What is the lower confidence interval bound? | Text<br>(888) Not reported<br>(999) Not applicable |
| 2.8.6 | Upper CI | What is the upper confidence interval bound? | Text<br>(888) Not reported<br>(999) Not applicable |
| 2.8.7 | p-value | What is the p-value? | Text<br>(888) Not reported<br>(999) Not applicable |
| 2.8.8 | Standard deviation | What is the standard deviation? | Text |

| | | | |
|---|---|---|---|
| | | | (888) Not reported<br>(999) Not applicable |
| | Notes | Any notes or comments to add about the study? | Text |

### Bias assessment

| **Bias Assessment** | | | | |
|---|---|---|---|---|
| MMAT (All articles) | | | | |
| S1 | Screening question 1 (for all types) | Are there clear research questions? | (0) No<br>(1) Yes<br>(888) Can't tell<br>(999) Not applicable | MMAT User Guide |
| S2 | Screening question 2 (for all types) | Do the collected data allow to address the research questions? | (0) No<br>(1) Yes<br>(888) Can't tell<br>(999) Not applicable | MMAT User Guide |
| Qualitative | | | | |
| 1.1 | Is the qualitative approach appropriate to answer the research question? | | (0) No<br>(1) Yes<br>(888) Can't tell<br>(999) Not applicable | MMAT User Guide |
| 1.2 | Are the qualitative data collection methods adequate to address the research question? | | (0) No<br>(1) Yes<br>(888) Can't tell<br>(999) Not applicable | MMAT User Guide |
| 1.3 | Are the findings adequately derived from the data? | | (0) No<br>(1) Yes<br>(888) Can't tell<br>(999) Not applicable | MMAT User Guide |
| 1.4 | Is the interpretation of results sufficiently substantiated by data? | | (0) No<br>(1) Yes<br>(888) Can't tell<br>(999) Not applicable | MMAT User Guide |
| 1.5 | Is there coherence between qualitative data sources, collection, analysis and interpretation? | | (0) No<br>(1) Yes<br>(888) Can't tell<br>(999) Not applicable | MMAT User Guide |
| Quantitative randomized controlled trials | | | | |

| | | | |
|---|---|---|---|
| 2.1 | Is randomization appropriately performed? | (0) No<br>(1) Yes<br>(888) Can't tell<br>(999) Not applicable | MMAT User Guide |
| 2.2 | Are the groups comparable at baseline? | (0) No<br>(1) Yes<br>(888) Can't tell<br>(999) Not applicable | MMAT User Guide |
| 2.3 | Are there complete outcome data? | (0) No<br>(1) Yes<br>(888) Can't tell<br>(999) Not applicable | MMAT User Guide |
| 2.4 | Are outcome assessors blinded to the intervention provided? | (0) No<br>(1) Yes<br>(888) Can't tell<br>(999) Not applicable | MMAT User Guide |
| 2.5 | Did the participants adhere to the assigned intervention? | (0) No<br>(1) Yes<br>(888) Can't tell<br>(999) Not applicable | MMAT User Guide |
| Quantitative non-randomized | | | |
| 3.1 | Are the participants representative of the target population? | (0) No<br>(1) Yes<br>(888) Can't tell<br>(999) Not applicable | MMAT User Guide |
| 3.2 | Are measurements appropriate regarding both the outcome and intervention (or exposure)? | (0) No<br>(1) Yes<br>(888) Can't tell<br>(999) Not applicable | MMAT User Guide |
| 3.3 | Are there complete outcome data? | (0) No<br>(1) Yes<br>(888) Can't tell<br>(999) Not applicable | MMAT User Guide |

| | | | |
|---|---|---|---|
| 3.4 | Are the confounders accounted for in the design and analysis? | (0) No<br>(1) Yes<br>(888) Can't tell<br>(999) Not applicable | MMAT User Guide |
| 3.5 | During the study period, is the intervention administered (or exposure occurred) as intended? | (0) No<br>(1) Yes<br>(888) Can't tell<br>(999) Not applicable | MMAT User Guide |
| Quantitative descriptive | | | |
| 4.1 | Is the sampling strategy relevant to address the research question? | (0) No<br>(1) Yes<br>(888) Can't tell<br>(999) Not applicable | MMAT User Guide |
| 4.2 | Is the sample representative of the target population? | (0) No<br>(1) Yes<br>(888) Can't tell<br>(999) Not applicable | MMAT User Guide |
| 4.3 | Are the measurements appropriate? | (0) No<br>(1) Yes<br>(888) Can't tell<br>(999) Not applicable | MMAT User Guide |
| 4.4 | Is the risk of nonresponse bias low? | (0) No<br>(1) Yes<br>(888) Can't tell<br>(999) Not applicable | MMAT User Guide |
| 4.5 | Is the statistical analysis appropriate to answer the research question? | (0) No<br>(1) Yes<br>(888) Can't tell<br>(999) Not applicable | MMAT User Guide |
| Mixed methods | | | |
| 5.1 | Is there an adequate rationale for using a mixed methods design to address the research question? | (0) No<br>(1) Yes<br>(888) Can't tell<br>(999) Not applicable | MMAT User Guide |

| | | | |
|---|---|---|---|
| 5.2 | Are the different components of the study effectively integrated to answer the research question? | (0) No<br>(1) Yes<br>(888) Can't tell<br>(999) Not applicable | MMAT User Guide |
| 5.3 | Are the outputs of the integration of qualitative and quantitative components adequately interpreted? | (0) No<br>(1) Yes<br>(888) Can't tell<br>(999) Not applicable | MMAT User Guide |
| 5.4 | Are divergences and inconsistencies between quantitative and qualitative results adequately addressed? | (0) No<br>(1) Yes<br>(888) Can't tell<br>(999) Not applicable | MMAT User Guide |
| 5.5 | Do the different components of the study adhere to the quality criteria of each tradition of the methods involved? | (0) No<br>(1) Yes<br>(888) Can't tell<br>(999) Not applicable | MMAT User Guide |
