## Supplementary material for "Interventions to improve hand hygiene in community settings: A systematic review of theories, barriers and enablers, behavior change techniques, and hand hygiene station design features": S5

**S5 –** Overview of included studies

**S5 –** Overview of included studies

**Supplemental Table 5**. Included studies and key characteristics

| **Study ID** | **Design** | **Country** | **Location** | **Setting** | **Participant category** | **Theory used** | **Hand hygiene outcome(s)** | **Quality Appraisal** | **Effective?** |
|---|---|---|---|---|---|---|---|---|---|
| Abbot 2012 | Non-randomized quantitative | United States | Unspecified | Universities | Adults (Women and Men) | Theory of Planned Behavior; Theory of reasoned action | Handwashing with soap and water | **2** | Yes |
| Aboud 2011 | Randomized controlled trial | Bangladesh | Rural | Households | Mother-child dyads | Social-cognitive learning theory | Handwashing - unspecified | **4** | Yes |
| Advaita 2021 | Non-randomized quantitative | India | Urban | Households | Children (Boys only) | No theory reported | Handwashing - unspecified | **3** | Yes |
| Aibana 2013 | Non-randomized quantitative | Haiti | Rural | Households | General populations | No theory reported | Handwashing with soap and water | **4** | Yes |
| Aiello 2012 | Randomized controlled trial | United States | Unspecified | Universities | Adults (Women and Men) | No theory reported | Handrubbing with alcohol-based rub | **4** | Yes |
| Akina 2020 | Randomized controlled trial | Nepal | Unspecified | Schools | Children (Girls and Boys) | No theory reported | Handwashing - unspecified | **3** | Yes |
| Akuokoasibey 1994 | Non-randomized quantitative | Ghana | Rural | Households | General populations | No theory reported | Handwashing with water; Handwashing with soap and water | **3** | Yes |
| Alam 1989 | Non-randomized quantitative | Bangladesh | Rural | Households | Mother-child dyads | No theory reported | Handwashing with water; Handwashing with soap alternatives | **4** | Yes |
| Alexander 2012 | Non-randomized quantitative | India | Rural | Households; Schools | General populations; Children (Girls and Boys) | No theory reported | Handwashing - unspecified | **4** | Yes |
| Ali 2020 | Non-randomized quantitative | Liberia | Urban | Households | General populations | No theory reported | Handwashing with soap alternatives | **3** | Yes |

**S5 –** Overview of included studies

| | | | | | | | | | |
|---|---|---|---|---|---|---|---|---|---|
| Alkon 2009 | Randomized controlled trial | United States | Rural; Urban | Childcare centers | Non-food occupational workers | No theory reported | Handwashing - unspecified | **4** | Yes |
| Almazan 2014 | Non-randomized quantitative | The Philippines | Rural | Households | Adults (Women and Men) | No theory reported | Handwashing with water; Handwashing with soap and water | **3** | Yes |
| Amon-Tanoh 2021 | Randomized controlled trial | Côte d'Ivoire | Urban | Households | General populations | Theory of normative social behaviour | Handwashing with soap and water | **5** | Yes |
| Andrade 2019 | Non-randomized quantitative | El Salvador | Rural | Households; Schools | General populations | No theory reported | Handwashing with soap and water | **4** | Yes |
| Anu 2018 | Mixed methods | Malawi | Rural | Households | Adults (Women and Men) | No theory reported | Handwashing with soap and water | **3** | Yes |
| Appiah-Brempong 2020 | Randomized controlled trial | Ghana | Unspecified | Schools | Children (Girls and Boys) | Bloom's Taxonomy of Learning Theory Social Cognitive Theory; Health Belief Model; Theory of Planned Behavior | Handwashing with soap and water | **5** | Yes |
| Ar 2008 | Non-randomized quantitative | Turkey | Unspecified | Schools | Adults (Women and Men); Children (Girls and Boys) | No theory reported | Handwashing - unspecified | **3** | Yes |
| Ara 2022 | Randomized controlled trial | Bangladesh | Rural | Households | Mother-child dyads | No theory reported | Handwashing with soap and water | **4** | Yes |
| Aragie 2021 | Randomized controlled trial | Ethiopia | Rural | Households; Schools | Children (Girls and Boys) | No theory reported | Handwashing with soap and water | **4** | Yes |
| Arbianingsih 2018 | Non-randomized quantitative | Indonesia | Unspecified | Schools | General populations; Children | No theory reported | Handwashing with soap and water | **2** | Yes |

**S5 –** Overview of included studies

| | | | | | | | | | |
|---|---|---|---|---|---|---|---|---|---|
| | | | | | (Girls and Boys) | | | | |
| Arnold 2009 | Non-randomized quantitative | Guatemala | Rural | Households | Children (Girls and Boys) | No theory reported | Handwashing with soap and water | **4** | No |
| Ankan 2018 | Randomized controlled trial | Turkey | Unspecified | Schools | Children (Girls and Boys) | No theory reported | Handwashing with soap and water | **1** | Yes |
| Ashtarian 2020 | Randomized controlled trial | Iran | Urban | Schools | Children (Girls and Boys) | No theory reported | Handwashing with soap and water | **3** | Yes |
| Ashutosh 2015 | Non-randomized quantitative | India | Urban | Schools | Children (Girls and Boys) | No theory reported | Handwashing with soap and water | **3** | Yes |
| Au 2010 | Non-randomized quantitative | China | Urban | Schools | Children (Girls and Boys) | No theory reported | Handwashing - unspecified | **3** | No |
| Ãzyazıcıoğlu 2011 | Non-randomized quantitative | Turkey | Rural | Schools | Children (Girls and Boys) | No theory reported | Handwashing with soap and water | **3** | Yes |
| Bai 2022 | Non-randomized quantitative | China | Unspecified | Schools | Children (Boys only) | Behavior Place Theory | Handwashing - unspecified | **3** | Yes |
| Bajracharya 2003 | Non-randomized quantitative | Myanmar | Unspecified | Households | General populations | No theory reported | Handwashing with soap and water; Handwashing with soap alternatives | **2** | Yes |
| Bickford 2017 | Non-randomized quantitative | Bangladesh; India | Rural; Urban | Households | Children (Girls and Boys) | No theory reported | Handwashing with soap and water | **4** | Yes |
| Bieri 2013 | Randomized controlled trial | China | Rural | Schools | Children (Girls and Boys) | No theory reported | Handwashing with soap and water | **3** | Yes |
| Biran 2009 | Randomized controlled trial | India | Rural | Households | Adults (Women and Men); Children (Girls and Boys) | No theory reported | Handwashing with soap and water | **4** | No |
| Biran 2014 | Randomized controlled trial | India | Rural | Households | General populations | Behavior-centered | Handwashing with soap and water | **4** | Yes |

**S5 –** Overview of included studies

| | | | | | | | | | |
|---|---|---|---|---|---|---|---|---|---|
| | | | | | | Design/Evo-Eco Model | | | |
| Biran 2020 | Randomized controlled trial | Nigeria | Rural | Households | Adults (Women only) | Behavior-centered Design/Evo-Eco Model | Handwashing with soap and water | **4** | No |
| Biswas 2019 | Randomized controlled trial | Bangladesh | Urban | Schools | Children (Girls and Boys) | No theory reported | Handwashing with water; Handwashing with soap and water; Handrubbing with alcohol-based rub | **2** | Yes |
| Blanton 2010 | Non-randomized quantitative | Kenya | Rural | Schools | Children (Girls and Boys) | No theory reported | Handwashing with soap and water | **3** | Yes |
| Bosomprah 2016 | Non-randomized quantitative | Zambia | Unspecified | Households | Mother-child dyads | No theory reported | Handwashing with soap and water | **3** | Yes |
| Bowen 2013 | Randomized controlled trial | Pakistan | Urban | Households | General populations | No theory reported | Handwashing with soap and water | **2** | Yes |
| Briceño 2017 | Randomized controlled trial | Tanzania | Rural | Households | Mother-child dyads | No theory reported | Handwashing with soap and water | **3** | Yes |
| Briere 2012 | Non-randomized quantitative | Kenya | Rural; Urban | Households | Children (Girls and Boys) | No theory reported | Handwashing - unspecified | **4** | Yes |
| Bulled 2017 | Non-randomized quantitative | South Africa | Unspecified | Schools | Children (Girls and Boys) | RANAS | Handwashing with water | **2** | Yes |
| Burke 2016 | Non-randomized quantitative | United States | Unspecified | Schools | Children (Girls and Boys) | No theory reported | Handwashing - unspecified | **4** | Yes |
| Burns 2018 | Randomized controlled trial | South Africa | Unspecified | Households | Children (Girls and Boys) | No theory reported | Handwashing with soap and water | **3** | Yes |
| Cairncross 2005 | Non-randomized quantitative | India | Rural | Households | General populations | No theory reported | Handwashing - unspecified | **2** | Yes |
| Capps 2022 | Non-randomized quantitative | United States | Urban | Universities | Adults (Women and Men) | Theory of Planned Behavior | Handrubbing with alcohol-based rub | **3** | No |

**S5 –** Overview of included studies

| | | | | | | | | | |
|---|---|---|---|---|---|---|---|---|---|
| Carabin 1999 | Randomized controlled trial | Canada | Unspecified | Schools | Children (Girls and Boys) | No theory reported | Handwashing - unspecified | **3** | Yes |
| CardinaleLagomarsino 2017 | Randomized controlled trial | Argentina | Urban | Universities | Adults (Men only) | No theory reported | Handwashing with soap and water | **3** | Yes |
| Chard 2018 | Randomized controlled trial | Laos | Unspecified | Schools | Children (Girls and Boys) | No theory reported | Handwashing with soap and water | **4** | Yes |
| Contzen 2013 | Non-randomized quantitative | Ethiopia | Rural | Households | Adults (Women only) | RANAS | Handwashing with soap and water | **3** | Yes |
| Costa 2019 | Non-randomized quantitative | Brazil | Unspecified | Schools | Children (Girls and Boys) | No theory reported | Handwashing with soap and water | **3** | Yes |
| Cowling 2009 | Randomized controlled trial | China | Urban | Households | General populations | No theory reported | Handwashing with soap and water; Handrubbing with alcohol-based rub | **3** | No |
| Croghan 2008 | Non-randomized quantitative | United Kingdom | Unspecified | Schools | Children (Girls and Boys) | No theory reported | Handwashing with soap and water | **2** | Yes |
| Curtis 2001 | Non-randomized quantitative | Burkina Faso | Urban | Households; Schools | Adults (Women only) | No theory reported | Handwashing with soap and water | **3** | Yes |
| Davis 2011 | Randomized controlled trial | Tanzania | Peri-urban | Households | Children (Girls and Boys) | No theory reported | Handwashing with soap and water | **3** | Yes |
| Davis 2013 | Non-randomized quantitative | United States | Unspecified | Universities | General populations | No theory reported | Handwashing - unspecified | **4** | Yes |
| Ditai 2019 | Randomized controlled trial | Uganda | Rural | Households | Adults (Women and Men) | No theory reported | Handrubbing with alcohol-based rub | **4** | Yes |
| Dreibelbis 2016 | Randomized controlled trial | Bangladesh | Rural | Schools | Children (Girls and Boys) | No theory reported | Handwashing with soap and water | **3** | Yes |
| Duijster 2020 | Randomized controlled trial | The Philippines | Peri-urban | Schools | Mother-child dyads | No theory reported | Handwashing with soap and water | **2** | No |
| Early 1998 | Non-randomized quantitative | United States | Urban | Schools | Children (Girls and Boys) | No theory reported | Handwashing - unspecified | **3** | Yes |

**S5 –** Overview of included studies

| | | | | | | | | | |
|---|---|---|---|---|---|---|---|---|---|
| Ebuehi 2010 | Non-randomized quantitative | Nigeria | Rural; Urban | Households | Children (Girls and Boys) | No theory reported | Handwashing with soap and water; Handwashing with soap alternatives | **3** | Yes |
| Edward 2019 | Non-randomized quantitative | Kenya; Zambia; Cambodia; Guatemala | Rural | Households | Adults (Women and Men); Children (Girls and Boys) | No theory reported | Handwashing with soap alternatives; Handwashing with soap and water | **5** | Yes |
| ErcanOruc 2020 | Non-randomized quantitative | Ukraine | Unspecified | Universities | Mother-child dyads | No theory reported | Handwashing - unspecified | **3** | Yes |
| Ercumen 2018 | Randomized controlled trial | Bangladesh | Rural | Households | Adults (Women and Men) | No theory reported | Handwashing - unspecified | **3** | No |
| Eun-Joo 2012 | Non-randomized quantitative | South Korea | Unspecified | Schools | Children (Girls and Boys) | Stages of change model | Handwashing with soap and water | **4** | Yes |
| EvansJr 2009 | Non-randomized quantitative | United States | Unspecified | Universities | Children (Girls and Boys) | Ecological theory of health promotion | Handrubbing with alcohol-based rub | **3** | Yes |
| Farhana 2022 | Mixed methods | Bangladesh | Rural | Schools | Adults (Women and Men) | IBM-WASH | Handwashing with soap and water | **3** | Yes |
| Ford 2014 | Non-randomized quantitative | United States | Urban | Universities | Children (Girls and Boys) | No theory reported | Hand drying; Handwashing with soap and water | **2** | Yes |
| Freeman 2020 | Randomized controlled trial | Kenya | Unspecified | Households | Adults (Women and Men) | COM-B | Handwashing with soap and water | **3** | Yes |
| Freeman 2022 | Randomized controlled trial | Ethiopia | Unspecified | Households | General populations; Mother-child dyads | COM-B; RANAS | Handwashing with soap and water | **4** | No |
| Friedrich 2018 | Randomized controlled trial | Zimbabwe | Peri-urban; Rural | Households | General populations | RANAS | Handwashing with soap and water | **3** | Yes |
| Galiani 2016 | Randomized controlled trial | Peru | Urban | Households; Schools | Adults (Women and Men) | No theory reported | Handwashing with soap and water | **4** | Yes |

**S5 –** Overview of included studies

| | | | | | | | | | |
|---|---|---|---|---|---|---|---|---|---|
| Gautam 2017 | Randomized controlled trial | Nepal | Rural | Households | Adults (Women only) | Behavior-centered Design/Evo-Eco Model | Handwashing with soap and water | **5** | Yes |
| Gedamu 2022 | Non-randomized quantitative | Ethiopia | Rural | Households | Mother-child dyads | No theory reported | Handwashing - unspecified | **4** | Yes |
| Geller 1980 | Non-randomized quantitative | United States | Rural | Universities | General populations | No theory reported | Handwashing with soap and water | **3** | Yes |
| George 2017 | Randomized controlled trial | Bangladesh | Urban | Households | Food workers | RANAS | Handwashing with soap and water | **5** | Yes |
| Gimaiyo 2019 | Randomized controlled trial | Kenya | Urban | Householsd | General populations | No theory reported | Handwashing - unspecified | **5** | Yes |
| Goel 2019 | Non-randomized quantitative | India | Rural | Households | General populations | No theory reported | Handwashing with soap and water | **3** | Yes |
| Goel 2020 | Non-randomized quantitative | India | Unspecified | Schools | Mother-child dyads | No theory reported | Handwashing - unspecified | **4** | No |
| Grace 2012 | Non-randomized quantitative | Nigeria | Unspecified | Markets | Children (Girls and Boys) | No theory reported | Handwashing - unspecified | **4** | Yes |
| Greene 2012 | Randomized controlled trial | Kenya | Unspecified | Schools | Food workers | No theory reported | Handwashing - unspecified | **3** | No |
| Greenland 2016 | Randomized controlled trial | Zambia | Unspecified | Households | Children (Girls and Boys) | Behavior-centered Design/Evo-Eco Model | Handwashing with soap and water | **4** | No |
| Grover 2018 | Randomized controlled trial | Bangladesh | Peri-urban; Rural | Schools | Mother-child dyads | Nudge Theory | Handwashing with soap and water | **2** | Yes |
| Guo 2018 | Randomized controlled trial | China | Rural | Households | Children (Girls and Boys) | No theory reported | Handwashing with soap and water | **2** | Yes |
| Hanson 2020 | Non-randomized quantitative | Indonesia | Rural | Households | Children (Girls and Boys) | No theory reported | Handwashing - unspecified | **2** | Yes |
| Her 2019 | Non-randomized quantitative | United States | Rural | Universities | Mother-child dyads | No theory reported | Handwashing with soap and water | **2** | Yes |

**S5 –** Overview of included studies

| | | | | | | | | | |
|---|---|---|---|---|---|---|---|---|---|
| Hetherington 2017 | Mixed methods | Tanzania | Unspecified | Schools | Food workers | IBM-WASH; Social Ecological Model | Handwashing with soap alternatives; Handwashing with soap and water | **4** | Yes |
| Huang 2021 | Mixed methods | Philippines | Rural | Schools | Children (Girls and Boys) | IBM-WASH; Choice-architecture approach | Handwashing with soap and water; Handwashing with water | **4** | Yes |
| Hurley 2021 | Non-randomized quantitative | Malawi | Unspecified | Households | Children (Girls and Boys) | No theory reported | Handwashing - unspecified | **2** | Yes |
| Hussam 2022 | Randomized controlled trial | India | Rural | Households | Children (Girls and Boys); Mother-child dyads | No theory reported | Handwashing with soap and water | **3** | Yes |
| Jafree 2023 | Non-randomized quantitative | Pakistan | Peri-urban; Rural | Households | Adults (Women and Men) | No theory reported | Handwashing - unspecified | **3** | Yes |
| Jagals 2004 | Non-randomized quantitative | South Africa | Rural | Households | Adults (Women only) | No theory reported | Handwashing - unspecified | **2** | No |
| Jetha 2021 | Randomized controlled trial | The Philippines | Urban | Schools | General populations | Behavior-centered Design/Evo-Eco Model | Handwashing - unspecified | **4** | Yes |
| Jinadu 2007 | Non-randomized quantitative | Nigeria | Unspecified | Households | Children (Girls and Boys) | No theory reported | Handwashing with soap and water | **3** | Yes |
| Johnson 2003 | Non-randomized quantitative | United States | Rural | Universities | Mother-child dyads | No theory reported | Handwashing with water; Handwashing with soap and water | **3** | Yes |
| Judah 2009 | Non-randomized quantitative | United Kingdom | Unspecified | Public transportation hubs | Adults (Women and Men) | No theory reported | Handwashing with soap and water | **4** | Yes |
| Kaewchana 2012 | Randomized controlled trial | Thailand | Unspecified | Households | General populations | No theory reported | Handwashing with soap and water | **5** | Yes |
| Kajjura 2019 | Non-randomized quantitative | Uganda | Urban | Households | General populations | Health Belief Model | Handwashing with soap and water | **3** | Yes |

**S5 –** Overview of included studies

| | | | | | | | | | |
|---|---|---|---|---|---|---|---|---|---|
| Kamm 2016 | Randomized controlled trial | Bangladesh | Rural | Households | Mother-child dyads | No theory reported | Handwashing with water; Handwashing with soap and water | **4** | Yes |
| Kang 2017 | Randomized controlled trial | Ethiopia | Rural | Households | Adults (Women only) | No theory reported | Handwashing - unspecified | **2** | Yes |
| Kapadia-Kundu 2014 | Randomized controlled trial | India | Rural | Schools | Mother-child dyads | Social Ecological Model | Handwashing with soap and water | **4** | Yes |
| Kariuki 2012 | Non-randomized quantitative | Kenya | Rural | Households | Children (Girls only) | No theory reported | Handwashing with soap and water | **3** | Yes |
| Karon 2017 | Non-randomized quantitative | Indonesia | Unspecified | Schools | Mother-child dyads | No theory reported | Handwashing with soap and water | **5** | Yes |
| Kitsanapun 2019 | Non-randomized quantitative | Thailand | Rural | Universities | Children (Girls and Boys) | Theory of Planned Behavior; Health Belief Model | Handwashing - unspecified | **3** | Yes |
| Koehn 2020 | Non-randomized quantitative | India | Unspecified | Households | Adults (Women and Men) | No theory reported | Handwashing with soap and water | **5** | No |
| Kumar 2018 | Mixed methods | India | Rural; Urban | Schools | Mother-child dyads | No theory reported | Handwashing - unspecified | **3** | Yes |
| Labović 2023 | Non-randomized quantitative | Montenegro | Urban | Workplaces | Children (Girls and Boys) | No theory reported | Handwashing with soap and water | **2** | No |
| Lange 2022 | Non-randomized quantitative | South Africa | Unspecified | Schools | Food workers | No theory reported | Handwashing - unspecified | **3** | Yes |
| Langford 2011 | Randomized controlled trial | Nepal | Unspecified | Households | Adults (Women and Men) | Theory of Planned Behavior | Handwashing with soap and water | **4** | Yes |
| Langford 2013 | Mixed methods | Nepal | Urban | Households | Mother-child dyads | Theory of Planned Behavior | Handwashing with soap and water | **5** | Yes |
| Lapinski 2013 | Randomized controlled trial | United States | Urban | Universities | Mother-child dyads | Theory of normative social | Handwashing - unspecified | **4** | Yes |

**S5 –** Overview of included studies

| | | | | | | | | | |
|---|---|---|---|---|---|---|---|---|---|
| | | | | | | behaviour; Focus Theory; Theory of reasoned action | | | |
| Lawson 2019 | Non-randomized quantitative | United Kingdom | Unspecified | Universities | Adults (Men only) | No theory reported | Handwashing with soap and water | **3** | No |
| Lee 2020 | Randomized controlled trial | China | Unspecified | Schools | Adults (Women and Men) | No theory reported | Handwashing with soap and water | **4** | Yes |
| Leventhal 2016 | Randomized controlled trial | India | Unspecified | Schools | Children (Girls and Boys) | COM-B | Handwashing with soap and water | **3** | Yes |
| Lhakhang 2015 | Randomized controlled trial | India | Rural | Universities | Children (Girls only) | COM-B; Health Action Process Approach | Handwashing with non-alcoholic antiseptics; Handwashing with water; Handwashing with soap and water | **3** | Yes |
| Liu 2019 | Non-randomized quantitative | China | Urban | Schools | Adults (Women and Men) | Health Belief Model | Handwashing - unspecified | **3** | Yes |
| Locks 2019 | Non-randomized quantitative | Democratic Republic of Congo | Peri-urban; Urban | Households | Adults (Women and Men) | No theory reported | Handwashing with soap and water | **5** | No |
| Lubna 2014 | Non-randomized quantitative | Bangladesh | Rural; Urban | Households | Children (Girls and Boys); Mother-child dyads | No theory reported | Handwashing with soap and water | **5** | Yes |
| Luby 2001 | Non-randomized quantitative | Pakistan | Rural | Households | Adults (Women only) | No theory reported | Handwashing with soap and water | **4** | Yes |
| Luby 2010 | Randomized controlled trial | Bangladesh | Urban | Households | General populations | Stages of change model | Handwashing with soap and water; Handwashing with non-alcoholic antiseptics | **4** | Yes |
| Machado 2018 | Randomized controlled trial | United States | Urban | Workplaces | Food workers | No theory reported | Handwashing with soap and water | **2** | Yes |

**S5 –** Overview of included studies

| | | | | | | | | | |
|---|---|---|---|---|---|---|---|---|---|
| Mackert 2013 | Non-randomized quantitative | United States | Unspecified | Universities | Adults (Women and Men) | Theory of Planned Behavior | Handwashing with water; Handwashing with soap and water | **4** | No |
| Mahfuza 2021 | Non-randomized quantitative | Bangladesh | Urban | Households | General populations | No theory reported | Handwashing - unspecified | **5** | Yes |
| Makata 2021 | Randomized controlled trial | Tanzania | Rural | Schools | Children (Girls and Boys) | No theory reported | Handwashing with soap and water | **4** | No |
| Malik 2022 | Non-randomized quantitative | Pakistan | Rural; Urban | Schools | Children (Boys only) | No theory reported | Handwashing - unspecified | **4** | Yes |
| Manaseki-Holland 2021 | Randomized controlled trial | The Gambia | Rural; Urban | Households | Adults (Women only) | | Handwashing with soap and water | **4** | Yes |
| Mane 2017 | Non-randomized quantitative | India | Rural | Schools | Children (Girls and Boys) | No theory reported | Handwashing with soap and water | **4** | Yes |
| Mathew 2018 | Non-randomized quantitative | India | Unspecified | Schools | Children (Girls and Boys) | Goal attainment model | Handwashing - unspecified | **3** | Yes |
| Maughan 2016 | Randomized controlled trial | United States | Rural | Markets | Adults (Women and Men) | No theory reported | Handwashing - unspecified | **3** | Yes |
| Mbakaya 2019 | Randomized controlled trial | Malawi | Urban | Schools | Children (Girls and Boys) | Social-cognitive learning theory | Handwashing with soap and water | **5** | Yes |
| McGuire-Wolfe 2012 | Non-randomized quantitative | United States | Unspecified | Workplaces | Non-food occupational workers | No theory reported | Handrubbing with alcohol-based rub | **3** | No |
| Mendes 2020 | Non-randomized quantitative | Brazil | Peri-urban | Childcare centers | Adults (Women only); Children (Girls and Boys) | Health Belief Model | Handwashing with soap and water | **3** | Yes |
| Mezaache 2021 | Mixed methods | France | Unspecified | Households | Adults (Women and Men) | COM-B | Handrubbing with alcohol-based rub | **3** | Yes |

**S5 –** Overview of included studies

| | | | | | | | | | |
|---|---|---|---|---|---|---|---|---|---|
| Mohamed 2019 | Non-randomized quantitative | Malaysia | Urban | Schools | Children (Girls and Boys) | No theory reported | Handwashing with soap and water | **2** | Yes |
| Moll 2007 | Non-randomized quantitative | Guatemala; Honduras; Nicaragua; El Salvador | Unspecified | Households | General populations | No theory reported | Handwashing - unspecified | **3** | Yes |
| Morse 2020 | Randomized controlled trial | Malawi | Peri-urban; Rural | Households | Children (Girls and Boys) | RANAS | Handwashing with soap and water | **2** | Yes |
| Mott 2007 | Randomized controlled trial | United States | Rural | Workplaces | Adults (Men only) | No theory reported | Handrubbing with alcohol-based rub | **2** | Yes |
| Nagapraveen 2016 | Non-randomized quantitative | India | Unspecified | Households | General populations | No theory reported | Handwashing with soap and water | **4** | Yes |
| Nair 2017 | Randomized controlled trial | India | Rural | Households | Mother-child dyads | No theory reported | Handwashing - unspecified | **2** | No |
| Naluonde 2019 | Randomized controlled trial | Zambia | Rural | Schools | Children (Girls and Boys) | No theory reported | Handwashing with soap and water | **4** | Yes |
| Nandrup-Bus 2009 | Randomized controlled trial | Denmark | Unspecified | Schools | Children (Girls and Boys) | No theory reported | Handwashing - unspecified | **2** | Yes |
| Newton-Lewis 2021 | Non-randomized quantitative | India | Unspecified | Households | Mother-child dyads | No theory reported | Handwashing - unspecified | **5** | No |
| NikRosmawati 2018 | Non-randomized quantitative | Malaysia | Unspecified | Schools | Food workers | Theory of Planned Behavior | Handwashing - unspecified | **5** | Yes |
| Nuhu 2019 | Randomized controlled trial | Bangladesh | Unspecified | Households | Mother-child dyads | No theory reported | Handwashing with soap and water | **4** | Yes |
| Öncü 2021 | Randomized controlled trial | Turkey | Urban | Schools | Children (Girls and Boys) | No theory reported | Handwashing with soap and water | **4** | Yes |
| Oruc 2021 | Non-randomized quantitative | Mozambique; Ethiopia; Uganda | Unspecified | Workplaces | Food workers | No theory reported | Handwashing - unspecified | **3** | Yes |
| Oswald 2014 | Non-randomized quantitative | Peru | Unspecified | Households | Adults (Women only) | No theory reported | Handwashing with water; Handwashing with soap and water | **3** | Yes |

**S5 –** Overview of included studies

| | | | | | | | | | |
|---|---|---|---|---|---|---|---|---|---|
| Ozcan 2020 | Non-randomized quantitative | Turkey | Peri-urban | Schools | Children (Girls and Boys) | No theory reported | Handwashing - unspecified | **4** | Yes |
| Patel 2012 | Non-randomized quantitative | Kenya | Rural | Schools | Children (Girls and Boys) | No theory reported | Handwashing with soap and water | **2** | Yes |
| Phuanukoonnon 2013 | Non-randomized quantitative | Papau New Guinea | Rural | Households | General populations | No theory reported | Handwashing with soap and water | **3** | Yes |
| Pickering 2013 | Randomized controlled trial | Kenya | Unspecified | Schools | Children (Girls and Boys) | No theory reported | Handwashing with soap and water; Handrubbing with alcohol-based rub | **2** | Yes |
| Pickering 2019 | Randomized controlled trial | Kenya | Urban | Households | General populations | No theory reported | Handwashing with soap and water | **4** | No |
| Pinfold 1990 | Randomized controlled trial | Thailand | Rural | Households | General populations | No theory reported | Handwashing - unspecified | **2** | Yes |
| Pokharel 2017 | Non-randomized quantitative | Armenia | Rural | Universities | Adults (Women and Men) | No theory reported | Handwashing - unspecified | **2** | Yes |
| Prado 2015 | Non-randomized quantitative | Brazil | Unspecified | Universities | Food workers | No theory reported | Handwashing - unspecified | **2** | No |
| Prasetyo 2022 | Non-randomized quantitative | Indonesia | Urban | Workplaces | Adults (Women and Men) | No theory reported | Handwashing with soap and water; Handwashing with water | **4** | Yes |
| Ram 2017 | Randomized controlled trial | Bangladesh | Unspecified | Households | Adults (Women only) | Heuristic model for teachable moments; Health Belief Model | Handwashing with water; Handwashing with soap and water | **4** | Yes |
| Ram 2020 | Randomized controlled trial | Bangladesh | Rural | Households | Mother-child dyads | Social Cognitive Theory; Theory of Planned Behavior; Health Belief Model | Handwashing with non-alcoholic antiseptics | **3** | Yes |
| Ray 2010 | Non-randomized quantitative | India | Rural | Households | General populations | No theory reported | Handwashing - unspecified | **3** | No |

**S5 –** Overview of included studies

| | | | | | | | | | |
|---|---|---|---|---|---|---|---|---|---|
| ReyesFernández 2015 | Randomized controlled trial | Costa Rica | Urban | Universities | Adults (Women and Men) | Health Action Process Approach | Handrubbing with alcohol-based rub | **4** | No |
| Riaz 2016 | Non-randomized quantitative | Bangladesh | Unspecified | Households | Adults (Women only) | No theory reported | Handwashing with soap and water; Hand drying | **3** | Yes |
| Rissman 2021 | Non-randomized quantitative | Malawi | Unspecified | Childcare centers | Adults (Women and Men); Children (Girls and Boys) | No theory reported | Handwashing with soap and water | **3** | Yes |
| Roberts 2008 | Non-randomized quantitative | United States | Rural | Workplaces | Food workers | No theory reported | Handwashing - unspecified | **2** | No |
| Roberts 2022 | Randomized controlled trial | United States | Unspecified | Schools | Food workers | Theory of Planned Behavior | Handwashing with soap and water | **1** | No |
| Rosen 2006 | Randomized controlled trial | Israel | Unspecified | Schools | Adults (Women and Men) | No theory reported | Handwashing with soap and water | **4** | Yes |
| Routh 2018 | Mixed methods | Malawi | Urban | Households | Children (Girls and Boys) | No theory reported | Handwashing with soap and water | **3** | Yes |
| Russo 2012 | Non-randomized quantitative | Malawi | Rural | Households | Adults (Women only) | No theory reported | Handwashing with soap and water | **4** | Yes |
| Rutter 2020 | Mixed methods | United Kingdom | Rural | Schools | Adults (Women and Men) | No theory reported | Handwashing with soap and water; Hand drying | **2** | Yes |
| Saboori 2013 | Randomized controlled trial | Kenya | Rural; Urban | Schools | Children (Girls and Boys) | No theory reported | Handwashing with soap and water; Handwashing with water | **3** | Yes |
| Samreen 2021 | Randomized controlled trial | Pakistan | Rural | Schools | Children (Girls and Boys) | No theory reported | Handwashing - unspecified | **2** | No |
| Sanders 2021 | Non-randomized quantitative | Tanzania; Uganda; Malawi | Urban | Households; Schools | Children (Girls and Boys) | No theory reported | Handwashing - unspecified | **3** | No |

**S5 –** Overview of included studies

| | | | | | | | | | |
|---|---|---|---|---|---|---|---|---|---|
| Sangalang 2021 | Randomized controlled trial | Philippines | Unspecified | Schools | General populations; Children (Girls and Boys) | No theory reported | Handwashing with soap and water | **4** | No |
| Sania 2017 | Mixed methods | Bangladesh | Urban | Households | Children (Girls and Boys) | IBM-WASH | Handwashing with soap and water | **5** | Yes |
| Schroeder 2016 | Non-randomized quantitative | United States | Rural | Workplaces | General populations | No theory reported | Handwashing with soap and water | **3** | Yes |
| Scott 2008 | Non-randomized quantitative | Ghana | Unspecified | Households | Food workers | No theory reported | Handwashing with soap and water | **2** | Yes |
| Shah 2021 | Non-randomized quantitative | India | Unspecified | Households | Mother-child dyads | No theory reported | Handwashing with soap and water | **3** | Yes |
| Shahar 2022 | Randomized controlled trial | Malaysia | Rural | Schools | Adults (Women and Men) | Health Belief Model | Handwashing with soap and water | **5** | No |
| Sheth 2004 | Non-randomized quantitative | India | Rural; Urban; Peri-urban | Schools | Children (Girls and Boys) | No theory reported | Handwashing with soap and water | **3** | Yes |
| Simiyu 2022 | Mixed methods | Kenya | Urban | Households | Mother-child dyads | Behavior-centered Design/Evo-Eco Model | Handwashing with soap and water | **2** | Yes |
| Simmerman 2011 | Randomized controlled trial | Thailand | Peri-urban | Households | Mother-child dyads | No theory reported | Handwashing with soap and water | **5** | No |
| Sneed 2015 | Non-randomized quantitative | United States | Urban | Households | General populations; Children (Girls and Boys) | No theory reported | Handwashing with soap and water | **3** | No |
| Snow 2008 | Randomized controlled trial | United States | Unspecified | Schools | Adults (Women and Men) | No theory reported | Handwashing with soap and water | **2** | Yes |
| Soares 2013 | Non-randomized quantitative | Portugal | Urban | Universities | Children (Girls and Boys) | No theory reported | Handwashing - unspecified | **3** | Yes |

**S5 –** Overview of included studies

| | | | | | | | | | |
|---|---|---|---|---|---|---|---|---|---|
| Sobel 2022 | Non-randomized quantitative | Not reported | Unspecified | Households | Food workers | No theory reported | Handwashing with soap and water | **3** | Yes |
| Solehati 2017 | Non-randomized quantitative | Indonesia | Unspecified | Schools | Mother-child dyads | No theory reported | Handwashing with soap and water | **3** | Yes |
| Stebbins 2010 | Randomized controlled trial | United States | Unspecified | Schools | Children (Girls and Boys) | No theory reported | Handwashing with soap and water; Handrubbing with alcohol-based rub | **2** | Yes |
| Stedman-Smith 2015 | Randomized controlled trial | United States | Urban | Workplaces | Children (Girls and Boys) | Theory of Planned Behavior | Handrubbing with alcohol-based rub | **4** | Yes |
| Strohbehn 2011 | Non-randomized quantitative | United States | Unspecified | Workplaces | Adults (Women and Men) | No theory reported | Handwashing with soap and water | **3** | No |
| Suen 2020 | Non-randomized quantitative | China | Unspecified | Schools | Food workers | No theory reported | Handrubbing with alcohol-based rub | **2** | Yes |
| Sutherland 2021 | Non-randomized quantitative | South Africa | Urban | Households | Children (Girls and Boys) | No theory reported | Handwashing with soap and water | **2** | Yes |
| Takanashi 2013 | Non-randomized quantitative | Vietnam | Urban | Households | General populations | No theory reported | Handwashing with soap and water | **3** | Yes |
| Taware 2018 | Non-randomized quantitative | India | Peri-urban | Schools | Mother-child dyads | Health Belief Model | Handwashing with soap and water | **3** | Yes |
| Thorseth 2021 | Mixed methods | Ethiopia | Urban | Internally displaced persons camps | Children (Girls and Boys) | No theory reported | Handwashing with soap and water | **4** | No |
| Tian 2019 | Mixed methods | China | Unspecified | Households | General populations | No theory reported | Handwashing - unspecified | **3** | Yes |
| Tidwell 2019 | Randomized controlled trial | India | Rural | Households | General populations | No theory reported | Handwashing with soap and water | **4** | Yes |
| Tidwell 2020 | Non-randomized quantitative | India | Urban | Schools | Adults (Women only) | No theory reported | Handwashing with soap and water | **4** | Yes |
| Topan 2020 | Non-randomized quantitative | Turkey | Unspecified | Schools | Children (Girls and Boys) | No theory reported | Handwashing with soap and water | **4** | Yes |

**S5 –** Overview of included studies

| | | | | | | | | | |
|---|---|---|---|---|---|---|---|---|---|
| Tousman 2007 | Non-randomized quantitative | United States | Unspecified | Schools | Children (Girls and Boys) | Health Behavior Change Model | Handwashing - unspecified | **2** | Yes |
| Tousman 2011 | Randomized controlled trial | United States | Unspecified | Not reported | Children (Girls and Boys) | No theory reported | Handwashing - unspecified | **4** | Yes |
| Umair 2019 | Non-randomized quantitative | Pakistan | Unspecified | Households | Adults (Women and Men) | No theory reported | Handwashing - unspecified | **3** | Yes |
| Underwood 2017 | Non-randomized quantitative | Nepal | Rural | Households | Adults (Women only) | Ideation by Cleland and Wilson | Handwashing with soap and water | **4** | Yes |
| Updegraff 2011 | Non-randomized quantitative | United States | Unspecified | Universities | Adults (Women and Men) | Health Belief Model; Theory of Planned Behavior | Handrubbing with alcohol-based rub | **3** | Yes |
| Vally 2019 | Non-randomized quantitative | The Philippines | Urban | Schools | General populations | No theory reported | Handwashing with soap and water; Handwashing with water; Handrubbing with alcohol-based rub | **2** | Yes |
| VazNery 2019 | Randomized controlled trial | Timor-Leste | Unspecified | Households | Children (Girls and Boys) | No theory reported | Handwashing with soap and water | **2** | No |
| Violant-Holz 2021 | Mixed methods | Spain | Rural | Schools | General populations | No theory reported | Handwashing - unspecified | **3** | Yes |
| Waterkeyn 2005 | Mixed methods | Zimbabwe | Unspecified | Community health clubs | Children (Girls and Boys) | No theory reported | Handwashing - unspecified | **4** | Yes |
| Watson 2019 | Non-randomized quantitative | Iraq | Rural | Internally displaced persons camps | Adults (Women and Men) | Behavior-centered Design/Evo-Eco Model | Handwashing with soap and water | **4** | Yes |
| Weijers 2020 | Non-randomized quantitative | The Netherlands | Unspecified | Markets | Children (Girls and Boys) | Dual Process Theory | Handrubbing with alcohol-based rub | **4** | No |
| White 2003 | Non-randomized quantitative | United States | Rural; Urban | Universities | General populations | No theory reported | Handrubbing with alcohol-based rub | **4** | Yes |

**S5 –** Overview of included studies

| | | | | | | | | | |
|---|---|---|---|---|---|---|---|---|---|
| Wichaidit 2019 | Randomized controlled trial | Kenya | Urban | Schools | Adults (Women and Men) | IBM-WASH | Handwashing with soap and water | **4** | No |
| Wichaidit 2019 | Randomized controlled trial | Bangladesh | Urban | Households | General populations | No theory reported | Handwashing with soap and water | **3** | Yes |
| Wilson 1993 | Non-randomized quantitative | Indonesia | Unspecified | Households | Children (Girls and Boys) | No theory reported | Handwashing with soap and water | **3** | Yes |
| Witt 2004 | Non-randomized quantitative | United States | Rural | Schools | Adults (Women only) | No theory reported | Handwashing with soap and water | **3** | Yes |
| Wong 2022 | Non-randomized quantitative | Malaysia | Unspecified | Schools | Children (Girls and Boys) | No theory reported | Handwashing with soap and water | **2** | Yes |
| Wu 2022 | Non-randomized quantitative | China | Unspecified | Schools | Food workers | Health Belief Model | Handwashing - unspecified | **3** | Yes |
| Yang 2017 | Randomized controlled trial | China | Unspecified | Markets | Adults (Women and Men) | No theory reported | Handwashing with soap and water | **3** | Yes |
| Yardley 2011 | Randomized controlled trial | England | Urban | Households | General populations | Theory of Planned Behavior | Handrubbing with alcohol-based rub; Handwashing with soap and water | **2** | Yes |
| Yeboah-Antwi 2019 | Non-randomized quantitative | Zambia | Unspecified | Households | Mother-child dyads | No theory reported | Handwashing - unspecified | **5** | Yes |
| York 2009 | Non-randomized quantitative | United States | Rural | Workplaces | Food workers | Theory of Planned Behavior | Handwashing - unspecified | **3** | No |
| Younie 2020 | Non-randomized quantitative | United Kingdom | Unspecified | Schools | Children (Girls and Boys) | COM-B | Handwashing with soap and water | **4** | Yes |
| Yu 2018 | Non-randomized quantitative | United States | Urban | Workplaces | Food workers | Organizational behavior modification model | Handwashing - unspecified | **3** | Yes |
| Zemichael 2020 | Non-randomized quantitative | Ethiopia | Unspecified | Households; Schools | Mother-child dyads | No theory reported | Handwashing - unspecified | **4** | Yes |

**S5 –** Overview of included studies

| | | | | | | | | | |
|---|---|---|---|---|---|---|---|---|---|
| Zhang 2013 | Non-randomized quantitative | Uganda | Rural | Schools | Children (Girls and Boys) | No theory reported | Handwashing with soap and water | **3** | Yes |
| Zhang 2021 | Randomized controlled trial | China | Rural; Urban; Peri-urban | Schools | Adults (Women and Men); Children (Girls and Boys) | No theory reported | Handwashing with soap and water | **3** | Yes |
| Zomer 2016 | Randomized controlled trial | The Netherlands | Urban | Schools | Adults (Women and Men) | No theory reported | Handwashing - unspecified | **4** | Yes |
