## Supplementary material for "Interventions to improve hand hygiene in community settings: A systematic review of theories, barriers and enablers, behavior change techniques, and hand hygiene station design features": S6

**S6** – Studies leveraged for each research question

**S6** – Studies leveraged for each research question

**Supplemental Table 6.** Included studies by research sub-question

| **Study ID** | A. Behavior change theories | B. Barriers and enablers | C. Behavior change techniques | D. Hand hygiene station designs | E. Hand hygiene station design adaptations | F. Frequency and intensity | G. Variation by population group, risk scenario, or time |
|---|---|---|---|---|---|---|---|
| Abbot 2012 | x | x | x | | | | x |
| Aboud 2011 | x | x | x | | | | x |
| Advaita 2021 | x | x | x | | | | x |
| Aibana 2013 | x | x | x | | | | x |
| Aiello 2012 | x | x | x | | | | x |
| Akina 2020 | x | x | x | x | | | x |
| Akuokoasibey 1994 | x | x | x | | | | x |
| Alam 1989 | x | x | x | x | | | x |
| Alexander 2012 | x | x | x | | | | x |
| Ali 2020 | x | x | x | | | | x |
| Alkon 2009 | x | x | x | | | | x |
| Almazan 2014 | x | x | x | | | | x |
| Amon-Tanoh 2021 | x | x | x | x | | x | x |
| Andrade 2019 | x | x | x | | | | x |
| Anu 2018 | x | x | x | | | | x |
| Appiah-Brempong 2020 | x | x | x | | | | x |
| Ar 2008 | x | x | x | | | | x |
| Ara 2022 | x | x | x | x | | | x |
| Aragie 2021 | x | x | x | x | | | x |
| Arbianingsih 2018 | x | x | x | | | | x |
| Arnold 2009 | x | x | x | | | | x |
| Ankan 2018 | x | x | x | | | | x |
| Ashtarian 2020 | x | x | x | | | | x |
| Ashutosh 2015 | x | x | x | | | | x |

**S6** – Studies leveraged for each research question

| | | | | | | | |
|---|---|---|---|---|---|---|---|
| Au 2010 | x | x | x | | | | x |
| Ãzyazıcıoğlu 2011 | x | x | x | | | | x |
| Bai 2022 | x | x | x | x | x | | x |
| Bajracharya 2003 | x | x | x | | | | x |
| Bickford 2017 | x | x | x | | | | x |
| Bieri 2013 | x | x | x | | | | x |
| Biran 2009 | x | x | x | | | | x |
| Biran 2014 | x | x | x | x | | | x |
| Biran 2020 | x | x | x | | | x | x |
| Biswas 2019 | x | x | x | | | | x |
| Blanton 2010 | x | x | x | x | | | x |
| Bosomprah 2016 | x | x | x | | | | x |
| Bowen 2013 | x | x | x | | | | x |
| Briceño 2017 | x | x | x | x | | | x |
| Briere 2012 | x | x | x | | | | x |
| Bulled 2017 | x | x | x | x | | | x |
| Burke 2016 | x | x | x | | | | x |
| Burns 2018 | x | x | x | | | | x |
| Cairncross 2005 | x | x | x | | | | x |
| Capps 2022 | x | x | x | | | | x |
| Carabin 1999 | x | x | x | | | | x |
| CardinaleLagomarsino 2017 | x | x | x | | | | x |
| Chard 2018 | x | x | x | x | | | x |
| Contzen 2013 | x | x | x | x | | | x |
| Costa 2019 | x | x | x | | | | x |
| Cowling 2009 | x | x | x | | | | x |
| Croghan 2008 | x | x | x | | | | x |
| Curtis 2001 | x | x | x | x | | | x |
| Davis 2011 | x | x | x | | | | x |

**S6** – Studies leveraged for each research question

| | | | | | | | |
|---|---|---|---|---|---|---|---|
| Davis 2013 | x | x | x | | x | | x |
| Ditai 2019 | x | x | x | | | | x |
| Dreibelbis 2016 | x | x | x | x | x | x | x |
| Duijster 2020 | x | x | x | x | | | x |
| Early 1998 | x | x | x | | | | x |
| Ebuehi 2010 | x | x | x | | | | x |
| Edward 2019 | x | x | x | | | | x |
| ErcanOruc 2020 | x | x | x | | | | x |
| Ercumen 2018 | x | x | x | x | | | x |
| Eun-Joo 2012 | x | x | x | | | | x |
| EvansJr 2009 | x | x | x | | | | x |
| Farhana 2022 | x | x | x | x | | | x |
| Ford 2014 | x | x | x | | x | | x |
| Freeman 2020 | x | x | x | | | x | x |
| Freeman 2022 | x | x | x | | | | x |
| Friedrich 2018 | x | x | x | x | | | x |
| Galiani 2016 | x | x | x | | | | x |
| Gautam 2017 | x | x | x | | | | x |
| Gedamu 2022 | x | x | x | | | | x |
| Geller 1980 | x | x | x | | | | x |
| George 2017 | x | x | x | x | | | x |
| Gimaiyo 2019 | x | x | x | | | | x |
| Goel 2019 | x | x | x | | | | x |
| Goel 2020 | x | x | x | | | | x |
| Grace 2012 | x | x | x | | | | x |
| Greene 2012 | x | x | x | x | | | x |
| Greenland 2016 | x | x | x | | | | x |
| Grover 2018 | x | x | x | x | x | | x |
| Guo 2018 | x | x | x | | | | x |
| Hanson 2020 | x | x | x | | | | x |

**S6** – Studies leveraged for each research question

| | | | | | | | |
|---|---|---|---|---|---|---|---|
| Her 2019 | x | x | x | | | | x |
| Hetherington 2017 | x | x | x | | | | x |
| Huang 2021 | x | x | x | x | x | | x |
| Hurley 2021 | x | x | x | | | | x |
| Hussam 2022 | x | x | x | | | | x |
| Jafree 2023 | x | x | x | | | | x |
| Jagals 2004 | x | x | x | | | | x |
| Jetha 2021 | x | x | x | | | | x |
| Jinadu 2007 | x | x | x | | | | x |
| Johnson 2003 | x | x | x | | | | x |
| Judah 2009 | x | x | x | | | | x |
| Kaewchana 2012 | x | x | x | | | | x |
| Kajjura 2019 | x | x | x | | | | x |
| Kamm 2016 | x | x | x | x | | | x |
| Kang 2017 | x | x | x | | | | x |
| Kapadia-Kundu 2014 | x | x | x | | | | x |
| Kariuki 2012 | x | x | x | | | | x |
| Karon 2017 | x | x | x | x | | | x |
| Kitsanapun 2019 | x | x | x | | | | x |
| Koehn 2020 | x | x | x | | | | x |
| Kumar 2018 | x | x | x | | | | x |
| Labović 2023 | x | x | x | | | | x |
| Lange 2022 | x | x | x | | | | x |
| Langford 2011 | x | x | x | | | | x |
| Langford 2013 | x | x | x | | | | x |
| Lapinski 2013 | x | x | x | | x | | x |
| Lawson 2019 | x | x | x | | | | x |
| Lee 2020 | x | x | x | | | | x |
| Leventhal 2016 | x | x | x | | | | x |
| Lhakhang 2015 | x | x | x | | | | x |

**S6** – Studies leveraged for each research question

| | | | | | | | |
|---|---|---|---|---|---|---|---|
| Liu 2019 | x | x | x | | | | x |
| Locks 2019 | x | x | x | | | | x |
| Lubna 2014 | x | x | x | | | | x |
| Luby 2001 | x | x | x | x | | | x |
| Luby 2010 | x | x | x | | | | x |
| Machado 2018 | x | x | x | | | | x |
| Mackert 2013 | x | x | x | | | | x |
| Mahfuza 2021 | x | x | x | | | | x |
| Makata 2021 | x | x | x | x | | | x |
| Malik 2022 | x | x | x | | | | x |
| Manaseki-Holland 2021 | x | x | x | | | | x |
| Mane 2017 | x | x | x | | | | x |
| Mathew 2018 | x | x | x | | | | x |
| Maughan 2016 | x | x | x | | | | x |
| Mbakaya 2019 | x | x | x | x | | | x |
| McGuire-Wolfe 2012 | x | x | x | | | | x |
| Mendes 2020 | x | x | x | | | | x |
| Mezaache 2021 | x | x | x | | | | x |
| Mohamed 2019 | x | x | x | | | | x |
| Moll 2007 | x | x | x | | | | x |
| Morse 2020 | x | x | x | x | | | x |
| Mott 2007 | x | x | x | | | | x |
| Nagapraveen 2016 | x | x | x | | | | x |
| Nair 2017 | x | x | x | | | | x |
| Naluonde 2019 | x | x | x | | | | x |
| Nandrup-Bus 2009 | x | x | x | | | | x |
| Newton-Lewis 2021 | x | x | x | | | | x |
| NikRosmawati 2018 | x | x | x | | | | x |
| Nuhu 2019 | x | x | x | | | x | x |
| Öncü 2021 | x | x | x | | | | x |

**S6** – Studies leveraged for each research question

| | | | | | | | |
|---|---|---|---|---|---|---|---|
| Oruc 2021 | x | x | x | | | | x |
| Oswald 2014 | x | x | x | x | | | x |
| Ozcan 2020 | x | x | x | | | | x |
| Patel 2012 | x | x | x | x | | | x |
| Phuanukoonnon 2013 | x | x | x | | | | x |
| Pickering 2013 | x | x | x | x | | | x |
| Pickering 2019 | x | x | x | x | | | x |
| Pinfold 1990 | x | x | x | x | | | x |
| Pokharel 2017 | x | x | x | | | | x |
| Prado 2015 | x | x | x | | | | x |
| Prasetyo 2022 | x | x | x | x | x | | x |
| Ram 2017 | x | x | x | x | | | x |
| Ram 2020 | x | x | x | | | | x |
| Ray 2010 | x | x | x | | | | x |
| ReyesFernández 2015 | x | x | x | | | | x |
| Riaz 2016 | x | x | x | | | | x |
| Rissman 2021 | x | x | x | | | | x |
| Roberts 2008 | x | x | x | | | | x |
| Roberts 2022 | x | x | x | | | | x |
| Rosen 2006 | x | x | x | | | | x |
| Routh 2018 | x | x | x | | | | x |
| Russo 2012 | x | x | x | | | | x |
| Rutter 2020 | x | x | x | | | | x |
| Saboori 2013 | x | x | x | | | | x |
| Samreen 2021 | x | x | x | | | | x |
| Sanders 2021 | x | x | x | x | | | x |
| Sangalang 2021 | x | x | x | x | | x | x |
| Sania 2017 | x | x | x | x | | | x |
| Schroeder 2016 | x | x | x | | | | x |
| Scott 2008 | x | x | x | | | | x |

**S6** – Studies leveraged for each research question

| | | | | | | | |
|---|---|---|---|---|---|---|---|
| Shah 2021 | x | x | x | | | | x |
| Shahar 2022 | x | x | x | | | | x |
| Sheth 2004 | x | x | x | | | | x |
| Simiyu 2022 | x | x | x | | | | x |
| Simmerman 2011 | x | x | x | | | | x |
| Sneed 2015 | x | x | x | | | | x |
| Snow 2008 | x | x | x | | | | x |
| Soares 2013 | x | x | x | | | | x |
| Sobel 2022 | x | x | x | | | | x |
| Solehati 2017 | x | x | x | | | | x |
| Stebbins 2010 | x | x | x | | | | x |
| Stedman-Smith 2015 | x | x | x | | | | x |
| Strohbehn 2011 | x | x | x | | | | x |
| Suen 2020 | x | x | x | | | | x |
| Sutherland 2021 | x | x | x | x | | | x |
| Takanashi 2013 | x | x | x | | | | x |
| Taware 2018 | x | x | x | | | | x |
| Thorseth 2021 | x | x | x | x | x | | x |
| Tian 2019 | x | x | x | | | | x |
| Tidwell 2019 | x | x | x | | | | x |
| Tidwell 2020 | x | x | x | | | | x |
| Topan 2020 | x | x | x | | | | x |
| Tousman 2007 | x | x | x | | | | x |
| Tousman 2011 | x | x | x | | | | x |
| Umair 2019 | x | x | x | | | | x |
| Underwood 2017 | x | x | x | | | | x |
| Updegraff 2011 | x | x | x | | | | x |
| Vally 2019 | x | x | x | x | | | x |
| VazNery 2019 | x | x | x | | | | x |
| Violant-Holz 2021 | x | x | x | | | | x |

**S6** – Studies leveraged for each research question

| | | | | | | | |
|---|---|---|---|---|---|---|---|
| Waterkeyn 2005 | x | x | x | | | | x |
| Watson 2019 | x | x | x | | | | x |
| Weijers 2020 | x | x | x | | x | | x |
| White 2003 | x | x | x | | | | x |
| Wichaidit 2019 | x | x | x | x | | | x |
| Wichaidit 2019 | x | x | x | x | | | x |
| Wilson 1993 | x | x | x | | | | x |
| Witt 2004 | x | x | x | | | | x |
| Wong 2022 | x | x | x | | | | x |
| Wu 2022 | x | x | x | | | | x |
| Yang 2017 | x | x | x | x | | | x |
| Yardley 2011 | x | x | x | | | | x |
| Yeboah-Antwi 2019 | x | x | x | | | | x |
| York 2009 | x | x | x | | | | x |
| Younie 2020 | x | x | x | | | | x |
| Yu 2018 | x | x | x | | | | x |
| Zemichael 2020 | x | x | x | x | | | x |
| Zhang 2013 | x | x | x | x | | | x |
| Zhang 2021 | x | x | x | | | | x |
| Zomer 2016 | x | x | x | | | | x |
