## Supplementary material for "Interventions to improve hand hygiene in community settings: A systematic review of theories, barriers and enablers, behavior change techniques, and hand hygiene station design features": S7

**S7** – MMAT Assessment

**S7** – MMAT Assessment

### *Supplemental file 7:* Quality Appraisal of all Included Articles Using the Mixed Methods Appraisal Tool

| Study | Final Score | Qualitative Score | Quantitative Score | Mixed Methods Score | Criteria from the Mixed Methods Appraisal Tool[1] | | | | | | | | | | | | | | | | | | | | | | | | |
|---|---|---|---|---|---|---|---|---|---|---|---|---|---|---|---|---|---|---|---|---|---|---|---|---|---|---|---|---|---|
| | | | | | KEY<br>Individual criteria scores can be either 0 (did not meet criteria) or 1 (met criteria); cells that are shaded in gray indicate that a criterion was not applicable to the study type.<br><br>Qualitative and quantitative studies were assessed using the five-criteria questionnaire. Mixed methods studies were assessed using the relevant independent questionnaires for qualitative and quantitative work and a five criteria questionnaire for mixed methods; the lowest of the three scores was used as the quality score. Possible scores are 0–5 across study types (5 is the best).<br><br>† Indicates that the MMAT was deemed inappropriate for quality appraisal of the article. | | | | | | | | | | | | | | | | | | | | | | | | |
| | | | | | **1.1** | **1.2** | **1.3** | **1.4** | **1.5** | **2.1** | **2.2** | **2.3** | **2.4** | **2.5** | **3.1** | **3.2** | **3.3** | **3.4** | **3.5** | **4.1** | **4.2** | **4.3** | **4.4** | **4.5** | **5.1** | **5.2** | **5.3** | **5.4** | **5.5** |
| Abbot 2012 | **2** | | 2 | | | | | | | | | | | | 0 | 1 | 0 | 0 | 1 | | | | | | | | | | |
| Aboud 2011 | **4** | | 4 | | | | | | | 1 | 0 | 1 | 1 | 1 | | | | | | | | | | | | | | | |
| Adam 2014 | **4** | | 4 | | | | | | | 1 | 1 | 1 | 0 | 1 | | | | | | | | | | | | | | | |
| Advaita 2021 | **3** | | 3 | | | | | | | | | | | | 0 | 1 | 1 | 0 | 1 | | | | | | | | | | |
| Aibana 2013 | **4** | | 4 | | | | | | | | | | | | 1 | 1 | 0 | 1 | 1 | | | | | | | | | | |
| Aiello 2012 | **4** | | 4 | | | | | | | 1 | 0 | 1 | 1 | 1 | | | | | | | | | | | | | | | |
| Akina 2020 | **3** | | 3 | | | | | | | 0 | 0 | 1 | 1 | 1 | | | | | | | | | | | | | | | |
| Akuokoasibey 1994 | **3** | | 3 | | | | | | | | | | | | 0 | 1 | 1 | 0 | 1 | | | | | | | | | | |
| Alam 1989 | **4** | | 4 | | | | | | | | | | | | 1 | 1 | 1 | 0 | 1 | | | | | | | | | | |
| Alexander 2012 | **4** | | 4 | | | | | | | | | | | | 1 | 1 | 1 | 0 | 1 | | | | | | | | | | |
| Ali 2020 | **3** | | 3 | | | | | | | | | | | | 0 | 1 | 1 | 0 | 1 | | | | | | | | | | |
| Alkon 2009 | **4** | | 4 | | | | | | | 1 | 1 | 1 | 0 | 1 | | | | | | | | | | | | | | | |
| Almazan 2014 | **3** | | 3 | | | | | | | | | | | | 0 | 1 | 1 | 0 | 1 | | | | | | | | | | |
| Amon-Tanoh 2021 | **5** | | 5 | | | | | | | 1 | 1 | 1 | 1 | 1 | | | | | | | | | | | | | | | |
| Andrade 2019 | **4** | | 4 | | | | | | | | | | | | 1 | 1 | 1 | 0 | 1 | | | | | | | | | | |
| Anu 2018 | **3** | 3 | 3 | 1 | 1 | 1 | 0 | 1 | 0 | | | | | | 0 | 1 | 1 | 0 | 1 | | | | | | 0 | 1 | 0 | 0 | 0 |
| Appiah-Brempong 2020 | **5** | | 5 | | | | | | | 1 | 1 | 1 | 1 | 1 | | | | | | | | | | | | | | | |

**S7** – MMAT Assessment

| | | | | | | | | | | | | | | | | | | | |
|---|---|---|---|---|---|---|---|---|---|---|---|---|---|---|---|---|---|---|---|
| Ar 2008 | **3** | | 3 | | | | | | | | | | | | 0 | 1 | 1 | 0 | 1 |
| Ara 2022 | **4** | | 4 | | | | | | | 1 | 1 | 1 | 1 | 0 | | | | | |
| Aragie 2021 | **4** | | 4 | | | | | | | 1 | 1 | 1 | 0 | 1 | | | | | |
| Arbianingsih 2018 | **2** | | 2 | | | | | | | | | | | | 0 | 1 | 0 | 0 | 1 |
| Arnold 2009 | **4** | | 4 | | | | | | | | | | | | 1 | 1 | 1 | 0 | 1 |
| Ankan 2018 | **1** | | 1 | | | | | | | 0 | 0 | 1 | 0 | 0 | | | | | |
| Ashtarian 2020 | **3** | | 3 | | | | | | | 1 | 0 | 1 | 0 | 1 | | | | | |
| Ashutosh 2015 | **3** | | 3 | | | | | | | | | | | | 0 | 1 | 1 | 0 | 1 |
| Au 2010 | **3** | | 3 | | | | | | | | | | | | 0 | 1 | 1 | 0 | 1 |
| Ãzyazıcıoğlu 2011 | **3** | | 3 | | | | | | | | | | | | 0 | 1 | 1 | 0 | 1 |
| Bai 2022 | **3** | | 3 | | | | | | | | | | | | 0 | 1 | 1 | 0 | 1 |
| Bajracharya 2003 | **2** | | 2 | | | | | | | | | | | | 0 | 1 | 0 | 0 | 1 |
| Bickford 2017 | **4** | | 4 | | | | | | | | | | | | 1 | 1 | 1 | 0 | 1 |
| Bieri 2013 | **3** | | 3 | | | | | | | 1 | 1 | 1 | 0 | 0 | | | | | |
| Biran 2009 | **4** | | 4 | | | | | | | 1 | 1 | 1 | 0 | 1 | | | | | |
| Biran 2020 | **4** | | 4 | | | | | | | 0 | 1 | 1 | 1 | 1 | | | | | |
| Biswas 2019 | **2** | | 2 | | | | | | | 1 | 1 | 0 | 0 | 0 | | | | | |
| Blanton 2010 | **3** | | 3 | | | | | | | | | | | | 1 | 1 | 0 | 0 | 1 |
| Bosomprah 2016 | **3** | | 3 | | | | | | | | | | | | 1 | 1 | 0 | 0 | 1 |
| Bowen 2013 | **2** | | 2 | | | | | | | 0 | 0 | 1 | 0 | 1 | | | | | |
| Briceño 2017 | **3** | | 3 | | | | | | | 1 | 0 | 1 | 0 | 1 | | | | | |
| Briere 2012 | **4** | | 4 | | | | | | | | | | | | 1 | 1 | 1 | 0 | 1 |
| Bulled 2017 | **2** | | 2 | | | | | | | | | | | | 0 | 1 | 0 | 0 | 1 |
| Burke 2016 | **4** | | 4 | | | | | | | | | | | | 1 | 1 | 1 | 0 | 1 |
| Burns 2018 | **3** | | 3 | | | | | | | 1 | 1 | 0 | 0 | 1 | | | | | |
| Cairncross 2005 | **2** | | 2 | | | | | | | | | | | | 0 | 1 | 0 | 0 | 1 |
| Capps 2022 | **3** | | 3 | | | | | | | | | | | | 0 | 1 | 0 | 1 | 1 |
| Carabin 1999 | **3** | | 3 | | | | | | | 0 | 1 | 1 | 0 | 1 | | | | | |

**S7** – MMAT Assessment

| | | | | | | | | | | | | | | | | | | | | | | | | | | | | | |
|---|---|---|---|---|---|---|---|---|---|---|---|---|---|---|---|---|---|---|---|---|---|---|---|---|---|---|---|---|---|
| CardinaleLagomarsino 2017 | **3** | | 3 | | | | | | | 1 | 0 | 1 | 0 | 1 | | | | | | | | | | | | | | | |
| Chard 2018 | **4** | | 4 | | | | | | | 1 | 0 | 1 | 1 | 1 | | | | | | | | | | | | | | | |
| Contzen 2013 | **3** | | 3 | | | | | | | | | | | | 0 | 1 | 1 | 0 | 1 | | | | | | | | | | |
| Costa 2019 | **3** | | 3 | | | | | | | | | | | | 0 | 1 | 1 | 0 | 1 | | | | | | | | | | |
| Cowling 2009 | **3** | | 3 | | | | | | | 1 | 1 | 1 | 0 | 0 | | | | | | | | | | | | | | | |
| Croghan 2008 | **2** | | 2 | | | | | | | | | | | | 0 | 1 | 0 | 0 | 1 | | | | | | | | | | |
| Curtis 2001 | **3** | | 3 | | | | | | | | | | | | 0 | 1 | 1 | 0 | 1 | | | | | | | | | | |
| Davis 2011 | **3** | | 3 | | | | | | | 1 | 0 | 1 | 0 | 1 | | | | | | | | | | | | | | | |
| Davis 2013 | **4** | | 4 | | | | | | | | | | | | 1 | 1 | 1 | 0 | 1 | | | | | | | | | | |
| Ditai 2019 | **4** | | 4 | | | | | | | 1 | 1 | 1 | 0 | 1 | | | | | | | | | | | | | | | |
| Duijster 2020 | **2** | | 2 | | | | | | | 1 | 0 | 0 | 0 | 1 | | | | | | | | | | | | | | | |
| Early 1998 | **3** | | 3 | | | | | | | | | | | | 1 | 1 | 0 | 0 | 1 | | | | | | | | | | |
| Ebuehi 2010 | **3** | | 3 | | | | | | | | | | | | 0 | 1 | 1 | 0 | 1 | | | | | | | | | | |
| Edward 2019 | **5** | | 5 | | | | | | | | | | | | 1 | 1 | 1 | 1 | 1 | | | | | | | | | | |
| ErcanOruc 2020 | **3** | | 3 | | | | | | | | | | | | 0 | 1 | 1 | 0 | 1 | | | | | | | | | | |
| Ercumen 2018 | **3** | | 3 | | | | | | | 1 | 0 | 1 | 0 | 1 | | | | | | | | | | | | | | | |
| Eun-Joo 2012 | **4** | | 4 | | | | | | | | | | | | 1 | 1 | 1 | 0 | 1 | | | | | | | | | | |
| EvansJr 2009 | **3** | | 3 | | | | | | | | | | | | 0 | 1 | 0 | 1 | 1 | | | | | | | | | | |
| Farhana 2022 | **3** | 5 | 3 | 5 | 1 | 1 | 1 | 1 | 1 | | | | | | 0 | 1 | 1 | 0 | 1 | | | | | | 1 | 1 | 1 | 1 | 1 |
| Ford 2014 | **2** | | 2 | | | | | | | | | | | | 0 | 1 | 0 | 0 | 1 | | | | | | | | | | |
| Freeman 2020 | **3** | | 3 | | | | | | | 1 | 1 | 0 | 0 | 1 | | | | | | | | | | | | | | | |
| Freeman 2022 | **4** | | 4 | | | | | | | 1 | 1 | 1 | 0 | 1 | | | | | | | | | | | | | | | |
| Friedrich 2018 | **3** | | 3 | | | | | | | 1 | 1 | 1 | 0 | 0 | | | | | | | | | | | | | | | |
| Galiani 2016 | **4** | | 4 | | | | | | | 1 | 1 | 1 | 0 | 1 | | | | | | | | | | | | | | | |
| Gautam 2017 | **5** | | 5 | | | | | | | 1 | 1 | 1 | 1 | 1 | | | | | | | | | | | | | | | |
| Gedamu 2022 | **4** | | 4 | | | | | | | | | | | | 1 | 1 | 1 | 0 | 1 | | | | | | | | | | |
| Geller 1980 | **3** | | 3 | | | | | | | | | | | | 0 | 1 | 1 | 0 | 1 | | | | | | | | | | |
| George 2017 | **5** | | 5 | | | | | | | 1 | 1 | 1 | 1 | 1 | | | | | | | | | | | | | | | |

**S7** – MMAT Assessment

| | | | | | | | | | | | | | | | | | | | | | | | | | | | | | |
|---|---|---|---|---|---|---|---|---|---|---|---|---|---|---|---|---|---|---|---|---|---|---|---|---|---|---|---|---|---|
| Gimaiyo 2019 | **5** | | 5 | | | | | | | 1 | 1 | 1 | 1 | 1 | | | | | | | | | | | | | | | |
| Goel 2019 | **3** | | 3 | | | | | | | | | | | | 0 | 1 | 1 | 0 | 1 | | | | | | | | | | |
| Goel 2020 | **4** | | 4 | | | | | | | | | | | | 1 | 1 | 1 | 0 | 1 | | | | | | | | | | |
| Grace 2012 | **4** | | 4 | | | | | | | | | | | | 1 | 1 | 1 | 0 | 1 | | | | | | | | | | |
| Greene 2012 | **3** | | 3 | | | | | | | 1 | 0 | 1 | 0 | 1 | | | | | | | | | | | | | | | |
| Greenland 2016 | **4** | | 4 | | | | | | | 1 | 0 | 1 | 1 | 1 | | | | | | | | | | | | | | | |
| Grover 2018 | **2** | | 2 | | | | | | | 0 | 0 | 1 | 0 | 1 | | | | | | | | | | | | | | | |
| Grover 2018 | **3** | | 3 | | | | | | | 1 | 0 | 1 | 0 | 1 | | | | | | | | | | | | | | | |
| Guo 2018 | **2** | | 2 | | | | | | | 1 | 0 | 0 | 0 | 1 | | | | | | | | | | | | | | | |
| Hanson 2020 | **2** | | 2 | | | | | | | | | | | | 0 | 1 | 0 | 1 | 0 | | | | | | | | | | |
| Her 2019 | **2** | | 2 | | | | | | | | | | | | 0 | 1 | 0 | 0 | 1 | | | | | | | | | | |
| Hetherington 2017 | **4** | 5 | 4 | 5 | 1 | 1 | 1 | 1 | 1 | | | | | | 1 | 1 | 1 | 0 | 1 | | | | | | 1 | 1 | 1 | 1 | 1 |
| Huang 2021 | **4** | 5 | 4 | 5 | 1 | 1 | 1 | 1 | 1 | 1 | 1 | 1 | 0 | 1 | | | | | | | | | | | 1 | 1 | 1 | 1 | 1 |
| Hurley 2021 | **2** | | 2 | | | | | | | | | | | | 0 | 1 | 0 | 0 | 1 | | | | | | | | | | |
| Hussam 2022 | **3** | | 3 | | | | | | | 1 | 0 | 1 | 0 | 1 | | | | | | | | | | | | | | | |
| Jafree 2023 | **3** | | 3 | | | | | | | | | | | | 0 | 1 | 1 | 0 | 1 | | | | | | | | | | |
| Jagals 2004 | **2** | | 2 | | | | | | | | | | | | 0 | 1 | 1 | 0 | 0 | | | | | | | | | | |
| Jetha 2021 | **4** | | 4 | | | | | | | 1 | 1 | 1 | 0 | 1 | | | | | | | | | | | | | | | |
| Jinadu 2007 | **3** | | 3 | | | | | | | | | | | | 0 | 1 | 1 | 0 | 1 | | | | | | | | | | |
| Johnson 2003 | **3** | | 3 | | | | | | | | | | | | 0 | 1 | 1 | 0 | 1 | | | | | | | | | | |
| Judah 2009 | **4** | | 4 | | | | | | | | | | | | 1 | 1 | 1 | 0 | 1 | | | | | | | | | | |
| Kaewchana 2012 | **5** | | 5 | | | | | | | 1 | 1 | 1 | 1 | 1 | | | | | | | | | | | | | | | |
| Kajjura 2019 | **3** | | 3 | | | | | | | | | | | | 0 | 1 | 1 | 0 | 1 | | | | | | | | | | |
| Kamm 2016 | **4** | | 4 | | | | | | | 1 | 1 | 1 | 0 | 1 | | | | | | | | | | | | | | | |
| Kang 2017 | **2** | | 2 | | | | | | | 1 | 0 | 0 | 0 | 1 | | | | | | | | | | | | | | | |
| Kapadia-Kundu 2014 | **4** | | 4 | | | | | | | 1 | 1 | 1 | 0 | 1 | | | | | | | | | | | | | | | |
| Kariuki 2012 | **3** | | 3 | | | | | | | | | | | | 0 | 1 | 1 | 0 | 1 | | | | | | | | | | |
| Karon 2017 | **5** | | 5 | | | | | | | | | | | | 1 | 1 | 1 | 1 | 1 | | | | | | | | | | |

**S7** – MMAT Assessment

| | | | | | | | | | | | | | | | | | | | | | | | | | | | | | |
|---|---|---|---|---|---|---|---|---|---|---|---|---|---|---|---|---|---|---|---|---|---|---|---|---|---|---|---|---|---|
| Kitsanapun 2019 | **3** | | 3 | | | | | | | | | | | | 0 | 1 | 1 | 0 | 1 | | | | | | | | | | |
| Koehn 2020 | **5** | | 5 | | | | | | | | | | | | 1 | 1 | 1 | 1 | 1 | | | | | | | | | | |
| Kumar 2018 | **3** | 3 | 3 | 3 | 1 | 1 | 0 | 0 | 1 | | | | | | 0 | 1 | 1 | 0 | 1 | | | | | | 1 | 1 | 0 | 0 | 1 |
| Labović 2023 | **2** | | 2 | | | | | | | | | | | | 0 | 1 | 0 | 0 | 1 | | | | | | | | | | |
| Lange 2022 | **3** | | 3 | | | | | | | | | | | | 1 | 1 | 0 | 0 | 1 | | | | | | | | | | |
| Langford 2011 | **4** | | 4 | | | | | | | 1 | 1 | 1 | 0 | 1 | | | | | | | | | | | | | | | |
| Langford 2013 | **5** | 5 | 5 | 5 | 1 | 1 | 1 | 1 | 1 | 1 | 1 | 1 | 1 | 1 | | | | | | | | | | | 1 | 1 | 1 | 1 | 1 |
| Lapinski 2013 | **4** | | 4 | | | | | | | 1 | 0 | 1 | 1 | 1 | | | | | | | | | | | | | | | |
| Lawson 2019 | **3** | | 3 | | | | | | | | | | | | 1 | 1 | 0 | 0 | 1 | | | | | | | | | | |
| Lee 2020 | **4** | | 4 | | | | | | | 1 | 0 | 1 | 1 | 1 | | | | | | | | | | | | | | | |
| Leventhal 2016 | **3** | | 3 | | | | | | | 1 | 0 | 1 | 0 | 1 | | | | | | | | | | | | | | | |
| Lhakhang 2015 | **3** | | 3 | | | | | | | 1 | 0 | 0 | 1 | 1 | | | | | | | | | | | | | | | |
| Liu 2019 | **3** | | 3 | | | | | | | | | | | | 0 | 1 | 1 | 0 | 1 | | | | | | | | | | |
| Locks 2019 | **5** | | 5 | | | | | | | | | | | | 1 | 1 | 1 | 1 | 1 | | | | | | | | | | |
| Lubna 2014 | **5** | | 5 | | | | | | | | | | | | 1 | 1 | 1 | 1 | 1 | | | | | | | | | | |
| Luby 2001 | **4** | | 4 | | | | | | | | | | | | 1 | 1 | 1 | 0 | 1 | | | | | | | | | | |
| Luby 2010 | **4** | | 4 | | | | | | | 1 | 1 | 1 | 0 | 1 | | | | | | | | | | | | | | | |
| Machado 2018 | **2** | | 2 | | | | | | | 1 | 0 | 0 | 0 | 1 | | | | | | | | | | | | | | | |
| Mackert 2013 | **4** | | 4 | | | | | | | | | | | | 0 | 1 | 1 | 1 | 1 | | | | | | | | | | |
| Mahfuza 2021 | **5** | | 5 | | | | | | | | | | | | 1 | 1 | 1 | 1 | 1 | | | | | | | | | | |
| Makata 2021 | **4** | | 4 | | | | | | | 1 | 1 | 0 | 1 | 1 | | | | | | | | | | | | | | | |
| Malik 2022 | **4** | | 4 | | | | | | | | | | | | 1 | 1 | 1 | 0 | 1 | | | | | | | | | | |
| Manaseki-Holland 2021 | **4** | | 4 | | | | | | | 1 | 1 | 1 | 0 | 1 | | | | | | | | | | | | | | | |
| Mane 2017 | **4** | | 4 | | | | | | | | | | | | 1 | 1 | 1 | 0 | 1 | | | | | | | | | | |
| Mathew 2018 | **3** | | 3 | | | | | | | | | | | | 1 | 1 | 1 | 0 | 0 | | | | | | | | | | |
| Maughan 2016 | **3** | | 3 | | | | | | | 1 | 0 | 1 | 0 | 1 | | | | | | | | | | | | | | | |
| Mbakaya 2019 | **5** | | 5 | | | | | | | 1 | 1 | 1 | 1 | 1 | | | | | | | | | | | | | | | |
| McGuire-Wolfe 2012 | **3** | | 3 | | | | | | | | | | | | 1 | 1 | 0 | 0 | 1 | | | | | | | | | | |

**S7** – MMAT Assessment

| | | | | | | | | | | | | | | | | | | | | | | | | | | | | | |
|---|---|---|---|---|---|---|---|---|---|---|---|---|---|---|---|---|---|---|---|---|---|---|---|---|---|---|---|---|---|
| Mendes 2020 | **3** | | 3 | | | | | | | | | | | | 0 | 1 | 1 | 0 | 1 | | | | | | | | | | |
| Mezaache 2021 | **3** | 5 | 3 | 2 | 1 | 1 | 1 | 1 | 1 | | | | | | 1 | 1 | 0 | 0 | 1 | | | | | | 1 | 0 | 1 | 0 | 0 |
| Mohamed 2019 | **2** | | 2 | | | | | | | | | | | | 0 | 1 | 1 | 0 | | | | | | | | | | | |
| Moll 2007 | **3** | | 3 | | | | | | | | | | | | 0 | 1 | 1 | 0 | 1 | | | | | | | | | | |
| Morse 2020 | **2** | | 2 | | | | | | | 0 | 1 | 0 | 0 | 1 | | | | | | | | | | | | | | | |
| Mott 2007 | **2** | | 2 | | | | | | | 0 | 0 | 1 | 0 | 1 | | | | | | | | | | | | | | | |
| Nagapraveen 2016 | **4** | | 4 | | | | | | | | | | | | 1 | 1 | 1 | 0 | 1 | | | | | | | | | | |
| Nair 2017 | **2** | | 2 | | | | | | | 1 | 1 | 0 | 0 | 0 | | | | | | | | | | | | | | | |
| Naluonde 2019 | **4** | | 4 | | | | | | | 1 | 1 | 0 | 1 | 1 | | | | | | | | | | | | | | | |
| Nandrup-Bus 2009 | **2** | | 2 | | | | | | | 0 | 0 | 1 | 0 | 1 | | | | | | | | | | | | | | | |
| Newton-Lewis 2021 | **5** | | 5 | | | | | | | | | | | | 1 | 1 | 1 | 1 | 1 | | | | | | | | | | |
| NikRosmawati 2018 | **5** | | 5 | | | | | | | | | | | | 1 | 1 | 1 | 1 | 1 | | | | | | | | | | |
| Nuhu 2019 | **4** | | 4 | | | | | | | 1 | 1 | 1 | 0 | 1 | | | | | | | | | | | | | | | |
| Öncü 2021 | **4** | | 4 | | | | | | | 1 | 0 | 1 | 1 | 1 | | | | | | | | | | | | | | | |
| Oruc 2021 | **3** | | 3 | | | | | | | | | | | | 0 | 1 | 1 | 0 | 1 | | | | | | | | | | |
| Oswald 2014 | **3** | | 3 | | | | | | | | | | | | 0 | 1 | 0 | 1 | 1 | | | | | | | | | | |
| Ozcan 2020 | **4** | | 4 | | | | | | | | | | | | 1 | 1 | 1 | 0 | 1 | | | | | | | | | | |
| Patel 2012 | **2** | | 2 | | | | | | | | | | | | 0 | 1 | 0 | 0 | 1 | | | | | | | | | | |
| Phuanukoonnon 2013 | **3** | | 3 | | | | | | | | | | | | 0 | 1 | 1 | 0 | 1 | | | | | | | | | | |
| Pickering 2013 | **2** | | 2 | | | | | | | 0 | 0 | 1 | 0 | 1 | | | | | | | | | | | | | | | |
| Pickering 2019 | **4** | | 4 | | | | | | | 1 | 1 | 1 | 0 | 1 | | | | | | | | | | | | | | | |
| Pinfold 1990 | **2** | | 2 | | | | | | | 0 | 1 | 0 | 0 | 1 | | | | | | | | | | | | | | | |
| Pokharel 2017 | **2** | | 2 | | | | | | | | | | | | 0 | 1 | 0 | 0 | 1 | | | | | | | | | | |
| Prado 2015 | **2** | | 2 | | | | | | | | | | | | 0 | 1 | 0 | 0 | 1 | | | | | | | | | | |
| Prasetyo 2022 | **4** | | 4 | | | | | | | | | | | | 1 | 1 | 1 | 0 | 1 | | | | | | | | | | |
| Ram 2017 | **4** | | 4 | | | | | | | 1 | 1 | 1 | 0 | 1 | | | | | | | | | | | | | | | |
| Ram 2020 | **3** | | 3 | | | | | | | 1 | 1 | 1 | 0 | 0 | | | | | | | | | | | | | | | |
| Ray 2010 | **3** | | 3 | | | | | | | | | | | | 0 | 1 | 1 | 0 | 1 | | | | | | | | | | |

**S7** – MMAT Assessment

| | | | | | | | | | | | | | | | | | | | | | | | | | | | | | |
|---|---|---|---|---|---|---|---|---|---|---|---|---|---|---|---|---|---|---|---|---|---|---|---|---|---|---|---|---|---|
| ReyesFernández 2015 | **4** | | 4 | | | | | | | 1 | 1 | 0 | 1 | 1 | | | | | | | | | | | | | | | |
| Riaz 2016 | **3** | | 3 | | | | | | | | | | | | 0 | 1 | 1 | 0 | 1 | | | | | | | | | | |
| Rissman 2021 | **3** | | 3 | | | | | | | | | | | | 0 | 1 | 1 | 0 | 1 | | | | | | | | | | |
| Roberts 2008 | **2** | | 2 | | | | | | | | | | | | 0 | 1 | 0 | 0 | 1 | | | | | | | | | | |
| Roberts 2022 | **1** | | 1 | | | | | | | 0 | 0 | 0 | 0 | 1 | | | | | | | | | | | | | | | |
| Rosen 2006 | **4** | | 4 | | | | | | | 1 | 1 | 0 | 1 | 1 | | | | | | | | | | | | | | | |
| Routh 2018 | **3** | 5 | 3 | 4 | 1 | 1 | 1 | 1 | 1 | | | | | | 0 | 1 | 1 | 0 | 1 | | | | | | 1 | 1 | 0 | 1 | 1 |
| Russo 2012 | **4** | | 4 | | | | | | | | | | | | 1 | 1 | 1 | 0 | 1 | | | | | | | | | | |
| Rutter 2020 | **2** | 5 | 2 | 4 | 1 | 1 | 1 | 1 | 1 | | | | | | 0 | 1 | 0 | 0 | 1 | | | | | | 1 | 1 | 1 | 0 | 1 |
| Saboori 2013 | **3** | | 3 | | | | | | | 0 | 1 | 1 | 0 | 1 | | | | | | | | | | | | | | | |
| Samreen 2021 | **2** | | 2 | | | | | | | 0 | 0 | 1 | 0 | 1 | | | | | | | | | | | | | | | |
| Sanders 2021 | **3** | | 3 | | | | | | | | | | | | 0 | 1 | 1 | 0 | 1 | | | | | | | | | | |
| Sangalang 2021 | **4** | | 4 | | | | | | | 1 | 1 | 1 | 0 | 1 | | | | | | | | | | | | | | | |
| Sania 2017 | **5** | 5 | 5 | 3 | 1 | 1 | 1 | 1 | 1 | | | | | | 1 | 1 | 1 | 1 | 1 | | | | | | 1 | 0 | 1 | 1 | 0 |
| Schroeder 2016 | **3** | | 3 | | | | | | | | | | | | 1 | 1 | 0 | 0 | 1 | | | | | | | | | | |
| Scott 2008 | **2** | | 2 | | | | | | | | | | | | 0 | 1 | 0 | 0 | 1 | | | | | | | | | | |
| Shah 2021 | **3** | | 3 | | | | | | | | | | | | 0 | 1 | 1 | 0 | 1 | | | | | | | | | | |
| Shahar 2022 | **5** | | 5 | | | | | | | 1 | 1 | 1 | 1 | 1 | | | | | | | | | | | | | | | |
| Sheth 2004 | **3** | | 3 | | | | | | | | | | | | 0 | 1 | 1 | 0 | 1 | | | | | | | | | | |
| Simiyu 2022 | **2** | 5 | 2 | 2 | 1 | 1 | 1 | 1 | 1 | 0 | 0 | 1 | 0 | 1 | | | | | | | | | | | 1 | 1 | 0 | 0 | 0 |
| Simmerman 2011 | **5** | | 5 | | | | | | | 1 | 1 | 1 | 1 | 1 | | | | | | | | | | | | | | | |
| Sneed 2015 | **3** | | 3 | | | | | | | | | | | | 0 | 1 | 1 | 0 | 1 | | | | | | | | | | |
| Snow 2008 | **2** | | 2 | | | | | | | 0 | 0 | 1 | 0 | 1 | | | | | | | | | | | | | | | |
| Soares 2013 | **3** | | 3 | | | | | | | | | | | | 0 | 1 | 1 | 0 | 1 | | | | | | | | | | |
| Sobel 2022 | **3** | | 3 | | | | | | | | | | | | 0 | 1 | 1 | 0 | 1 | | | | | | | | | | |
| Solehati 2017 | **3** | | 3 | | | | | | | | | | | | 0 | 1 | 1 | 0 | 1 | | | | | | | | | | |
| Stebbins 2010 | **2** | | 2 | | | | | | | 0 | 0 | 1 | 0 | 1 | | | | | | | | | | | | | | | |
| Stedman-Smith 2015 | **4** | | 4 | | | | | | | 1 | 1 | 1 | 0 | 1 | | | | | | | | | | | | | | | |

**S7** – MMAT Assessment

| | | | | | | | | | | | | | | | | | | | | | | | | | | | | | |
|---|---|---|---|---|---|---|---|---|---|---|---|---|---|---|---|---|---|---|---|---|---|---|---|---|---|---|---|---|---|
| Strohbehn 2011 | 3 | | 3 | | | | | | | | | | | | 0 | 1 | 1 | 0 | 1 | | | | | | | | | | |
| Suen 2020 | 2 | | 2 | | | | | | | | | | | | 0 | 1 | 0 | 0 | 1 | | | | | | | | | | |
| Sutherland 2021 | 2 | | 2 | | | | | | | | | | | | 0 | 1 | 0 | 0 | 1 | | | | | | | | | | |
| Takanashi 2013 | 3 | | 3 | | | | | | | | | | | | 0 | 1 | 1 | 0 | 1 | | | | | | | | | | |
| Taware 2018 | 3 | | 3 | | | | | | | | | | | | 1 | 1 | 0 | 0 | 1 | | | | | | | | | | |
| Thorseth 2021 | 4 | 5 | 4 | 5 | 1 | 1 | 1 | 1 | 1 | | | | | | 1 | 1 | 1 | 0 | 1 | | | | | | 1 | 1 | 1 | 1 | 1 |
| Tian 2019 | 3 | 2 | 3 | 0 | 1 | 1 | 0 | 0 | 0 | | | | | | 0 | 1 | 1 | 0 | 1 | | | | | | 0 | 0 | 0 | 0 | 0 |
| Tidwell 2019 | 4 | | 4 | | | | | | | 0 | 1 | 1 | 1 | 1 | | | | | | | | | | | | | | | |
| Tidwell 2020 | 4 | | 4 | | | | | | | | | | | | 1 | 1 | 1 | 0 | 1 | | | | | | | | | | |
| Topan 2020 | 4 | | 4 | | | | | | | | | | | | 1 | 1 | 1 | 0 | 1 | | | | | | | | | | |
| Tousman 2007 | 2 | | 2 | | | | | | | | | | | | 0 | 1 | 0 | 0 | 1 | | | | | | | | | | |
| Tousman 2011 | 4 | | 4 | | | | | | | 1 | 1 | 1 | 0 | 1 | | | | | | | | | | | | | | | |
| Umair 2019 | 3 | | 3 | | | | | | | | | | | | 0 | 1 | 1 | 0 | 1 | | | | | | | | | | |
| Underwood 2017 | 4 | | 4 | | | | | | | | | | | | 1 | 1 | 1 | 0 | 1 | | | | | | | | | | |
| Updegraff 2011 | 3 | | 3 | | | | | | | | | | | | 0 | 1 | 1 | 0 | 1 | | | | | | | | | | |
| Vally 2019 | 2 | | 2 | | | | | | | | | | | | 0 | 1 | 0 | 1 | 0 | | | | | | | | | | |
| VazNery 2019 | 2 | | 2 | | | | | | | 0 | 1 | 0 | 0 | 1 | | | | | | | | | | | | | | | |
| Violant-Holz 2021 | 3 | 5 | 3 | 5 | 1 | 1 | 1 | 1 | 1 | | | | | | 0 | 1 | 1 | 0 | 1 | | | | | | 1 | 1 | 1 | 1 | 1 |
| Waterkeyn 2005 | 4 | 5 | 4 | 5 | 1 | 1 | 1 | 1 | 1 | | | | | | 1 | 1 | 1 | 0 | 1 | | | | | | 1 | 1 | 1 | 1 | 1 |
| Watson 2019 | 4 | | 4 | | | | | | | | | | | | 0 | 1 | 1 | 1 | 1 | | | | | | | | | | |
| Weijers 2020 | 4 | | 4 | | | | | | | | | | | | 1 | 1 | 1 | 0 | 1 | | | | | | | | | | |
| White 2003 | 4 | | 4 | | | | | | | | | | | | 1 | 1 | 1 | 0 | 1 | | | | | | | | | | |
| Wichaidit 2019 | 4 | | 4 | | | | | | | 1 | 1 | 1 | 0 | 1 | | | | | | | | | | | | | | | |
| Wichaidit 2019 | 3 | | 3 | | | | | | | 1 | 0 | 1 | 0 | 1 | | | | | | | | | | | | | | | |
| Wilson 1993 | 3 | | 3 | | | | | | | | | | | | 0 | 1 | 1 | 0 | 1 | | | | | | | | | | |
| Witt 2004 | 3 | | 3 | | | | | | | | | | | | 0 | 1 | 1 | 0 | 1 | | | | | | | | | | |
| Wong 2022 | 2 | | 2 | | | | | | | | | | | | 0 | 1 | 1 | 0 | | | | | | | | | | | |
| Wu 2022 | 3 | | 3 | | | | | | | | | | | | 0 | 1 | 1 | 0 | 1 | | | | | | | | | | |

**S7** – MMAT Assessment

| | | | | | | | | | | | | | | | | | | | |
|---|---|---|---|---|---|---|---|---|---|---|---|---|---|---|---|---|---|---|---|
| Yang 2017 | **3** | | 3 | | | | | | | 0 | 1 | 1 | 0 | 1 | | | | | |
| Yardley 2011 | **2** | | 2 | | | | | | | 0 | 1 | 0 | 0 | 1 | | | | | |
| Yeboah-Antwi 2019 | **5** | | 5 | | | | | | | | | | | | 1 | 1 | 1 | 1 | 1 |
| York 2009 | **3** | | 3 | | | | | | | | | | | | 0 | 1 | 1 | 0 | 1 |
| Younie 2020 | **4** | | 4 | | | | | | | | | | | | 1 | 1 | 1 | 0 | 1 |
| Yu 2018 | **3** | | 3 | | | | | | | | | | | | 0 | 1 | 1 | 0 | 1 |
| Zemichael 2020 | **4** | | 4 | | | | | | | | | | | | 1 | 1 | 1 | 0 | 1 |
| Zhang 2013 | **3** | | 3 | | | | | | | | | | | | 0 | 1 | 1 | 0 | 1 |
| Zhang 2021 | **3** | | 3 | | | | | | | 1 | 1 | 1 | 0 | 0 | | | | | |
| Zomer 2016 | **4** | | 4 | | | | | | | 1 | 1 | 1 | 0 | 1 | | | | | |

[1]Hong, Q.N., Pluye, P., et al. Mixed Methods Appraisal Tool (MMAT) Version 2018 User Guide. McGill Department of Family Medicine. 2018.

**Criteria from the MMAT:**
1.1 Is the qualitative approach appropriate to answer the research question?
1.2 Are the qualitative data collection methods adequate to address the research question?
1.3 Are the findings adequately derived from the data?
1.4 Is the interpretation of results sufficiently substantiated by data?
1.5 Is there coherence between qualitative data sources, collection, analysis and interpretation?
2.1 Is randomization appropriately performed?
2.2 Are the groups comparable at baseline?
2.3 Are there complete outcome data?
2.4 Are outcome assessors blinded to the intervention provided?
2.5 Did the participants adhere to the assigned intervention?
3.1 Are the participants representative of the target population?
3.2 Are measurements appropriate regarding both the outcome and intervention (or exposure)?
3.3 Are there complete outcome data?
3.4 Are the confounders accounted for in the design and analysis?
3.5 During the study period, is the intervention administered (or exposure occurred) as intended?
4.1 Is the sampling strategy relevant to address the research question?
4.2 Is the sample representative of the target population?
4.3 Are the measurements appropriate?
4.4 Is the risk of nonresponse bias low?
4.5 Is the statistical analysis appropriate to answer the research question?
5.1 Is there an adequate rationale for using a mixed methods design to address the research question?

5.2 Are the different components of the study effectively integrated to answer the research question?
5.3 Are the outputs of the integration of qualitative and quantitative components adequately interpreted?
5.4 Are divergences and inconsistencies between quantitative and qualitative results adequately addressed?
5.5 Do the different components of the study adhere to the quality criteria of each tradition of the methods involved?
