## Supplementary material for "Interventions to improve hand hygiene in community settings: A systematic review of theories, barriers and enablers, behavior change techniques, and hand hygiene station design features": S8

**S8** – Theory table

**S8** – Theory table

***Supplementary File 8*****.** Theories or models reported to be used in included studies (N=223).

| **Theory** | **Total n (%)** | **Effective** n (%)** |
|---|---|---|
| **Total Studies** | **223** | **183 (82.1)** |
| **Study did not report using theory for intervention design** | **160 (71.8)** | **131 (81.9)** |
| **Study did report using theory for intervention design*** | **63 (28.2)** | **52 (82.5)** |
| Theory of Planned Behavior | 14 (22.2) | 10 (71.4) |
| Health Belief Model | 11 (17.5) | 10 (90.9) |
| Behaviour Centered Design/Evo-Eco Model | 7 (11.1) | 5 (71.4) |
| RANAS | 6 (9.5) | 5 (83.3) |
| COM-B | 6 (9.5) | 5 (83.3) |
| IBM-WASH | 5 (7.9) | 4 (80.0) |
| Health Access Process Approach | 2 (3.2) | 1 (50.0) |
| Social Ecological Model | 2 (3.2) | 2 (100.0) |
| Social-cognitive Learning Theory | 2 (3.2) | 2 (100.0) |
| Stages of Change Model | 2 (3.2) | 2 (100.0) |
| Theory of Normative Social Behavior | 2 (3.2) | 2 (100.0) |
| Theory of Reasoned Action | 2 (3.2) | 2 (100.0) |
| Behavior Place Theory | 1 (1.6) | 1 (100.0) |
| Bloom's Taxonomy of Learning Theory | 1 (1.6) | 1 (100.0) |
| Choice-architecture Approach | 1 (1.6) | 1 (100.0) |
| Ideation Theory | 1 (1.6) | 1 (100.0) |
| Dual Process Theory | 1 (1.6) | 0 (0.0) |
| Ecological Theory of Health Promotion | 1 (1.6) | 1 (100.0) |
| Health Behavior Change Model | 1 (1.6) | 1 (100.0) |
| Heuristic Model for Teachable Moments | 1 (1.6) | 1 (100.0) |
| Organization Behavior Modification Model | 1 (1.6) | 1 (100.0) |
| Focus Theory | 1 (1.6) | 1 (100.0) |
| Nudge Theory | 1 (1.6) | 1 (100.0) |
| Goal Attainment Model | 1 (1.6) | 1 (100.0) |
| Social Cognitive Theory | 1 (1.6) | 1 (100.0) |

***Note:*** **Multiple theories could have been reported per study so the total number of theories (74) is greater than the total number of studies that report using theory (63).; **Effectiveness is determined if authors reported that the intervention was effective at improving hand hygiene outcomes*
