## Supplementary material for "Interventions to improve hand hygiene in community settings: A systematic review of theories, barriers and enablers, behavior change techniques, and hand hygiene station design features": S9

**S9 –** Theory bar chart

**S9 –** Theory bar chart

**Supplemental Figure 9**. Studies that reported theory across settings and by year

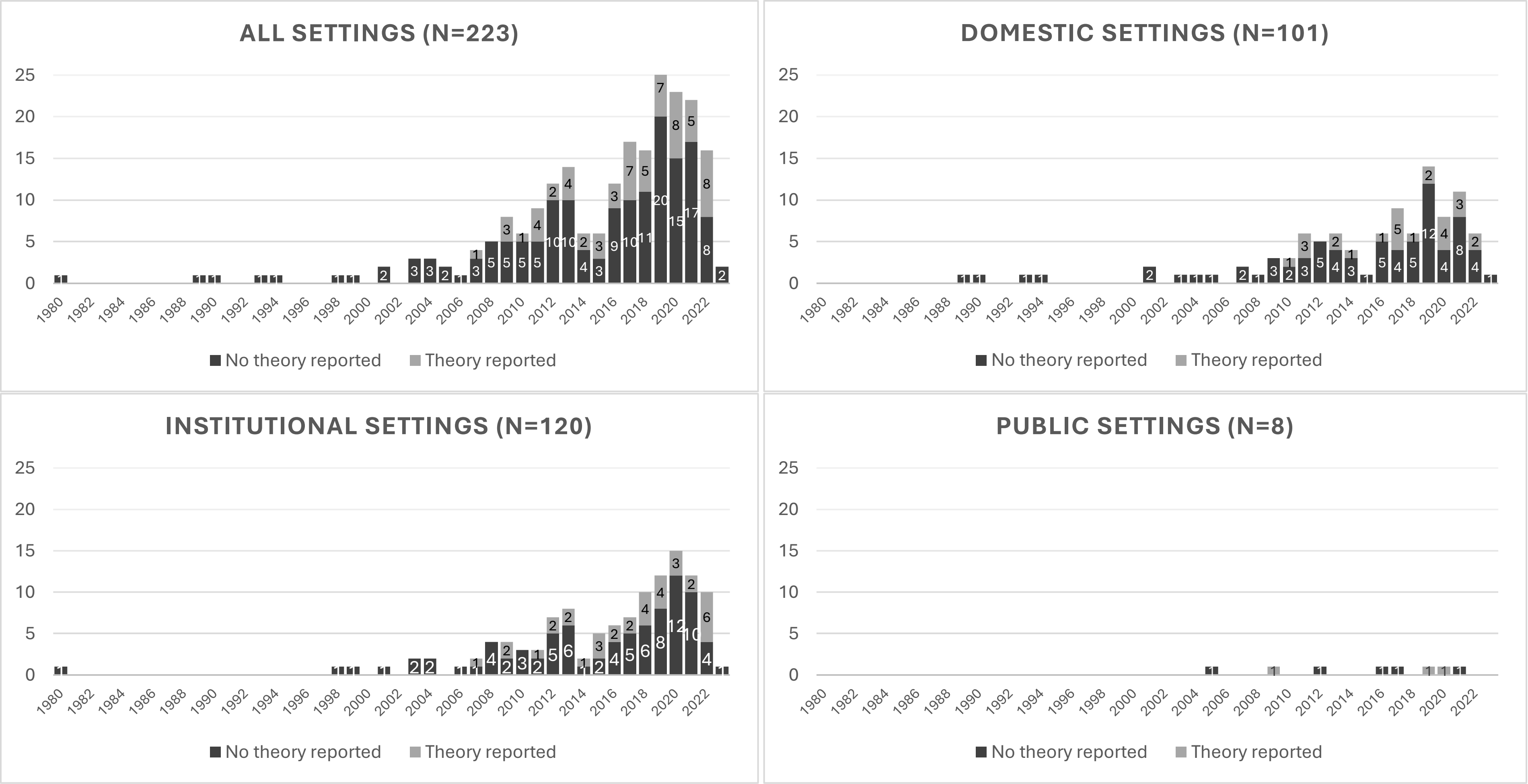


* Six studies were counted twice since they specifically contained both domestic and institutional settings, making up the total number of studies across all settings (N=223). This overlap accounts for the discrepancy between the total number of studies across all settings and the sum of studies reported within individual settings.
