## Supplementary material for "Interventions to improve hand hygiene in community settings: A systematic review of theories, barriers and enablers, behavior change techniques, and hand hygiene station design features": S10

**S10** – Examples of addressed vs reported misalignment

*Supplementary File 10*. **Representative examples of reported and addressed classifications**

| COM-B Component | Reported but not addressed | Addressed but not reported |
|---|---|---|
| **Capability** | | |
| *Physical* | N/A | N/A |
| *Psychological* | N/A | Briceño (2017) used multimedia messages and community health worker visits to improve knowledge of hand hygiene behavior but did not report **action knowledge** as a barrier |
| **Opportunity** | | |
| *Physical* | Sania (2017) reported **water availability** as an enabler but the intervention provided soap and a handwashing station without water provision | Biran (2014) provided a handwashing station but did not report any barriers or enablers related to **handwashing infrastructure** |
| *Social* | Capps (2022) reported **social pressure** as a barrier and enabler but the intervention involved placing posters near hand sanitizer dispensers | Edward (2019) used visits from community health workers to improve hand hygiene behavior but did not report **network support** or **role modeling** as enablers |
| **Motivation** | | |
| *Automatic* | Huang (2021) reported **internal motivation** as a barrier but provided a handwashing station | Tousman (2011) used diaries and journals to stimulate hand hygiene as a habit but did not report **internal motivation** as a barrier |
| *Reflective* | Abbot (2012) reported **time prioritization** as a barrier but used multimedia videos to instruct on how to perform hand hygiene behavior | Greene (2012) implemented a school-based education program that discussed the health consequences from inadequate hand hygiene but did not report **disease risk** as an enabler |
