## Supplementary material for "Interventions to improve hand hygiene in community settings: A systematic review of theories, barriers and enablers, behavior change techniques, and hand hygiene station design features": S11

**S11** – Frequency of BCT packages

**S11** – Frequency of BCT packages

***Supplementary Table 11*****.** Frequency of identified behavior change technique (BCT) packages and frequency of BCTs per package with effectiveness across included studies (N=223)

| Number of BCT packages per study | Number and % of studies | Effectiveness[2] n (%) | Number of BCTs per package | Number and % of studies[1] | Effectiveness[2] n (%) |
|---|---|---|---|---|---|
| 1 package | 100 (44.8%) | 78 (78.0%) | 1 BCT | 71 (31.8%) | 57 (80.3%) |
| 2 packages | 62 (27.8%) | 53 (85.5%) | 2 BCTs | 167 (74.9%) | 143 (85.6%) |
| 3 packages | 38 (17.0%) | 33 (86.8%) | 3 BCTs | 109 (48.8%) | 87 (79.8%) |
| 4 packages | 17 (7.6%) | 13 (76.5%) | | | |
| 5 packages | 5 (2.2%) | 5 (100.0%) | | | |
| 6 packages | 1 (0.5%) | 1 (100.0%) | | | |
| Total Packages[1] | 437 | | | | |

***Note:*** [1]Adds up to more than the total number of studies included as studies could have evaluated packages with different numbers of BCTs; [2]*Effectiveness is determined if authors reported that the intervention was effective at improving hand hygiene outcomes*
