## Supplementary material for "Interventions to improve hand hygiene in community settings: A systematic review of theories, barriers and enablers, behavior change techniques, and hand hygiene station design features": S12

**S12** – IA, BCTs and activities

**S12** – IA, BCTs and activities

**Supplemental Table 12.** Intervention functions, behavior change techniques and intervention activities leveraged, by study

| Study ID | Intervention function | Behavior Change Technique | Intervention activity |
|---|---|---|---|
| Abbot 2012 | Training | Information on how to perform the behavior | Multimedia - Videos |
| | Education; Enablement | Action planning; Information on others; approval; Goal setting (behavior) | Group discussions |
| Aboud 2011 | Enablement; Training | Demonstration of behavior; Social support (practical) | Training - Adults/Community-based |
| | Education; Training | Instruction on how to perform the behavior; Information about health consequences | Education - Adults/Community-based |
| Advaita 2021 | Education; Training | Instruction on how to perform the behavior; Information about health consequences | Education - Adults/Community-based |
| | Enablement; Training | Demonstration of behavior; Social support (practical) | Training - Adults/Community-based |
| Aibana 2013 | Education; Training | Instruction on how to perform the behavior; Information about health consequences | Education - Adults/Community-based |
| Aiello 2012 | Environmental Restructuring | Adding objects to the environment | Alcohol rub provision |
| Akina 2020 | Education; Training; Environmental Restructuring | Information about health consequences; Instruction on how to perform the behavior; Adding objects to the environment | Multimedia - Posters |
| | Training | Demonstration of behavior; Feedback on behavior; Instruction on how to perform the behavior | School-based Training |
| | Environmental Restructuring | Restructuring the physical environmnent; Adding objects to the environment | HW station |
| | Environmental Restructuring | Adding objects to the environment | Soap provision |
| Akuokoasibey 1994 | Education; Training | Instruction on how to perform the behavior; Information about health consequences | Education - Adults/Community-based |
| Alam 1989 | Environmental Restructuring | Restructuring the physical environmnent; Adding objects to the environment | HW station |
| | Education; Training | Instruction on how to perform the behavior; Information about health consequences | Education - Adults/Community-based |

**S12** – IA, BCTs and activities

| | | | |
|---|---|---|---|
| Alexander 2012 | Education; Training; Environmental Restructuring | Information about health consequences; Instruction on how to perform the behavior; Adding objects to the environment | Multimedia - Multiple |
| | Education; Training | Instruction on how to perform the behavior; Information about health consequences | Education - Adults/Community-based |
| Ali 2020 | Environmental Restructuring | Adding objects to the environment | Soap provision |
| Alkon 2009 | Education; Training; Enablement | Demonstration of behavior; Information on health consequences; Social support (practical) | CHW visits |
| Almazan 2014 | Education; Training | Instruction on how to perform the behavior; Information about health consequences | Education - Adults/Community-based |
| Amon-Tanoh 2021 | Training | Information on how to perform the behavior | Multimedia - Videos |
| | Enablement; Training | Demonstration of behavior; Social support (practical) | Training - Adults/Community-based |
| | Environmental Restructuring | Prompts/cues; Adding objects to the environment | Soap provision |
| | Training | Information on how to perform the behavior | Multimedia - Videos |
| | Environmental Restructuring | Restructuring the physical environmnent; Adding objects to the environment | HW station |
| Andrade 2019 | Enablement; Training | Demonstration of behavior; Social support (practical) | Training - Adults/Community-based |
| Ankan 2018 | Training | Information on how to perform the behavior | Multimedia - Videos |
| | Education; Training | Instruction on how to perform the behavior; Information about health consequences | School-based Education |
| Anu 2018 | Environmental Restructuring | Adding objects to the environment | Soap provision |
| | Education; Training | Instruction on how to perform the behavior; Information about health consequences | Education - Adults/Community-based |
| | Education; Training | Instruction on how to perform the behavior; Information about health consequences | Multimedia - radio |
| Appiah-Brempong 2020 | Training | Demonstration of behavior; Feedback on behavior; Instruction on how to perform the behavior | School-based Training |
| | Education; Training | Instruction on how to perform the behavior; Information about health consequences | School-based Education |

**S12** – IA, BCTs and activities

| | | | |
|---|---|---|---|
| Ar 2008 | Training | Demonstration of behavior; Feedback on behavior; Instruction on how to perform the behavior | School-based Training |
| Ara 2022 | Education; Training | Instruction on how to perform the behavior; Information about health consequences | Education - Adults/Community-based |
| | Environmental Restructuring | Adding objects to the environment | Soap provision |
| | Environmental Restructuring | Restructuring the physical environmnent; Adding objects to the environment | HW station |
| Aragie 2021 | Education; Training | Instruction on how to perform the behavior; Information about health consequences | Education - Adults/Community-based |
| | Enablement; Training | Demonstration of behavior; Social support (practical) | Training - Adults/Community-based |
| | Education; Training; Enablement | Demonstration of behavior; Information on health consequences; Social support (practical) | CHW visits |
| | Environmental Restructuring | Restructuring the physical environmnent; Adding objects to the environment | HW station |
| | Environmental Restructuring | Adding objects to the environment | Soap provision |
| Arbianingsih 2018 | Education; Training; Incentivization | Information about health consequences; Demonstration of behavior; Material incentive | Games or competitions |
| Arnold 2009 | Enablement; Training | Demonstration of behavior; Social support (practical) | Training - Adults/Community-based |
| Ashtarian 2020 | Training | Demonstration of behavior; Feedback on behavior; Instruction on how to perform the behavior | School-based Training |
| | Education; Training | Instruction on how to perform the behavior; Information about health consequences | School-based Education |
| Ashutosh 2015 | Education; Training | Instruction on how to perform the behavior; Information about health consequences | School-based Education |
| Au 2010 | Education; Training | Instruction on how to perform the behavior; Information about health consequences | School-based Education |
| | Education; Training | Instruction on how to perform the behavior; Information about health consequences | Plays, skits or songs |
| Ãzyazıcıoğlu 2011 | Education; Training | Instruction on how to perform the behavior; Information about health consequences | School-based Education |

**S12** – IA, BCTs and activities

| | | | |
|---|---|---|---|
| Bai 2022 | Environmental Restructuring | Restructuring the physical environmnent; Adding objects to the environment | HW station |
| Bajracharya 2003 | Education; Training; Environmental Restructuring | Information about health consequences; Instruction on how to perform the behavior; Adding objects to the environment | Multimedia - Multiple |
| | Education; Training; Enablement | Demonstration of behavior; Information on health consequences; Social support (practical) | CHW visits |
| | Enablement; Training | Demonstration of behavior; Social support (practical) | Training - Adults/Community-based |
| Bickford 2017 | Training | Demonstration of behavior; Feedback on behavior; Instruction on how to perform the behavior | School-based Training |
| | Training | Information on how to perform the behavior | Multimedia - Videos |
| Bieri 2013 | Education; Training | Instruction on how to perform the behavior; Information about health consequences | Multimedia - Unspecified |
| | Education; Training | Instruction on how to perform the behavior; Information about health consequences | School-based Education |
| Biran 2009 | Education; Training | Instruction on how to perform the behavior; Information about health consequences | Education - Adults/Community-based |
| | Education; Training; Environmental Restructuring | Information about health consequences; Instruction on how to perform the behavior; Adding objects to the environment | Multimedia - Art/flipcharts |
| | Incentivization | Incentive (outcome) | Other incentives provided |
| Biran 2014 | Enablement; Training | Demonstration of behavior; Social support (practical) | Training - Adults/Community-based |
| | Education; Training | Instruction on how to perform the behavior; Information about health consequences | Plays, skits or songs |
| | Education; Enablement | Commitment; Action planning; Information on others' approval | Public pledging ceremonies |
| | Environmental Restructuring | Restructuring the physical environmnent; Adding objects to the environment | HW station |
| Biran 2020 | Education; Training | Instruction on how to perform the behavior; Information about health consequences | Education - Adults/Community-based |
| | Training | Information on how to perform the behavior | Multimedia - Videos |
| | Enablement; Training | Demonstration of behavior; Social support (practical) | Training - Adults/Community-based |
| Biswas 2019 | Training | Information on how to perform the behavior | Multimedia - Videos |

**S12** – IA, BCTs and activities

| | | | |
|---|---|---|---|
| | Education; Training | Instruction on how to perform the behavior; Information about health consequences | School-based Education |
| | Training | Demonstration of behavior; Feedback on behavior; Instruction on how to perform the behavior | School-based Training |
| | Environmental Restructuring | Adding objects to the environment | Alcohol rub provision |
| Blanton 2010 | Environmental Restructuring | Restructuring the physical environmnent; Adding objects to the environment | HW station |
| | Training | Demonstration of behavior; Feedback on behavior; Instruction on how to perform the behavior | School-based Training |
| | Education; Training; Environmental Restructuring | Information about health consequences; Instruction on how to perform the behavior; Adding objects to the environment | Multimedia - Art/flipcharts |
| Bosomprah 2016 | Education; Training | Instruction on how to perform the behavior; Information about health consequences | Education - Adults/Community-based |
| Bowen 2013 | Education; Training | Instruction on how to perform the behavior; Information about health consequences | Education - Adults/Community-based |
| | Environmental Restructuring | Adding objects to the environment | Soap provision |
| Briceño 2017 | Education; Training; Enablement | Demonstration of behavior; Information on health consequences; Social support (practical) | CHW visits |
| | Environmental Restructuring | Restructuring the physical environmnent; Adding objects to the environment | HW station |
| | Education; Training; Environmental Restructuring | Information about health consequences; Instruction on how to perform the behavior; Adding objects to the environment | Multimedia - Multiple |
| Briere 2012 | Education; Training; Environmental Restructuring | Information about health consequences; Instruction on how to perform the behavior; Adding objects to the environment | Multimedia - Art/flipcharts |
| | Environmental Restructuring | Adding objects to the environment | Soap provision |
| | Enablement; Training | Demonstration of behavior; Social support (practical) | Training - Adults/Community-based |

**S12** – IA, BCTs and activities

| | | | |
|---|---|---|---|
| Bulled 2017 | Environmental Restructuring | Restructuring the physical environmnent; Adding objects to the environment | HW station |
| | Education; Training | Instruction on how to perform the behavior; Information about health consequences | School-based Education |
| Burke 2016 | Education; Training; Environmental Restructuring | Information about health consequences; Instruction on how to perform the behavior; Adding objects to the environment | Multimedia - Art/flipcharts |
| | Education; Training | Instruction on how to perform the behavior; Information about health consequences | School-based Education |
| Burns 2018 | Environmental Restructuring | Adding objects to the environment | Soap provision |
| Cairncross 2005 | Education; Training | Instruction on how to perform the behavior; Information about health consequences | Education - Adults/Community-based |
| Capps 2022 | Education; Training; Environmental Restructuring | Information about health consequences; Instruction on how to perform the behavior; Adding objects to the environment | Multimedia - Posters |
| Carabin 1999 | Training | Demonstration of behavior; Feedback on behavior; Instruction on how to perform the behavior | School-based Training |
| CardinaleLagomarsino 2017 | Environmental Restructuring; Training | Instruction on how to perform the behavior; Prompts/cues | Other |
| Chard 2018 | Environmental Restructuring | Restructuring the physical environmnent; Adding objects to the environment | HW station |
| | Education; Training | Instruction on how to perform the behavior; Information about health consequences | School-based Education |
| Contzen 2013 | Education; Enablement | Commitment; Action planning; Information on others' approval | Public pledging ceremonies |
| | Education; Training | Instruction on how to perform the behavior; Information about health consequences | Education - Adults/Community-based |
| | Environmental Restructuring | Restructuring the physical environmnent; Adding objects to the environment | HW station |
| Costa 2019 | Education; Training | Instruction on how to perform the behavior; Information about health consequences | School-based Education |

**S12** – IA, BCTs and activities

| | | | |
|---|---|---|---|
| Cowling 2009 | Environmental Restructuring | Adding objects to the environment | Alcohol rub provision |
| | Enablement; Training | Demonstration of behavior; Social support (practical) | Training - Adults/Community-based |
| | Environmental Restructuring | Adding objects to the environment | Soap provision |
| Croghan 2008 | Education; Training | Instruction on how to perform the behavior; Information about health consequences | School-based Education |
| | Environmental Restructuring | Prompts/cues; Adding objects to the environment | Soap provision |
| Curtis 2001 | Environmental Restructuring | Adding objects to the environment | Soap provision |
| | Education; Training | Instruction on how to perform the behavior; Information about health consequences | Education - Adults/Community-based |
| | Education; Training | Instruction on how to perform the behavior; Information about health consequences | Multimedia - radio |
| | Environmental Restructuring | Restructuring the physical environmnent; Adding objects to the environment | HW station |
| Davis 2011 | Enablement; Training | Demonstration of behavior; Social support (practical) | Training - Adults/Community-based |
| Davis 2013 | Education; Training; Environmental Restructuring | Information about health consequences; Instruction on how to perform the behavior; Adding objects to the environment | Multimedia - Posters |
| Ditai 2019 | Environmental Restructuring | Adding objects to the environment | Alcohol rub provision |
| | Education; Training; Environmental Restructuring | Information about health consequences; Instruction on how to perform the behavior; Adding objects to the environment | Multimedia - Posters |
| | Enablement; Training | Demonstration of behavior; Social support (practical) | Training - Adults/Community-based |
| Dreibelbis 2016 | Environmental Restructuring | Restructuring the physical environmnent; Adding objects to the environment | HW station |
| | Education; Training | Instruction on how to perform the behavior; Information about health consequences | School-based Education |
| Duijster 2020 | Environmental Restructuring | Adding objects to the environment | Soap provision |

| | | | |
|---|---|---|---|
| | Education; Training | Instruction on how to perform the behavior; Information about health consequences | Multimedia - Unspecified |
| | Environmental Restructuring | Adding objects to the environment | HW station |
| Early 1998 | Education; Training | Instruction on how to perform the behavior; Information about health consequences | School-based Education |
| | Environmental Restructuring | Adding objects to the environment | Alcohol rub provision |
| Ebuehi 2010 | Education; Training | Instruction on how to perform the behavior; Information about health consequences | Education - Adults/Community-based |
| | Enablement | Social support (unspecified); Restructuring the social environment | Policy or institutional strengthening |
| Edward 2019 | Education; Training; Enablement | Demonstration of behavior; Information on health consequences; Social support (practical) | CHW visits |
| ErcanOruc 2020 | Enablement; Training | Demonstration of behavior; Social support (practical) | Training - Adults/Community-based |
| Ercumen 2018 | Environmental Restructuring | Adding objects to the environment | Soap provision |
| | Environmental Restructuring | Restructuring the physical environmnent; Adding objects to the environment | HW station |
| Eun-Joo 2012 | Education; Training | Instruction on how to perform the behavior; Information about health consequences | School-based Education |
| EvansJr 2009 | Education; Training; Environmental Restructuring | Information about health consequences; Instruction on how to perform the behavior; Adding objects to the environment | Multimedia - Posters |
| | Environmental Restructuring | Adding objects to the environment | Alcohol rub provision |
| | Education; Training | Instruction on how to perform the behavior; Information about health consequences | Education - Adults/Community-based |
| Farhana 2022 | Environmental Restructuring | Restructuring the physical environmnent; Adding objects to the environment | HW station |
| | Education; Training; Environmental Restructuring | Information about health consequences; Instruction on how to perform the behavior; Adding objects to the environment | Multimedia - Art/flipcharts |

**S12** – IA, BCTs and activities

| | | | |
|---|---|---|---|
| | Enablement | Social support (unspecified); Restructuring the social environment | Policy or institutional strengthening |
| | Training | Demonstration of behavior; Feedback on behavior; Instruction on how to perform the behavior | School-based Training |
| Ford 2014 | Environmental Restructuring; Training | Instruction on how to perform the behavior; Prompts/cues | Other |
| | Environmental Restructuring | Adding objects to the environment | Soap provision |
| Freeman 2020 | Enablement; Training | Demonstration of behavior; Social support (practical) | Training - Adults/Community-based |
| Freeman 2022 | Enablement | Social support (unspecified); Restructuring the social environment | Policy or institutional strengthening |
| Friedrich 2018 | Education; Training | Instruction on how to perform the behavior; Information about health consequences | Education - Adults/Community-based |
| | Environmental Restructuring | Prompts/cues; Restructuring the physical environment; Adding objects to the environment | HW station |
| | Enablement; Training | Demonstration of behavior; Social support (practical) | Training - Adults/Community-based |
| | Education; Enablement | Action planning; Information on others; approval; Goal setting (behavior) | Group discussions |
| | Education; Training; Environmental Restructuring | Information about health consequences; Instruction on how to perform the behavior; Adding objects to the environment | Multimedia - Posters |
| Galiani 2016 | Education; Training | Instruction on how to perform the behavior; Information about health consequences | Education - Adults/Community-based |
| | Enablement; Training | Demonstration of behavior; Social support (practical) | Training - Adults/Community-based |
| | Education; Training | Instruction on how to perform the behavior; Information about health consequences | Multimedia - Unspecified |
| Gautam 2017 | Enablement; Training | Demonstration of behavior; Social support (practical) | Training - Adults/Community-based |
| | Education; Training | Instruction on how to perform the behavior; Information about health consequences | Plays, skits or songs |
| Gedamu 2022 | Environmental Restructuring; Training | Instruction on how to perform the behavior; Prompts/cues | Other |
| Geller 1980 | Training | Demonstration of behavior; Feedback on behavior; Instruction on how to perform the behavior | Food Hygiene Training |

**S12** – IA, BCTs and activities

| | | | |
|---|---|---|---|
| | Enablement | Social support (unspecified); Restructuring the social environment | Policy or institutional strengthening |
| George 2017 | Education; Training; Environmental Restructuring | Information about health consequences; Instruction on how to perform the behavior; Adding objects to the environment | Multimedia - pamphlets |
| | Environmental Restructuring | Restructuring the physical environmnent; Adding objects to the environment | HW station |
| George 2017 | Environmental Restructuring | Adding objects to the environment | Soap provision |
| Gimaiyo 2019 | Enablement; Training | Demonstration of behavior; Social support (practical) | Training - Adults/Community-based |
| Goel 2019 | Education; Enablement | Action planning; Information on others; approval; Goal setting (behavior) | Group discussions |
| | Enablement; Training | Demonstration of behavior; Social support (practical) | Training - Adults/Community-based |
| | Environmental Restructuring | Adding objects to the environment | Soap provision |
| Goel 2020 | Training | Demonstration of behavior; Feedback on behavior; Instruction on how to perform the behavior | School-based Training |
| Grace 2012 | Enablement; Training | Demonstration of behavior; Social support (practical) | Training - Adults/Community-based |
| | Education; Training; Environmental Restructuring | Information about health consequences; Instruction on how to perform the behavior; Adding objects to the environment | Multimedia - Posters |
| | Environmental Restructuring | Prompts/cues; Adding objects to the environment | Soap provision |
| Greene 2012 | Environmental Restructuring | Restructuring the physical environmnent; Adding objects to the environment | HW station |
| | Training | Demonstration of behavior; Feedback on behavior; Instruction on how to perform the behavior | School-based Training |
| Greenland 2016 | Education; Training | Instruction on how to perform the behavior; Information about health consequences | Multimedia - radio |
| | Education; Training | Instruction on how to perform the behavior; Information about health consequences | Education - Adults/Community-based |
| Grover 2018 | Education; Training | Instruction on how to perform the behavior; Information about health consequences | School-based Education |

| | | | |
|---|---|---|---|
| | Environmental Restructuring | Restructuring the physical environmnent; Adding objects to the environment | HW station |
| | Environmental Restructuring; Training | Instruction on how to perform the behavior; Prompts/cues | Other |
| Guo 2018 | Education; Training | Instruction on how to perform the behavior; Information about health consequences | Education - Adults/Community-based |
| | Education; Training; Environmental Restructuring | Information about health consequences; Instruction on how to perform the behavior; Adding objects to the environment | Multimedia - Posters |
| Hanson 2020 | Education; Training | Instruction on how to perform the behavior; Information about health consequences | Training - Adults/Community-based |
| | Education; Training | Instruction on how to perform the behavior; Information about health consequences | Multimedia - radio |
| Her 2019 | Education; Training; Environmental Restructuring | Information about health consequences; Instruction on how to perform the behavior; Adding objects to the environment | Multimedia - Posters |
| | Environmental Restructuring; Training | Instruction on how to perform the behavior; Prompts/cues | Other |
| Hetherington 2017 | Enablement | Social support (unspecified); Restructuring the social environment | Policy or institutional strengthening |
| | Training | Demonstration of behavior; Feedback on behavior; Instruction on how to perform the behavior | School-based Training |
| Huang 2021 | Environmental Restructuring | Restructuring the physical environmnent; Adding objects to the environment | HW station |
| Hurley 2021 | Education; Training | Instruction on how to perform the behavior; Information about health consequences | Multimedia - Unspecified |
| Hussam 2022 | Environmental Restructuring | Adding objects to the environment | Soap provision |
| Jafree 2023 | Enablement | Self-monitoring of behavior; Goal setting (behavior) | Diaries and journals |
| | Training | Information on how to perform the behavior | Multimedia - Videos |
| Jagals 2004 | Education; Enablement | Action planning; Information on others; approval; Goal setting (behavior) | Group discussions |

**S12** – IA, BCTs and activities

| | | | |
|---|---|---|---|
| Jetha 2021 | Education; Training; Incentivization | Information about health consequences; Demonstration of behavior; Material incentive | Games or competitions |
| | Education; Training; Environmental Restructuring | Information about health consequences; Instruction on how to perform the behavior; Adding objects to the environment | Multimedia - Art/flipcharts |
| Jinadu 2007 | Education; Training | Instruction on how to perform the behavior; Information about health consequences | Education - Adults/Community-based |
| Johnson 2003 | Education; Training; Environmental Restructuring | Information about health consequences; Instruction on how to perform the behavior; Adding objects to the environment | Multimedia - Posters |
| Judah 2009 | Education; Training; Environmental Restructuring | Information about health consequences; Instruction on how to perform the behavior; Adding objects to the environment | Multimedia - Posters |
| Kaewchana 2012 | Enablement; Training | Demonstration of behavior; Social support (practical) | Training - Adults/Community-based |
| | Education; Training | Instruction on how to perform the behavior; Information about health consequences | Education - Adults/Community-based |
| | Education; Training; Environmental Restructuring | Information about health consequences; Instruction on how to perform the behavior; Adding objects to the environment | Multimedia - Posters |
| | Environmental Restructuring | Prompts/cues; Adding objects to the environment | Soap provision |
| Kajjura 2019 | Education; Training | Instruction on how to perform the behavior; Information about health consequences | Education - Adults/Community-based |
| Kamm 2016 | Education; Training; Environmental Restructuring | Information about health consequences; Instruction on how to perform the behavior; Adding objects to the environment | Multimedia - Posters |
| | Education; Training | Instruction on how to perform the behavior; Information about health consequences | Education - Adults/Community-based |
| | Environmental Restructuring | Restructuring the physical environmnent; Adding objects to the environment | HW station |
| Kamm 2016 | Environmental Restructuring | Prompts/cues; Adding objects to the environment | Soap provision |

**S12** – IA, BCTs and activities

| | | | |
|---|---|---|---|
| Kang 2017 | Education; Training | Instruction on how to perform the behavior; Information about health consequences | Education - Adults/Community-based |
| Kapadia-Kundu 2014 | Education; Training | Instruction on how to perform the behavior; Information about health consequences | School-based Education |
| | Enablement | Self-monitoring of behavior; Goal setting (behavior) | Diaries and journals |
| Kariuki 2012 | Education; Training; Enablement | Demonstration of behavior; Information on health consequences; Social support (practical) | CHW visits |
| | Training | Information on how to perform the behavior | Multimedia - Videos |
| Karon 2017 | Environmental Restructuring | Prompts/cues; Restructuring the physical environment; Adding objects to the environment | HW station |
| | Enablement | Social support (unspecified); Restructuring the social environment | Policy or institutional strengthening |
| Kitsanapun 2019 | Education; Training; Environmental Restructuring | Information about health consequences; Instruction on how to perform the behavior; Adding objects to the environment | Multimedia - Posters |
| | Enablement; Training | Demonstration of behavior; Social support (practical) | Training - Adults/Community-based |
| | Education; Training | Instruction on how to perform the behavior; Information about health consequences | Education - Adults/Community-based |
| Koehn 2020 | Education; Training; Enablement | Demonstration of behavior; Information on health consequences; Social support (practical) | CHW visits |
| Kumar 2018 | Education; Training; Environmental Restructuring | Information about health consequences; Instruction on how to perform the behavior; Adding objects to the environment | Multimedia - Posters |
| | Education; Training | Instruction on how to perform the behavior; Information about health consequences | School-based Education |
| Labović 2023 | Training | Demonstration of behavior; Feedback on behavior; Instruction on how to perform the behavior | Food Hygiene Training |
| Lange 2022 | Enablement | Self-monitoring of behavior; Goal setting (behavior) | Multimedia - SMS |
| | Training | Demonstration of behavior; Feedback on behavior; Instruction on how to perform the behavior | School-based Training |
| Langford 2011 | Environmental Restructuring | Adding objects to the environment | Soap provision |

| | | | |
|---|---|---|---|
| | Education; Training; Environmental Restructuring | Information about health consequences; Instruction on how to perform the behavior; Adding objects to the environment | Multimedia - Posters |
| | Education; Training | Instruction on how to perform the behavior; Information about health consequences | Education - Adults/Community-based |
| Langford 2013 | Education; Enablement | Action planning; Information on others; approval; Goal setting (behavior) | Group discussions |
| | Environmental Restructuring | Prompts/cues; Adding objects to the environment | Soap provision |
| | Education; Training; Environmental Restructuring | Information about health consequences; Instruction on how to perform the behavior; Adding objects to the environment | Multimedia - Posters |
| Lapinski 2013 | Education; Training; Environmental Restructuring | Information about health consequences; Instruction on how to perform the behavior; Adding objects to the environment | Multimedia - Posters |
| Lawson 2019 | Education; Training; Environmental Restructuring | Information about health consequences; Instruction on how to perform the behavior; Adding objects to the environment | Multimedia - Posters |
| Lee 2020 | Education; Training; Environmental Restructuring | Information about health consequences; Instruction on how to perform the behavior; Adding objects to the environment | Multimedia - Multiple |
| | Education; Training | Instruction on how to perform the behavior; Information about health consequences | School-based Education |
| Leventhal 2016 | Education; Training | Instruction on how to perform the behavior; Information about health consequences | School-based Education |
| Lhakhang 2015 | Education; Training | Instruction on how to perform the behavior; Information about health consequences | Education - Adults/Community-based |
| | Environmental Restructuring | Prompts/cues; Adding objects to the environment | Soap provision |
| | Education; Enablement | Action planning; Information on others; approval; Goal setting (behavior) | Group discussions |
| Liu 2019 | Environmental Restructuring | Adding objects to the environment | Soap provision |

| | | | |
|---|---|---|---|
| | Education; Training | Instruction on how to perform the behavior; Information about health consequences | School-based Education |
| | Education; Training; Environmental Restructuring | Information about health consequences; Instruction on how to perform the behavior; Adding objects to the environment | Multimedia - Multiple |
| | Training | Demonstration of behavior; Feedback on behavior; Instruction on how to perform the behavior | School-based Training |
| Locks 2019 | Education; Training; Environmental Restructuring | Information about health consequences; Instruction on how to perform the behavior; Adding objects to the environment | Multimedia - Multiple |
| | Education; Training | Instruction on how to perform the behavior; Information about health consequences | Education - Adults/Community-based |
| Lubna 2014 | Education; Training | Instruction on how to perform the behavior; Information about health consequences | Education - Adults/Community-based |
| Luby 2001 | Environmental Restructuring | Adding objects to the environment | Soap provision |
| | Education; Training | Instruction on how to perform the behavior; Information about health consequences | Education - Adults/Community-based |
| | Environmental Restructuring | Restructuring the physical environmnent; Adding objects to the environment | HW station |
| Machado 2018 | Training | Demonstration of behavior; Feedback on behavior; Instruction on how to perform the behavior | Food Hygiene Training |
| | Training | Information on how to perform the behavior | Multimedia - Videos |
| Mackert 2013 | Education; Training; Environmental Restructuring | Information about health consequences; Instruction on how to perform the behavior; Adding objects to the environment | Multimedia - Posters |
| Mahfuza 2021 | Education; Training | Instruction on how to perform the behavior; Information about health consequences | Multimedia - radio |
| Makata 2021 | Education; Training | Instruction on how to perform the behavior; Information about health consequences | School-based Education |
| | Environmental Restructuring | Prompts/cues; Restructuring the physical environment; Adding objects to the environment | HW station |

**S12** – IA, BCTs and activities

| | | | |
|---|---|---|---|
| Malik 2022 | Training | Demonstration of behavior; Feedback on behavior; Instruction on how to perform the behavior | School-based Training |
| | Education; Training; Environmental Restructuring | Information about health consequences; Instruction on how to perform the behavior; Adding objects to the environment | Multimedia - Art/flipcharts |
| Manaseki-Holland 2021 | Enablement; Training | Demonstration of behavior; Social support (practical) | Training - Adults/Community-based |
| Mane 2017 | Training | Demonstration of behavior; Feedback on behavior; Instruction on how to perform the behavior | School-based Training |
| Mathew 2018 | Training | Demonstration of behavior; Feedback on behavior; Instruction on how to perform the behavior | School-based Training |
| | Education; Enablement | Action planning; Information on others; approval; Goal setting (behavior) | Group discussions |
| Maughan 2016 | Environmental Restructuring; Training | Instruction on how to perform the behavior; Prompts/cues | Other |
| Mbakaya 2019 | Environmental Restructuring | Restructuring the physical environmnent; Adding objects to the environment | HW station |
| | Education; Training; Environmental Restructuring | Information about health consequences; Instruction on how to perform the behavior; Adding objects to the environment | Multimedia - Posters |
| | Enablement | Social support (unspecified); Restructuring the social environment | Policy or institutional strengthening |
| | Training | Demonstration of behavior; Feedback on behavior; Instruction on how to perform the behavior | School-based Training |
| | Education; Training | Instruction on how to perform the behavior; Information about health consequences | School-based Education |
| | Environmental Restructuring | Adding objects to the environment | Soap provision |
| McGuire-Wolfe 2012 | Education; Training; Environmental Restructuring | Information about health consequences; Instruction on how to perform the behavior; Adding objects to the environment | Multimedia - Posters |
| | Environmental Restructuring | Adding objects to the environment | Alcohol rub provision |
| | Enablement; Training | Demonstration of behavior; Social support (practical) | Training - Adults/Community-based |

**S12** – IA, BCTs and activities

| | | | |
|---|---|---|---|
| Mendes 2020 | Enablement; Training | Demonstration of behavior; Social support (practical) | Training - Adults/Community-based |
| | Education; Training; Environmental Restructuring | Information about health consequences; Instruction on how to perform the behavior; Adding objects to the environment | Multimedia - Posters |
| | Environmental Restructuring | Adding objects to the environment | Alcohol rub provision |
| Mezaache 2021 | Education; Training | Instruction on how to perform the behavior; Information about health consequences | Education - Adults/Community-based |
| | Environmental Restructuring | Adding objects to the environment | Alcohol rub provision |
| Mohamed 2019 | Training | Demonstration of behavior; Feedback on behavior; Instruction on how to perform the behavior | School-based Training |
| Moll 2007 | Education; Training | Instruction on how to perform the behavior; Information about health consequences | Education - Adults/Community-based |
| Morse 2020 | Enablement; Training | Demonstration of behavior; Social support (practical) | Training - Adults/Community-based |
| | Training | Information on how to perform the behavior | Multimedia - Videos |
| | Environmental Restructuring | Restructuring the physical environmnent; Adding objects to the environment | HW station |
| Mott 2007 | Environmental Restructuring; Training | Instruction on how to perform the behavior; Prompts/cues | Other |
| | Enablement; Training | Demonstration of behavior; Social support (practical) | Training - Adults/Community-based |
| | Environmental Restructuring | Adding objects to the environment | Alcohol rub provision |
| | Education; Training; Environmental Restructuring | Information about health consequences; Instruction on how to perform the behavior; Adding objects to the environment | Multimedia - Posters |
| | Environmental Restructuring | Adding objects to the environment | Soap provision |
| Nagapraveen 2016 | Education; Training | Instruction on how to perform the behavior; Information about health consequences | Education - Adults/Community-based |
| | Environmental Restructuring | Adding objects to the environment | Soap provision |

**S12** – IA, BCTs and activities

| | | | |
|---|---|---|---|
| | Education; Training; Environmental Restructuring | Information about health consequences; Instruction on how to perform the behavior; Adding objects to the environment | Multimedia - pamphlets |
| Nair 2017 | Education; Training; Enablement | Demonstration of behavior; Information on health consequences; Social support (practical) | CHW visits |
| Naluonde 2019 | Training | Demonstration of behavior; Feedback on behavior; Instruction on how to perform the behavior | School-based Training |
| | Environmental Restructuring | Adding objects to the environment | Soap provision |
| Nandrup-Bus 2009 | Education; Training; Environmental Restructuring | Information about health consequences; Instruction on how to perform the behavior; Adding objects to the environment | Multimedia - Posters |
| | Incentivization | Incentive (outcome) | Other incentives provided |
| | Training | Demonstration of behavior; Feedback on behavior; Instruction on how to perform the behavior | School-based Training |
| Newton-Lewis 2021 | Education; Training; Enablement | Demonstration of behavior; Information on health consequences; Social support (practical) | CHW visits |
| NikRosmawati 2018 | Education; Enablement | Social support (practical); Information about health consequences | Food Hygiene Education |
| | Environmental Restructuring; Training | Instruction on how to perform the behavior; Prompts/cues | Other |
| Nuhu 2019 | Enablement; Training | Demonstration of behavior; Social support (practical) | Training - Adults/Community-based |
| Öncü 2021 | Training | Demonstration of behavior; Feedback on behavior; Instruction on how to perform the behavior | School-based Training |
| Oruc 2021 | Training | Demonstration of behavior; Feedback on behavior; Instruction on how to perform the behavior | Food Hygiene Training |
| Oswald 2014 | Environmental Restructuring | Restructuring the physical environmnent; Adding objects to the environment | HW station |
| Ozcan 2020 | Education; Training | Instruction on how to perform the behavior; Information about health consequences | Plays, skits or songs |
| | Training | Demonstration of behavior; Feedback on behavior; Instruction on how to perform the behavior | School-based Training |

| | | | |
|---|---|---|---|
| | Education; Training; Incentivization | Information about health consequences; Demonstration of behavior; Material incentive | Games or competitions |
| Patel 2012 | Environmental Restructuring | Restructuring the physical environmnent; Adding objects to the environment | HW station |
| | Education; Training | Instruction on how to perform the behavior; Information about health consequences | School-based Education |
| | Environmental Restructuring | Adding objects to the environment | Soap provision |
| Phuanukoonnon 2013 | Environmental Restructuring | Adding objects to the environment | Soap provision |
| Pickering 2013 | Environmental Restructuring | Adding objects to the environment | Alcohol rub provision |
| | Environmental Restructuring | Adding objects to the environment | Soap provision |
| | Training | Demonstration of behavior; Feedback on behavior; Instruction on how to perform the behavior | School-based Training |
| | Environmental Restructuring | Restructuring the physical environmnent; Adding objects to the environment | HW station |
| | Education; Training; Environmental Restructuring | Information about health consequences; Instruction on how to perform the behavior; Adding objects to the environment | Multimedia - Multiple |
| Pickering 2019 | Environmental Restructuring | Restructuring the physical environmnent; Adding objects to the environment | HW station |
| | Enablement; Training | Demonstration of behavior; Social support (practical) | Training - Adults/Community-based |
| | Environmental Restructuring | Adding objects to the environment | Soap provision |
| Pinfold 1990 | Environmental Restructuring | Restructuring the physical environmnent; Adding objects to the environment | HW station |
| Pokharel 2017 | Enablement; Training | Demonstration of behavior; Social support (practical) | Training - Adults/Community-based |
| Prado 2015 | Education; Enablement | Social support (practical); Information about health consequences | Food Hygiene Education |
| Prasetyo 2022 | Environmental Restructuring; Training | Instruction on how to perform the behavior; Prompts/cues | Other |

| | | | |
|---|---|---|---|
| | Education; Training; Environmental Restructuring | Information about health consequences; Instruction on how to perform the behavior; Adding objects to the environment | Multimedia - Posters |
| | Environmental Restructuring | Restructuring the physical environmnent; Adding objects to the environment | HW station |
| Ram 2017 | Environmental Restructuring; Training | Instruction on how to perform the behavior; Prompts/cues | Other |
| | Enablement; Training | Demonstration of behavior; Social support (practical) | Training - Adults/Community-based |
| | Environmental Restructuring | Prompts/cues; Restructuring the physical environment; Adding objects to the environment | HW station |
| | Education; Training; Environmental Restructuring | Information about health consequences; Instruction on how to perform the behavior; Adding objects to the environment | Multimedia - Posters |
| Ram 2020 | Environmental Restructuring | Adding objects to the environment | Soap provision |
| Ray 2010 | Enablement; Training | Demonstration of behavior; Social support (practical) | Training - Adults/Community-based |
| ReyesFernández 2015 | Education; Enablement | Action planning; Information on others; approval; Goal setting (behavior) | Group discussions |
| | Education; Training; Environmental Restructuring | Information about health consequences; Instruction on how to perform the behavior; Adding objects to the environment | Multimedia - Posters |
| Riaz 2016 | Enablement; Training | Demonstration of behavior; Social support (practical) | Training - Adults/Community-based |
| Rissman 2021 | Education; Training | Instruction on how to perform the behavior; Information about health consequences | Education - Adults/Community-based |
| Roberts 2008 | Training | Demonstration of behavior; Feedback on behavior; Instruction on how to perform the behavior | Food Hygiene Training |
| Roberts 2022 | Training | Information on how to perform the behavior | Multimedia - Videos |
| Rosen 2006 | Education; Training | Instruction on how to perform the behavior; Information about health consequences | School-based Education |
| Rosen 2006 | Environmental Restructuring | Adding objects to the environment | Soap provision |
| Routh 2018 | Environmental Restructuring | Adding objects to the environment | Soap provision |

**S12** – IA, BCTs and activities

| | | | |
|---|---|---|---|
| | Education; Training | Instruction on how to perform the behavior; Information about health consequences | Education - Adults/Community-based |
| Russo 2012 | Environmental Restructuring | Adding objects to the environment | Soap provision |
| | Education; Training | Instruction on how to perform the behavior; Information about health consequences | Multimedia - radio |
| Rutter 2020 | Education; Training; Environmental Restructuring | Information about health consequences; Instruction on how to perform the behavior; Adding objects to the environment | Multimedia - Posters |
| Saboori 2013 | Training | Demonstration of behavior; Feedback on behavior; Instruction on how to perform the behavior | School-based Training |
| | Environmental Restructuring | Adding objects to the environment | Soap provision |
| Samreen 2021 | Training | Demonstration of behavior; Feedback on behavior; Instruction on how to perform the behavior | School-based Training |
| Sanders 2021 | Environmental Restructuring | Prompts/cues; Restructuring the physical environment; Adding objects to the environment | HW station |
| | Education; Training | Instruction on how to perform the behavior; Information about health consequences | Education - Adults/Community-based |
| | Education; Training; Environmental Restructuring | Information about health consequences; Instruction on how to perform the behavior; Adding objects to the environment | Multimedia - Multiple |
| | Education; Training; Enablement | Demonstration of behavior; Information on health consequences; Social support (practical) | CHW visits |
| Sangalang 2021 | Environmental Restructuring | Adding objects to the environment | Soap provision |
| | Training | Demonstration of behavior; Feedback on behavior; Instruction on how to perform the behavior | School-based Training |
| | Education; Training | Instruction on how to perform the behavior; Information about health consequences | School-based Education |
| | Environmental Restructuring | Restructuring the physical environmnent; Adding objects to the environment | HW station |

**S12** – IA, BCTs and activities

| | | | |
|---|---|---|---|
| Sania 2017 | Environmental Restructuring | Adding objects to the environment | Soap provision |
| | Environmental Restructuring | Restructuring the physical environmnent; Adding objects to the environment | HW station |
| | Enablement; Training | Demonstration of behavior; Social support (practical) | Training - Adults/Community-based |
| Schroeder 2016 | Education; Training; Environmental Restructuring | Information about health consequences; Instruction on how to perform the behavior; Adding objects to the environment | Multimedia - Art/flipcharts |
| Scott 2008 | Education; Training | Instruction on how to perform the behavior; Information about health consequences | Multimedia - radio |
| | Education; Training | Instruction on how to perform the behavior; Information about health consequences | Education - Adults/Community-based |
| Shah 2021 | Training | Information on how to perform the behavior | Multimedia - Videos |
| Shahar 2022 | Education; Training | Instruction on how to perform the behavior; Information about health consequences | School-based Education |
| Sheth 2004 | Education; Training | Instruction on how to perform the behavior; Information about health consequences | School-based Education |
| Simiyu 2022 | Education; Training | Instruction on how to perform the behavior; Information about health consequences | Education - Adults/Community-based |
| Simmerman 2011 | Environmental Restructuring | Adding objects to the environment | Soap provision |
| | Education; Training | Instruction on how to perform the behavior; Information about health consequences | Education - Adults/Community-based |
| Sneed 2015 | Education; Training | Instruction on how to perform the behavior; Information about health consequences | Education - Adults/Community-based |
| | Training | Information on how to perform the behavior | Multimedia - Videos |
| Snow 2008 | Training | Demonstration of behavior; Feedback on behavior; Instruction on how to perform the behavior | School-based Training |
| | Education; Training | Instruction on how to perform the behavior; Information about health consequences | School-based Education |
| Soares 2013 | Training | Demonstration of behavior; Feedback on behavior; Instruction on how to perform the behavior | Food Hygiene Training |
| Sobel 2022 | Enablement; Training | Demonstration of behavior; Social support (practical) | Training - Adults/Community-based |

| | | | |
|---|---|---|---|
| | Training | Information on how to perform the behavior | Multimedia - Videos |
| Solehati 2017 | Education; Training; Environmental Restructuring | Information about health consequences; Instruction on how to perform the behavior; Adding objects to the environment | Multimedia - Posters |
| | Training | Demonstration of behavior; Feedback on behavior; Instruction on how to perform the behavior | School-based Training |
| | Enablement | Social support (unspecified); Restructuring the social environment | Policy or institutional strengthening |
| | Education; Training | Instruction on how to perform the behavior; Information about health consequences | School-based Education |
| Stebbins 2010 | Education; Training | Instruction on how to perform the behavior; Information about health consequences | School-based Education |
| | Environmental Restructuring | Adding objects to the environment | Alcohol rub provision |
| Stedman-Smith 2015 | Training | Information on how to perform the behavior | Multimedia - Videos |
| | Environmental Restructuring | Adding objects to the environment | Alcohol rub provision |
| Strohbehn 2011 | Education; Enablement | Social support (practical); Information about health consequences | Food Hygiene Education |
| | Training | Demonstration of behavior; Feedback on behavior; Instruction on how to perform the behavior | Food Hygiene Training |
| | Environmental Restructuring | Adding objects to the environment | Soap provision |
| | Education; Training; Environmental Restructuring | Information about health consequences; Instruction on how to perform the behavior; Adding objects to the environment | Multimedia - pamphlets |
| Suen 2020 | Training | Information on how to perform the behavior | Multimedia - Videos |
| | Education; Training | Instruction on how to perform the behavior; Information about health consequences | School-based Education |
| | Training | Demonstration of behavior; Feedback on behavior; Instruction on how to perform the behavior | School-based Training |
| Sutherland 2021 | Environmental Restructuring | Restructuring the physical environmnent; Adding objects to the environment | HW station |

| | | | |
|---|---|---|---|
| | Environmental Restructuring | Adding objects to the environment | Soap provision |
| Takanashi 2013 | Education; Training; Environmental Restructuring | Information about health consequences; Instruction on how to perform the behavior; Adding objects to the environment | Multimedia - pamphlets |
| Taware 2018 | Education; Training | Instruction on how to perform the behavior; Information about health consequences | School-based Education |
| | Education; Training; Environmental Restructuring | Information about health consequences; Instruction on how to perform the behavior; Adding objects to the environment | Multimedia - Posters |
| Thorseth 2021 | Environmental Restructuring | Adding objects to the environment | Soap provision |
| | Environmental Restructuring | Restructuring the physical environmnent; Adding objects to the environment | HW station |
| Tian 2019 | Education; Enablement | Action planning; Information on others; approval; Goal setting (behavior) | Group discussions |
| | Education; Training | Instruction on how to perform the behavior; Information about health consequences | Education - Adults/Community-based |
| | Enablement; Training | Demonstration of behavior; Social support (practical) | Training - Adults/Community-based |
| | Education; Training; Environmental Restructuring | Information about health consequences; Instruction on how to perform the behavior; Adding objects to the environment | Multimedia - Posters |
| Tidwell 2019 | Training | Information on how to perform the behavior | Multimedia - Videos |
| | Enablement | Self-monitoring of behavior; Goal setting (behavior) | Multimedia - SMS |
| Tidwell 2020 | Education; Training | Instruction on how to perform the behavior; Information about health consequences | School-based Education |
| Topan 2020 | Education; Training | Instruction on how to perform the behavior; Information about health consequences | School-based Education |
| Tousman 2007 | Education; Training | Instruction on how to perform the behavior; Information about health consequences | School-based Education |
| Tousman 2011 | Enablement | Self-monitoring of behavior; Goal setting (behavior) | Diaries and journals |
| Umair 2019 | Education; Training | Instruction on how to perform the behavior; Information about health consequences | Education - Adults/Community-based |

**S12** – IA, BCTs and activities

| | | | |
|---|---|---|---|
| Underwood 2017 | Education; Training | Instruction on how to perform the behavior; Information about health consequences | Plays, skits or songs |
| Updegraff 2011 | Education; Training; Environmental Restructuring | Information about health consequences; Instruction on how to perform the behavior; Adding objects to the environment | Multimedia - Posters |
| Vally 2019 | Education; Training | Instruction on how to perform the behavior; Information about health consequences | School-based Education |
| | Education; Training; Environmental Restructuring | Information about health consequences; Instruction on how to perform the behavior; Adding objects to the environment | Multimedia - Multiple |
| | Enablement | Social support (unspecified); Restructuring the social environment | Policy or institutional strengthening |
| | Environmental Restructuring | Prompts/cues; Restructuring the physical environment; Adding objects to the environment | HW station |
| | Environmental Restructuring | Adding objects to the environment | Soap provision |
| VazNery 2019 | Education; Training | Instruction on how to perform the behavior; Information about health consequences | Education - Adults/Community-based |
| Violant-Holz 2021 | Training | Demonstration of behavior; Feedback on behavior; Instruction on how to perform the behavior | School-based Training |
| Waterkeyn 2005 | Enablement; Training | Demonstration of behavior; Social support (practical) | Training - Adults/Community-based |
| Watson 2019 | Environmental Restructuring | Adding objects to the environment | Soap provision |
| | Enablement; Training | Demonstration of behavior; Social support (practical) | Training - Adults/Community-based |
| Weijers 2020 | Education; Training; Environmental Restructuring | Information about health consequences; Instruction on how to perform the behavior; Adding objects to the environment | Multimedia - Posters |
| White 2003 | Environmental Restructuring | Adding objects to the environment | Alcohol rub provision |
| | Education; Training; Environmental Restructuring | Information about health consequences; Instruction on how to perform the behavior; Adding objects to the environment | Multimedia - Posters |

**S12** – IA, BCTs and activities

| | | | |
|---|---|---|---|
| Wichaidit 2019 | Education; Training | Instruction on how to perform the behavior; Information about health consequences | Plays, skits or songs |
| | Enablement; Training | Demonstration of behavior; Social support (practical) | Training - Adults/Community-based |
| | Environmental Restructuring | Adding objects to the environment | Soap provision |
| Wichaidit 2019 | Environmental Restructuring | Restructuring the physical environmnent; Adding objects to the environment | HW station |
| | Education; Training | Instruction on how to perform the behavior; Information about health consequences | Education - Adults/Community-based |
| Wilson 1993 | Environmental Restructuring | Adding objects to the environment | Soap provision |
| | Education; Training; Environmental Restructuring | Information about health consequences; Instruction on how to perform the behavior; Adding objects to the environment | Multimedia - Multiple |
| | Enablement; Training | Demonstration of behavior; Social support (practical) | Training - Adults/Community-based |
| Witt 2004 | Training | Demonstration of behavior; Feedback on behavior; Instruction on how to perform the behavior | School-based Training |
| Wong 2022 | Environmental Restructuring; Training | Instruction on how to perform the behavior; Prompts/cues | Other |
| | Training | Demonstration of behavior; Feedback on behavior; Instruction on how to perform the behavior | Food Hygiene Training |
| Wu 2022 | Training | Demonstration of behavior; Feedback on behavior; Instruction on how to perform the behavior | School-based Training |
| | Education; Training | Instruction on how to perform the behavior; Information about health consequences | School-based Education |
| Yang 2017 | Training | Information on how to perform the behavior | Multimedia - Videos |
| | Enablement | Social support (unspecified); Restructuring the social environment | Policy or institutional strengthening |
| | Education; Training | Instruction on how to perform the behavior; Information about health consequences | Education - Adults/Community-based |
| | Environmental Restructuring | Restructuring the physical environmnent; Adding objects to the environment | HW station |

**S12** – IA, BCTs and activities

| | | | |
|---|---|---|---|
| Yardley 2011 | Education; Training | Instruction on how to perform the behavior; Information about health consequences | Education - Adults/Community-based |
| Yeboah-Antwi 2019 | Enablement | Social support (unspecified); Restructuring the social environment | Policy or institutional strengthening |
| York 2009 | Education; Training; Environmental Restructuring | Information about health consequences; Instruction on how to perform the behavior; Adding objects to the environment | Multimedia - Posters |
| | Education; Training; Incentivization | Information about health consequences; Demonstration of behavior; Material incentive | Games or competitions |
| Younie 2020 | Education; Training | Instruction on how to perform the behavior; Information about health consequences | School-based Education |
| Yu 2018 | Training | Information on how to perform the behavior | Multimedia - Videos |
| | Environmental Restructuring | Adding objects to the environment | Soap provision |
| | Incentivization | Incentive (outcome) | Other incentives provided |
| Zemichael 2020 | Education; Training | Instruction on how to perform the behavior; Information about health consequences | Education - Adults/Community-based |
| | Enablement | Social support (unspecified); Restructuring the social environment | Policy or institutional strengthening |
| | Environmental Restructuring | Restructuring the physical environmnent; Adding objects to the environment | HW station |
| | Education; Training; Environmental Restructuring | Information about health consequences; Instruction on how to perform the behavior; Adding objects to the environment | Multimedia - pamphlets |
| Zhang 2013 | Environmental Restructuring | Adding objects to the environment | Soap provision |
| | Education; Training | Instruction on how to perform the behavior; Information about health consequences | School-based Education |
| | Environmental Restructuring | Restructuring the physical environmnent; Adding objects to the environment | HW station |
| Zhang 2021 | Training | Demonstration of behavior; Feedback on behavior; Instruction on how to perform the behavior | School-based Training |
| | Enablement | Self-monitoring of behavior; Goal setting (behavior) | Multimedia - SMS |

| | | | |
|---|---|---|---|
| | Education; Training | Instruction on how to perform the behavior; Information about health consequences | School-based Education |
| Zomer 2016 | Environmental Restructuring | Adding objects to the environment | Alcohol rub provision |
| | Environmental Restructuring | Adding objects to the environment | Soap provision |
| | Education; Training; Environmental Restructuring | Information about health consequences; Instruction on how to perform the behavior; Adding objects to the environment | Multimedia - Posters |
| | Training | Demonstration of behavior; Feedback on behavior; Instruction on how to perform the behavior | School-based Training |
