## Supplementary material for "Interventions to improve hand hygiene in community settings: A systematic review of theories, barriers and enablers, behavior change techniques, and hand hygiene station design features": S13

**S13** – Hand hygiene station designs

**S13** – Hand hygiene station designs

***Supplementary File 13:***

Hand hygiene station types and designs features reported in included studies (N=223)

| Hand hygiene station types and designs features | Total n (%) | Effective** n (%) |
|---|---|---|
| **Studies reporting any handwashing station types and design features** | **46 (20.6)** | **37 (80.4)** |
| **Users** | | |
| School Populations | 24 (52.2) | 20 (83.3) |
| Household Populations | 15 (32.6) | 12 (80.0) |
| Community Members | 3 (6.5) | 2 (66.7) |
| Migrant Workers | 1 (2.2) | 1 (100.0) |
| Workplace Populations | 1 (2.2) | 1 (100.0) |
| Not reported | 2 (4.4) | 1 (50.0) |
| **Total Studies Reporting on Hand Washing Stations** | **46 (100.0)** | **37 (80.4)** |
| Raised bucket with tap/outlet | 20 (44.4) | 15 (75.0) |
| Sink with tap | 7 (15.6) | 7 (100.0) |
| Tippy tap | 4 (8.9) | 4 (100.0) |
| Free standing water tank with taps | 1 (2.2) | 1 (100.0) |
| Foot pump sink | 1 (2.2) | 0 ( 0.0) |
| Hand pump | 1 (2.2) | 1 (100.0) |
| Purpose-built all-in-one system | 1 (2.2) | 1 (100.0) |
| Two buckets suspended | 1 (2.2) | 1 (100.0) |
| Handwashing Station Type Not reported | 10 (22.2) | 7 (70.0) |
| **Mobility** | | |
| Mobile | 13 (28.3) | 9 (69.2) |
| Fixed | 12 (26.1) | 11 (91.7) |
| Not reported | 21 (45.7) | 17 (81.0) |
| **Permanency** | | |
| Temporary | 15 (32.6) | 11 (73.3) |
| Permanent | 11 (23.9) | 10 (90.9) |
| Not reported | 20 (43.5) | 16 (80.0) |
| **Water supply** | | |
| Individual storage tank | 13 (28.3) | 11 (84.6) |
| Piped water | 5 (10.9) | 5 (100.0) |
| Not reported | 28 (60.9) | 21 (75.0) |
| **Material** | | |
| Plastic | 7 (15.2) | 7 (100.0) |
| Not reported | 39 (84.8) | 30 (76.9) |
| **Location*** | | |
| Near baby's room | 2 (4.1) | 1 (50.0) |
| Near cooking area | 2 (4.1) | 0 (0.0) |
| Near construction sites | 1 (2.0) | 1 (100.0) |

| | | |
|---|---|---|
| Near markets | 1 (2.0) | 1 (100.0) |
| Near bathroom | 1 (2.0) | 0 (0.0) |
| Not reported | 42 (85.7) | 35 (83.3) |
| **Additional features** | | |
| Additional soap provided | 5 (10.9) | 4 (80.0) |
| Added cues to action (stickers, painted footpaths) | 3 (6.5) | 1 (33.3) |
| Bowl for run-off water provided | 3 (6.5) | 3 (100.0) |
| Additional water storage provided | 2 (4.4) | 1 (50.0) |
| Mirror | 2 (4.4) | 2 (100.0) |
| Aesthetic features | 1 (2.2) | 1 (100.0) |
| Addition features not reported | 30 (65.2) | 25 (83.3) |
| **Studies reporting handwashing station storage capacity (n=13)** | **Total mean (SD)** | **Effective mean (SD)** |
| Average storage capacity in L | 43.5 (29.8) | 46 (31.7) |

*Note: *Location is multi-select so could have multiple locations per handwashing station study; ** Effectiveness is determined if authors reported that the intervention was effective at improving hand hygiene outcomes*
