## Supplementary material for "Interventions to improve hand hygiene in community settings: A systematic review of theories, barriers and enablers, behavior change techniques, and hand hygiene station design features": S14

**S14** – HH station design adaptations

**S14** – HH station design adaptations

***Supplementary Table 14:***

Design adaptations of hand hygiene stations.

| Study ID, Country, Setting | Outcome of interest | Standard design | Design adaptation | Effectiveness* |
|---|---|---|---|---|
| Davis 2013** United States; Universities | Daily soap usage | Handwashing instructions on posters | **Cues to action** - Poster warnings of flu transmission | Adaptation performed worse |
| Lapinski 2013** United States; Universities | Observed handwashing | Low-prevalence poster ("One out of five college students wash their hands EVERY time they use the bathroom") | **Cues to action** - High-prevalence poster ("Four out of five college students wash their hands EVERY time they use the bathroom") | Adaptation performed better |
| Ford 2014** United States; Universities | Weekly soap consumption | Motion-activated towel dispenser | **Cues to action** - Towel was automatically available without action from users | Adaptation performed better |
| Dreibelbis 2016 Bangladesh; Schools | Observed handwashing after toileting events | No nudges | **"Nudges"***** - Paved footpath with painted handwashing station | Adaptation performed the same |
| Grover 2018 Bangladesh; Schools | Observed handwashing after toileting events | No nudges | **"Nudges"***** - Paved footpath, painted handwashing station, and painted shoeprints | Adaptation performed the same |
| Huang 2021 Philippines; Schools | Observed handwashing with soap after toilet use | No painted footpath or other nudges added | **"Nudges"***** - Painted footpath with an arrow sticker | Adaptation performed better |
| Prasetyo 2022 Indonesia; Workplaces | Observed handwashing with water or soap | Sink alone | **"Nudges"*** -** Bright yellow footprints painted | Adaptation performed better |
| Weijers 2020** The Netherlands; Markets | Observed hand disinfection upon entering store | Message on dispenser | **"Nudges"***** - Message combined with three blue arrows pointing to dispenser | Adaptation performed better |
| Bai 2022 China; Schools | Observed handwashing rates | Simple background with manual faucet | **Placement** – Aesthetic background (nature/wood) with spotlight and automatic faucet | Adaptation performed better |
| Thorseth 2021 Ethiopia; Internally displaced people camps | Observed handwashing with soap at critical events (after defecation, before preparing food, before eating, before serving/feeding another person food, and after | No additions | **Placement** - Addition of a mirror | Adaptation performed worse |

| | |
|---|---|
| | cleaning a child's bottom) |

***Notes:*** *Grey highlights are adaptations that performed better; *Effectiveness is determined if authors reported that the intervention was effective at improving hand hygiene outcomes; **Interventions that did not provide a handwashing station but used cues and "nudges" to modify/adapt an existing handwashing station; ***Interventions are classified as "nudges" as study authors used the term to characterize their own work;*
