## Supplementary material for "Interventions to improve hand hygiene in community settings: A systematic review of theories, barriers and enablers, behavior change techniques, and hand hygiene station design features": S15

**S15** – Frequency and Intensity

***Supplementary Table 15:*** Variations in frequency and intensity of hand hygiene interventions

| **Study ID Country, Setting** | **Outcome of interest** | **Standard intervention** | **Intervention adaptation** | **Effectiveness*** |
|---|---|---|---|---|
| Dreibelbis 2016 Bangladesh; Schools | Observed handwashing after toileting events | Handwashing station built first before hygiene education provision | **Sequencing** - Hygiene education provided at same time as handwashing station provision | Adaptation performed the same |
| Amon-Tanoh 2021 Côte d'Ivoire; Households | Observed handwashing with soap after toilet use | Handwashing station only | **Intensity -** Handwashing station combined with hygiene promotion activities (videos, Glo Germ demonstrations, posters) | Adaptation performed better |
| Biran 2020 Nigeria; Households | Observed handwashing with soap at key events (before eating or serving a meal, after defecation or latrine-use and after cleaning a child's bottom post-defecation) | Standard community-led total sanitation (CLTS) | **Intensity –** CLTS+, including additional discussions, skits, films, pledges, stickers, and report cards | Adaptation performed better |
| Freeman 2020 Kenya; Households | Households with a functional handwashing station | Standard program (THRIVE II) Training | **Intensity** - THRIVE II Training plus household visits from social workers and community health volunteers | Adaptation performed better |
| Nuhu 2019 Bangladesh; Households | Compliance with handwashing steps | Simple handwashing instructions | **Intensity** - Complex handwashing instructions | Adaptation performed the same |
| Sangalang 2021 Philippines; Schools | Handwashing practice score | WASH policy workshop for teachers and two health education sessions for students | **Intensity** - Medium-intensity (three 1-hr sessions) and high-intensity education sessions (four 1-hr sessions) | Adaptation performed worse |

***Notes:*** *Grey highlights are adaptations that performed better; *Effectiveness is determined if authors reported that the intervention was effective at improving hand hygiene outcomes*
