## Supplementary material for "Interventions to improve hand hygiene in community settings: A systematic review of theories, barriers and enablers, behavior change techniques, and hand hygiene station design features": S16

**S16** – Population groups, risk scenario and time bar charts

**S16** – Population groups, risk scenario and time bar charts

### *Supplemental file 16:*

Proportion of studies reporting effective interventions stratified by (A) key population groups, (B) vulnerable groups, (C) risk scenarios, and (D) data points collected post-intervention.

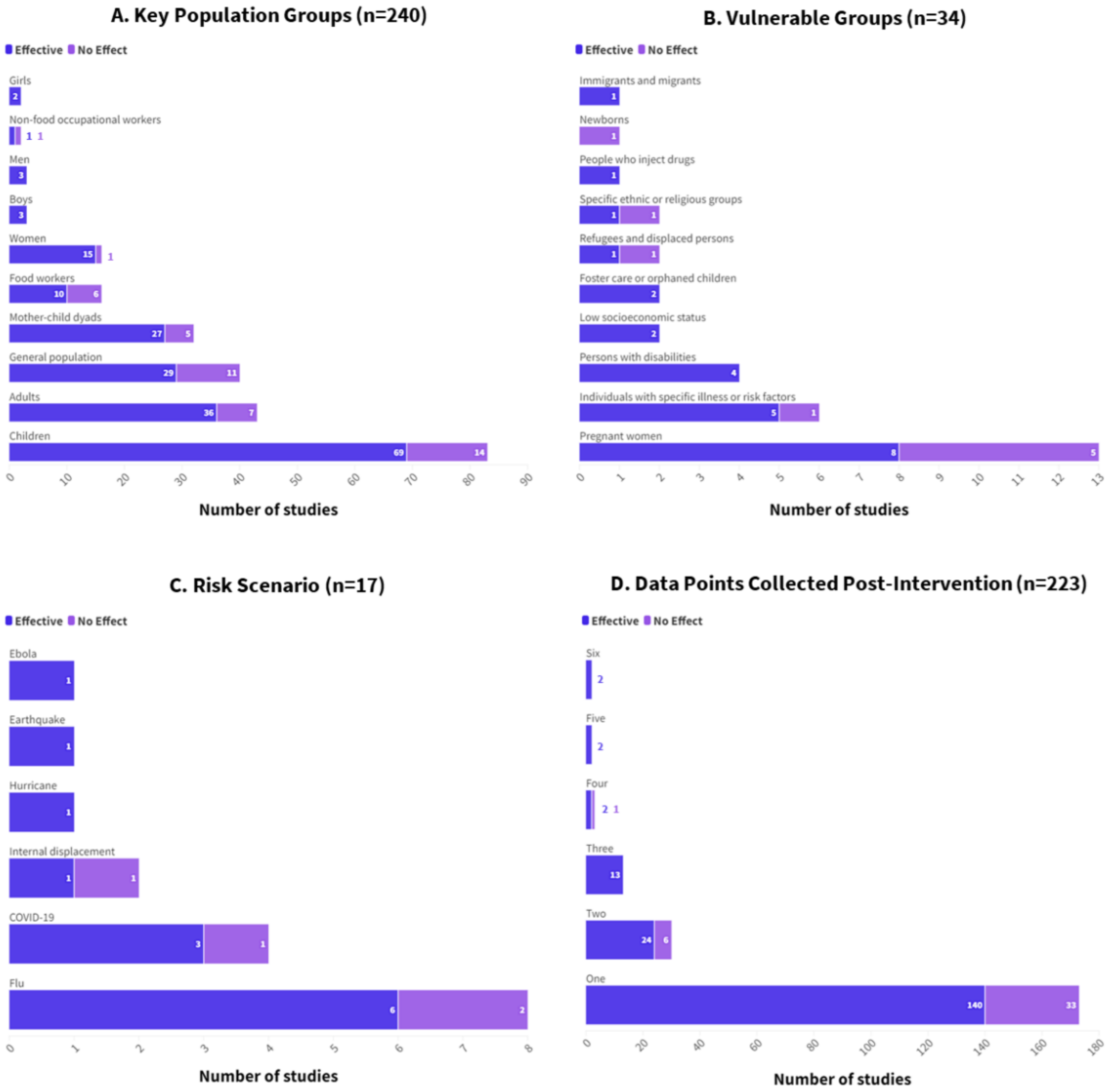
