## Supplementary material for "Interventions to improve hand hygiene in community settings: A systematic review of theories, barriers and enablers, behavior change techniques, and hand hygiene station design features": S17

**S17** – Full reference list

***Supplementary file 17:***

Full References of included studies

32. Bickford AR, Lee JH, Borzekowski DLG. Cleaner, Happier, Healthier: Sesame Workshop’s Water, Sanitation, and Hygiene Intervention among Low-Income Groups in Bangladesh and

India. Frontiers in Communication [Internet]. 2017;2. Available from: https://www.scopus.com/inward/record.uri?eid=2-s2.0-85096139069&doi=10.3389%2ffcomm.2017.00020&partnerID=40&md5=6b2ac6dcaabc71b1a47cca73be5dbd77

72. Ercan Oruc D, Pokharel S, Anantheswaran RC, Bucknavage MW, Gourama H, Shanina O, et al. A comprehensive food safety short course (FSSC) improves food safety knowledge,

behaviors, attitudes, and skills of Ukrainian participants. Journal of Food Science Education. 2020;19(4):263–77.

130. Lee RLT, Leung C, Chen H, Lee PH, Kwok SWH. A cluster randomized controlled trial of a simplified 5-step handwashing technique versus a conventional 7-step handwashing technique

among Chinese students with intellectual disabilities. Journal of Applied Research in Intellectual Disabilities. 2020;33(5):1090–9.

191. Roberts KR, Paez P, Sauer K, Alcorn M, Johnson DE. Impact of Training on Employees' Handwashing Behaviors in School Nutrition Programs. J Acad Nutr Diet. 2022;

207. Shah SN, Shah D, Desai N, Shah SH, Bhowmick S. Analysis of change in knowledge, attitude, and practices about COVID-19 following and awareness session in rural population of Western India. Ind Psychiatry J. 2021;30(Suppl 1):S35-s40.

222. Sutherland C, Reynaert E, Sindall RC, Riechmann ME, Magwaza F, Lienert J, et al. Innovation for improved hand hygiene: Field testing the Autarky handwashing station in

collaboration with informal settlement residents in Durban, South Africa. Sci Total Environ. 2021;796:149024.
